# Prevalence and disease expression of pathogenic and likely pathogenic variants associated with inherited cardiomyopathies in the general population

**DOI:** 10.1101/2022.01.06.22268837

**Authors:** Mimount Bourfiss, Marion van Vugt, Abdulrahman I. Alasiri, Bram Ruijsink, Jessica van Setten, Amand F. Schmidt, Dennis Dooijes, Esther Puyol-Antón, Birgitta K. Velthuis, J. Peter van Tintelen, Anneline S.J.M. te Riele, Annette F. Baas, Folkert W. Asselbergs

## Abstract

**Background:** Pathogenic and likely pathogenic variants associated with arrhythmogenic right ventricular cardiomyopathy (ARVC), dilated cardiomyopathy (DCM) and hypertrophic cardiomyopathy (HCM) are recommended to be reported as secondary findings in genome sequencing studies. This provides opportunities for early diagnosis, but also fuels uncertainty in variant carriers (G+), since disease penetrance is incomplete. We assessed the prevalence and disease expression of G+ in the general population.

**Methods:** We identified pathogenic and likely pathogenic variants associated with ARVC, DCM and/or HCM in 200,643 UK Biobank individuals, who underwent whole exome sequencing. We calculated the prevalence of G+ and analysed the frequency of cardiomyopathy/heart failure diagnosis. In undiagnosed individuals, we analysed early signs of disease expression using available electrocardiography and cardiac magnetic resonance imaging data.

**Results:** We found a prevalence of 1:578, 1:251 and 1:149 for pathogenic and likely pathogenic variants associated with ARVC, DCM and HCM respectively. Compared to controls, cardiovascular mortality was higher in DCM G+ (OR 1.67 [95% CI 1.04;2.59], p=0.030), but similar in ARVC and HCM G+ (p≥0.100). Cardiomyopathy or heart failure diagnosis were more frequent in DCM G+ (OR 3.66 [95% CI 2.24;5.81], p=4.9×10^−7^) and HCM G+ (OR 3.03 [95% CI 1.98;4.56], p=5.8×10^−7^), but comparable in ARVC G+ (p=0.172). In contrast, ARVC G+ had more ventricular arrhythmias (p=3.3×10^−4^). In undiagnosed individuals, left ventricular ejection fraction was reduced in DCM G+ (p=0.009).

**Conclusions:** In the general population, pathogenic and likely pathogenic variants associated with ARVC, DCM or HCM are not uncommon. Although G+ have increased mortality and morbidity, disease penetrance in these carriers from the general population remains low (1.2-3.1%). Follow-up decisions in case of incidental findings should not be based solely on a variant, but on multiple factors, including family history and disease expression.

## Background

The major inherited cardiomyopathies arrhythmogenic right ventricular cardiomyopathy (ARVC), dilated cardiomyopathy (DCM) and hypertrophic cardiomyopathy (HCM) are characterized by ventricular dysfunction and ventricular arrhythmias that can lead to progressive heart failure and sudden cardiac death^1^. ARVC is mainly caused by pathogenic variants in desmosomal genes, whereas DCM and HCM are mainly caused by sarcomeric gene variants^2^. These cardiomyopathies are typically inherited in an autosomal dominant fashion with incomplete penetrance and variable expressivity. As such, phenotypic expression may vary greatly, even among family members or individuals carrying the same pathogenic variant.

With the implementation of next-generation sequencing (NGS), genetic testing has become an important part of routine clinical care in the diagnosis of inherited cardiomyopathies^3^. Technical advances and commercial availability of NGS, have led to more affordable and accessible genetic testing. The American College of Medical Genetics and Genomics (ACMG) has developed recommendations for the reporting of incidental or secondary findings unrelated to the test indication^4^. In this framework, variants in genes associated with ARVC, DCM and HCM are recommended to be reported as secondary findings from clinical exome and other genome sequencing tests^4^. Although this offers the potential to prevent morbidity and mortality of heart failure and sudden cardiac death by early treatment, it also fuels uncertainty in carriers of likely pathogenic or pathogenic variants (G+) and their family members, since factors that influence disease penetrance in the general population are largely unknown. More knowledge about disease penetrance of these variants in an unselected population cohort is needed to determine screening protocols in these individuals.

In this study, we aimed to assess the prevalence of pathogenic and likely pathogenic variants in the general population using a set of recently curated genes for ARVC^5^, DCM^6^ and HCM^7^ in two (inter)national databases^8^ (see **Figure 1** and **Table I**). In order to do so, we leveraged data from the UK Biobank (UKB), a population-based cohort with whole exome sequencing (WES) data available of 200,643 individuals^9^. Furthermore, we looked into the UKB-reported phenotypical characteristics of these G+ and assessed the occurrence of early signs of disease in undiagnosed G+ using available electrocardiography and cardiac magnetic resonance (CMR) imaging data.

**Figure 1:**
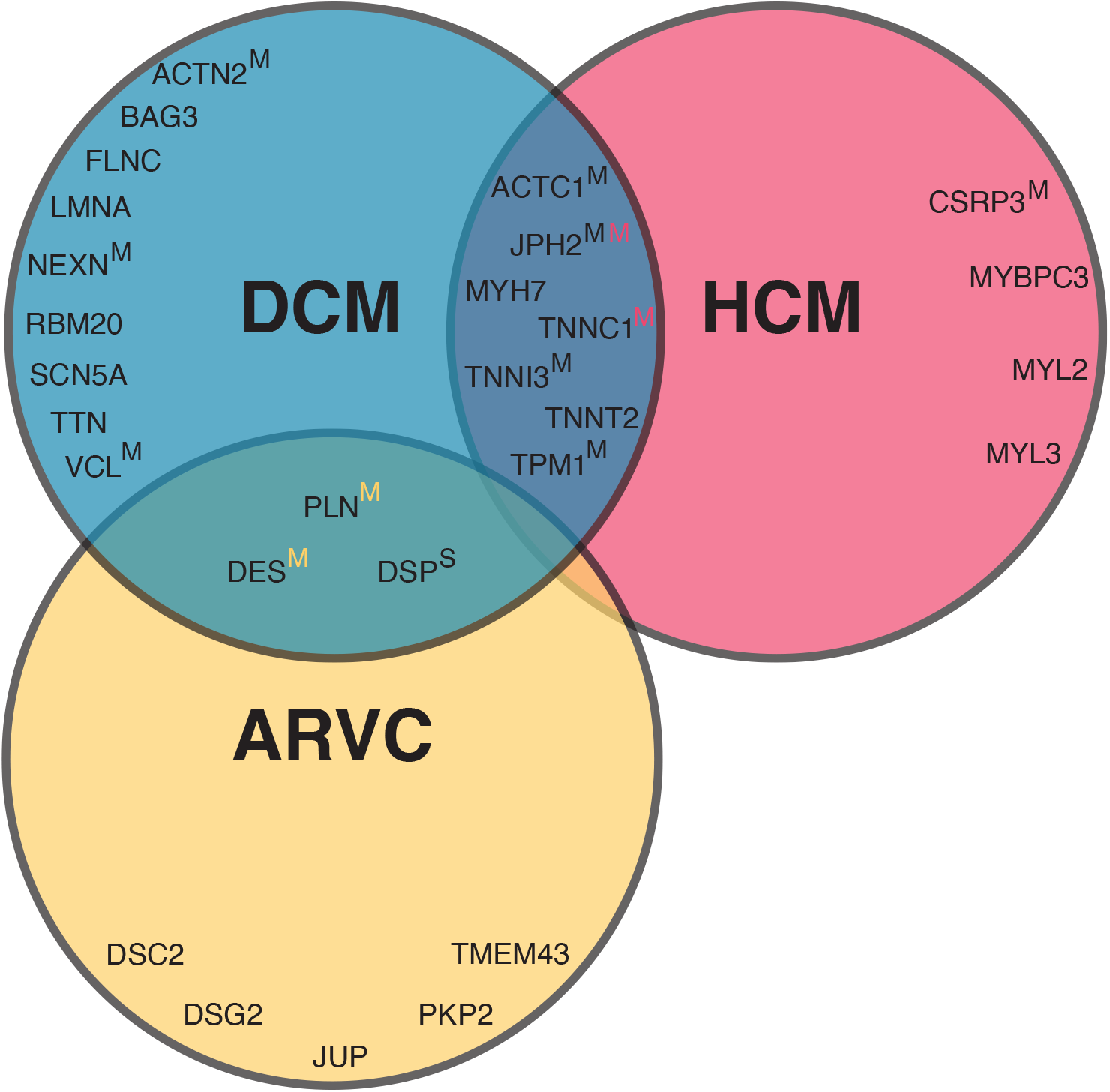
Included curated genes per cardiomyopathy. The Venn diagram of curated genes included in this study shows the overlap in genes per cardiomyopathy. Unless otherwise indicated, pathogenicity of genes are classified as definitive. If a superscript S or M is given, genes are classified as having a strong or moderate pathogenicity respectively. In the overlapping circles, yellow, black and red colors refer to ARVC, DCM, and HCM respectively. **Table I** *gives an overview of the included genes and pathogenicity classification per gene and abbreviation per gene*. *Abbreviations: ARVC= arrhythmogenic right ventricular cardiomyopathy; DCM= dilated cardiomyopathy; HCM= hypertrophic cardiomyopathy*.

## Methods

Ethics approval for the UKB study was obtained from the North West Centre for Research Ethics Committee (11/NW/0382) and all participants provided informed consent. All data and materials have been made publicly available on Github and can be accessed at https://github.com/CirculatoryHealth/Inherited-cardiomyopathies. Full methods are available in the **Supplemental Material**. Disease definitions are given in **Table II**.

## Results

### Population characteristics

We identified 2,207/200,643 unique G+ individuals from a total of 2,493 included individuals (89%) (see **Figure 2**) classified as 1) ARVC G+ (n=347, 54% female, median age of 57 [50-64] years); 2) DCM G+ (n=800, 56% female, median age of 58 [51-64] years) and; 3) HCM G+ (n=1,346, 54% female, median age of 56 [49-63] years). The matched control group consisted of 9,972 individuals (55% females, median age of 57 [49-63] years). **Table 1** and **Table III** show the baseline characteristics of the included individuals. The majority of G+ were of white ethnicity (ARVC 90%, DCM 96%, HCM 75%), followed by Asian (ARVC 3%, DCM 1%, HCM 19%) and black ethnicity (ARVC 2%, DCM 2% and HCM 2%). This is comparable to what is observed in the UKB, where the majority is of white ethnicity (94%), followed by Asian (2%) and black ethnicity (2%).

**Table 1:**
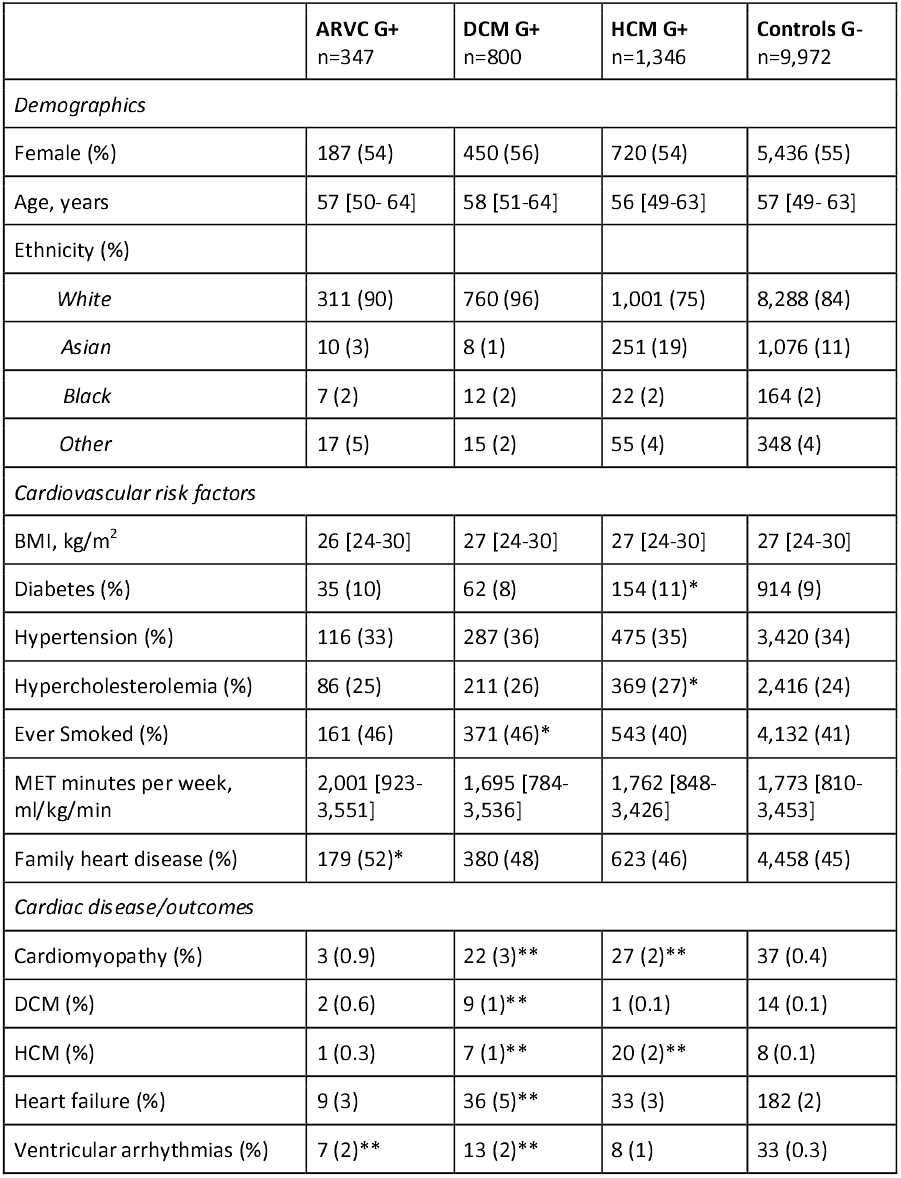
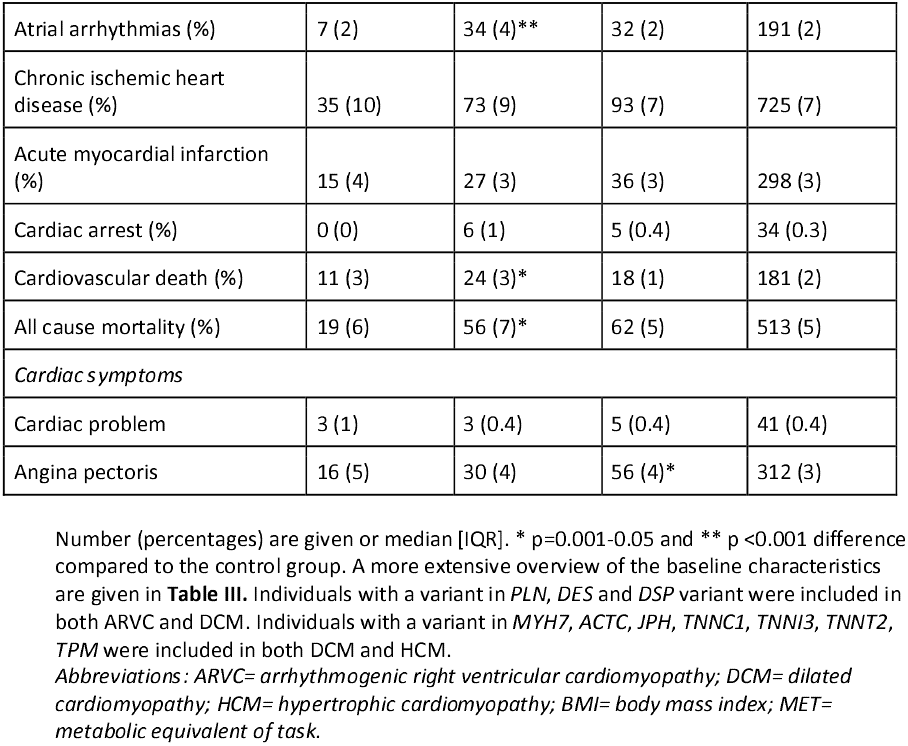
Baseline characteristics of variant carriers and controls.

**Figure 2:**
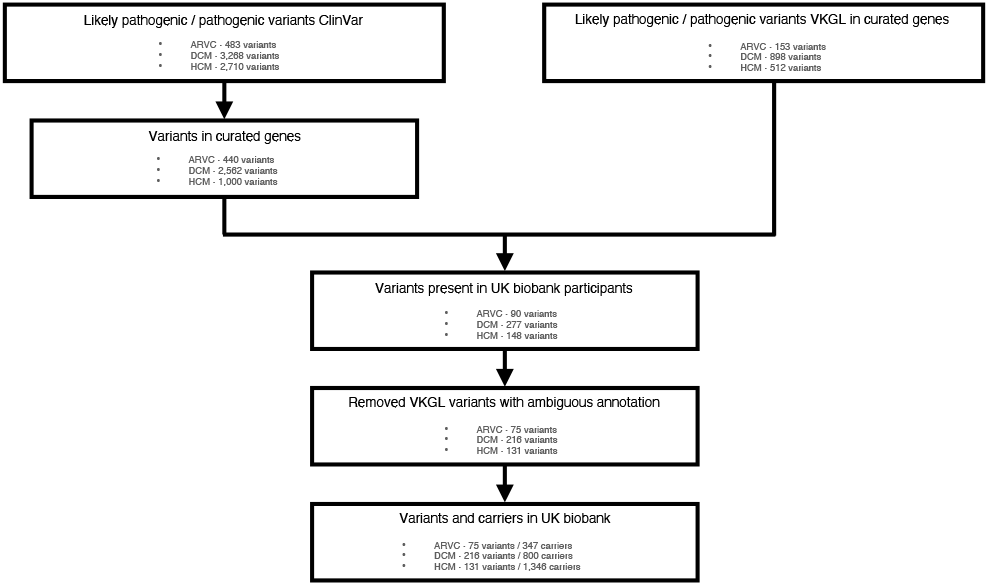
Flowchart inclusion of variants. Flowchart depicting the inclusion of pathogenic and likely pathogenic variants associated with arrhythmogenic cardiomyopathy, dilated cardiomyopathy and hypertrophic cardiomyopathy from the ClinVar^8^ and VKGL database. *Abbreviations: ARVC= arrhythmogenic right ventricular cardiomyopathy; DCM= dilated cardiomyopathy; HCM= hypertrophic cardiomyopathy; VKGL= Vereniging Klinische Genetische Laboratoriumdiagnostiek*.

### Genotypic characteristics of pathogenic and likely pathogenic variant carriers

#### Prevalence of pathogenic and likely pathogenic variants in the general population

We found a prevalence of 1 ARVC G+ in 578 people in the general population (1:578 [1:521; 1:644]) and identified 75 variants out of the 593 (13%) pathogenic and likely pathogenic variants described in ClinVar and VKGL: 13 missense and 62 loss of function (LoF) (**Table IV**). As shown in **Figure 3**, most ARVC G+ harbored a pathogenic variant in *PKP2* (n=185, 53%), followed by *DSP* (n=49, 14%), *DSC2* (n=42, 12%), *DSG2* (n=31, 9%), *JUP* (n=24, 7%), *DES* (n=15, 4%), and *PLN* (n=1, 0.3%).

**Figure 3:**
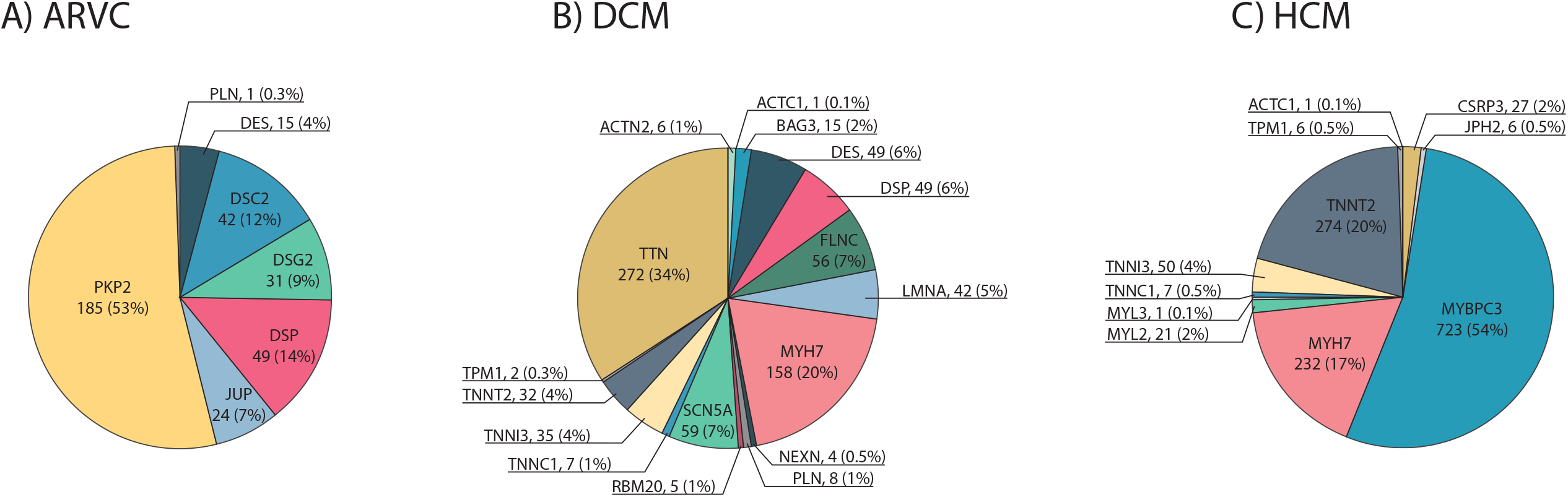
Distribution of genes per cardiomyopathy. Piecharts with the distribution of curated genes for A) arrhythmogenic right ventricular cardiomyopathy (ARVC); B) dilated cardiomyopathy (DCM); C) hypertrophic cardiomyopathy (HCM). *Abbreviations of the different genes are given in* ***Table III*** *Abbreviations: G+= pathogenic variant carrier*.

We found a prevalence of 1:251 [1:234; 1:269] DCM G+ and identified 216 out of 3,460 (6%) pathogenic and likely pathogenic variants described in ClinVar and VKGL: 80 missense and 136 LoF (**Table IV**). Variants in *TTN* (n=272, 34%) and *MYH7* (n=158, 20%) were most prevalent among DCM G+, followed by *SCN5A* (n=59, 7%), *FLNC* (n=56, 7%), *DSP* (n=49, 6%), *DES* (n=49, 6%), *LMNA* (n=42, 5%), *TNNI3* (n=35, 4%) and *TNNT2* (n=32, 4%). Eight more genes with a frequency of less than 3% were identified: *BAG3, PLN, TNNC1, ACTN2, RBM20, NEXN, TPM1*, and *ACTC1* (**Figure 3**).

We found a prevalence of 1:149 [1:141; 1:157] HCM G+ and identified 131 out of 1,512 (9%) pathogenic and likely pathogenic variants from ClinVar and VKGL: 98 missense and 23 LoF (**Table IV**). Most individuals carried a pathogenic and likely pathogenic variant in *MYBPC3* (n=723, 54%), followed by *TNNT2* (n=274, 20%), *MYH7* (n=232, 17%), and *TNNI3* (n=50, 4%). A frequency of less than 3% was found in *CSRP3, MYL2, TNNC1, JPH2, TPM1, ACTC1*, and *MYL3* (**Figure 3**). Interestingly, a variant in *TNNT2* (c.862C>T p.Arg288Cys) affected 242 individuals (18%). Four of these carriers were diagnosed with heart failure, of whom one also with HCM. All four heart failure patients also suffered from chronic ischemic heart disease. Furthermore, a variant in *MYBPC3* (c.3628-41_3628-17del) was mainly seen in individuals with an Asian ethnicity (n=237, 18% of the total HCM G+). Four of these individuals were diagnosed with heart failure, of whom two also had coronary artery disease and one was diagnosed with DCM, however none were diagnosed with HCM. When excluding these two variants, we found a HCM G+ prevalence of 1:250 [1:234; 1:269]. *MYBPC3* remained the most prevalent gene (52%), whereas the *TNNT2* frequency decreased to 4%.

The prevalence of G+ per gene for all cardiomyopathies is depicted in **Table V**.

#### Overlapping variants and individuals

Some pathogenic and likely pathogenic variants were identified in multiple cardiomyopathy subtypes. First, 26 variants were described in both ARVC and DCM, affecting 53 individuals. Most of these variants (n=20/26 variants, 77%) were found in *DSP* (n=37 individuals, 70%), of whom one individual (3%) had heart failure and one (3%) was diagnosed with a cardiomyopathy. Five variants out of 26 (19%) were found in *DES* (n=15 individuals, 28%) of whom two individuals (13%) had heart failure, and one was diagnosed with both DCM and HCM. One variant out of 26 (4%) was found in *PLN* (c.26G>A; p.Arg9His, NM_002667.5) in one individual (2%) who was not diagnosed with a cardiomyopathy or heart failure.

Second, 52 variants were described in DCM and HCM, affecting 232 individuals. Most of these variants (n=33/52 variants, 63%) were found in *MYH7* (n=158 individuals, 68%), followed by 10 variants (19%) in *TNNT2* (n=29 individuals, 13%), 6 variants (12%) in *TNNI3* (n=35 individuals, 15%), and 1 (2%) variant in *TNNC1, ACTC1* as well as *TPM1*. In this group of 232 individuals, 9 (4%) individuals had a cardiomyopathy or heart failure diagnosis, of whom 5 were diagnosed with HCM and none with DCM.

Furthermore, three individuals carried two pathogenic variants. Individual 1 was a 65-year old male, carrying variants in *MYBPC3* (c.3628-41_3628-17del, NM_000256.3) and *TNNT2* (c.460C>T; p.Arg154Trp, NM_001276345.2) and was diagnosed with heart failure, with underlying chronic ischemic heart disease. Individual 2 was a 65-year old male, carrying variants in *FLNC* (c.7450G>A; p.Gly2484Ser, NM_001458.5) and *PKP2* (c.1867G>T; p.Glu623Ter, NM_001005242.3) and was therefore included in both the ARVC as well as the DCM G+ group. Individual 3 was a 64-year old male, carrying variants in *MYBPC3* (c.1504C>T; p.Arg502Trp, NM_000256.3) and *MYH7* (c.5655G>A; p.Ala1885=, NM_000257.4). Individuals 2 and 3 were not diagnosed with a cardiomyopathy or heart failure and none had CMR data available.

**Table VI** shows the prevalence of the cardiomyopathy variants, with and without the inclusion of overlapping variants.

### Phenotypic characteristics of pathogenic and likely pathogenic variant carriers

#### Cardiovascular risk factors

Hypertension, BMI, and level of activity in metabolic equivalent of task minutes (MET) per week were comparable between G- and G+ for all cardiomyopathies (p≥0.055; **Table 1** and **Table VII**). Diabetes was more prevalent in G+ HCM (9.2% (G-) vs 11.4% (G+), p=0.008), while smoking was more prevalent in DCM G+ (41.4% vs 46.4%, p=0.007) (**Table VIII**).

#### Cardiovascular disease

As seen in **Figure 4** and **Table VIII**, compared to G-, cardiomyopathy/heart failure without previous ischemic heart disease (P+, phenotype positive) was more often diagnosed in DCM G+ (OR 3.66 [95% CI 2.24;5.81], p=4.9×10^−7^) and HCM G+ (OR 3.03 [95% CI 1.98;4.56], p=5.8×10^−7^). Among DCM G+, 25 individuals (3.1%, genes: 8 *MYH7*, 8 *TTN*, 2 *BAG3*, 2 *DSP*, 2 *FLNC*, 1 *DES*, 1 *SCN5A* and 1 *TNNT2*) were P+. Within HCM G+, 32 individuals (4.0%, genes: 20 *MYBPC3*, 10 *MYH7*, 1 *TNNI3* and 1 *TNNT2*) were P+. For ARVC G+, 4 individuals (1.2%, genes: 2 *DSP*, 1 *DES* and 1 *PKP2*) were P+, being comparable to G- controls (87 subjects, 0.8%).

**Figure 4:**
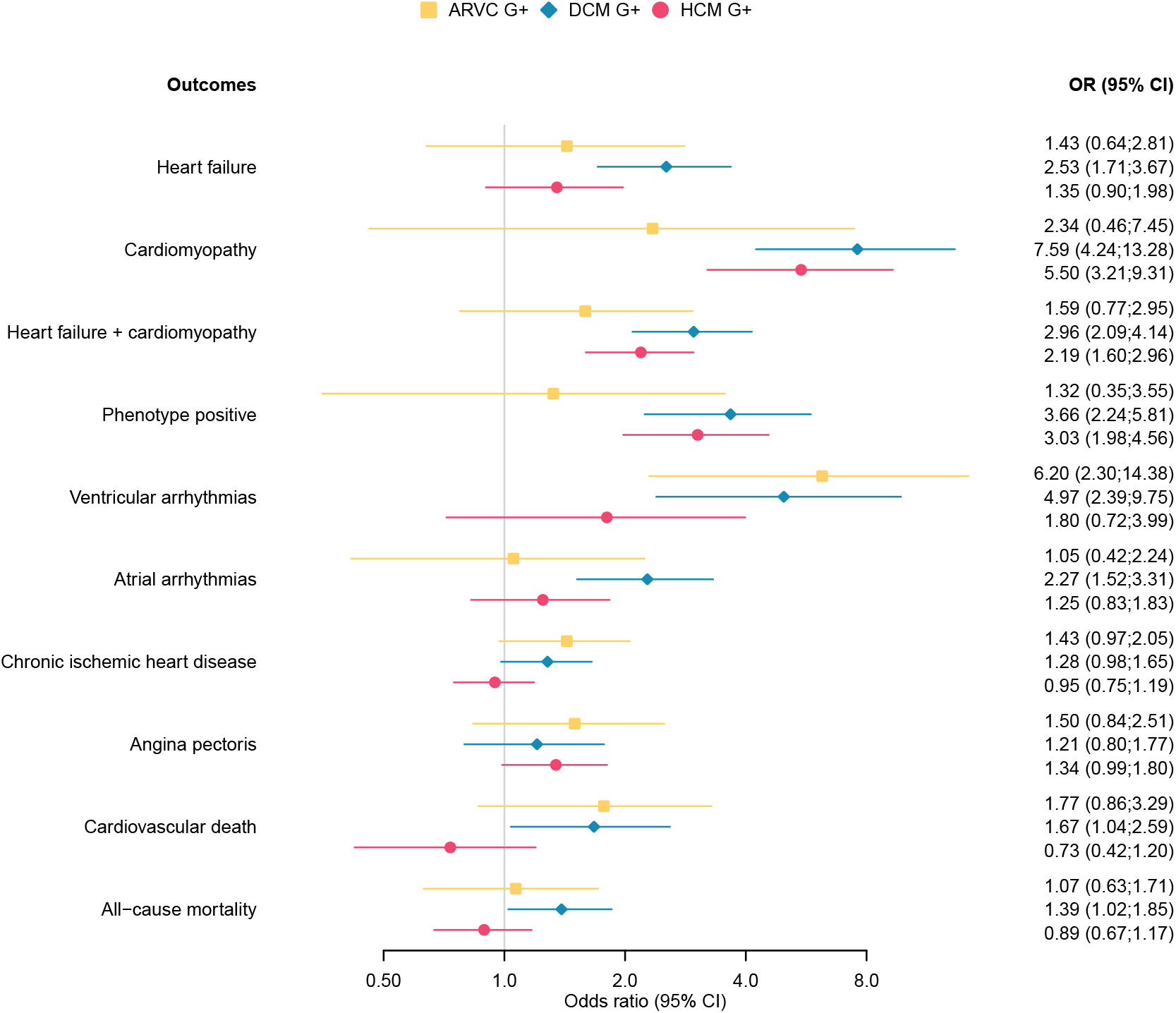
Forest plot cardiac outcomes stratified per inherited cardiomyopathy. Odds ratios and 95% confidence interval are given for the associations between cardiac outcomes and ARVC, DCM, or HCM pathogenic variant carriers. *Abbreviations: ARVC= arrhythmogenic right ventricular cardiomyopathy; DCM= dilated cardiomyopathy; G+= pathogenic variant carrier; HCM= hypertrophic cardiomyopathy*.

Ventricular arrhythmias occurred more often in G+ compared to G-, reaching statistical significance for ARVC (OR 6.20 [95% CI 2.30;14.38], p=3.3×10^−4^) and DCM (OR 4.97 [95% CI 2.39;9.75], p=1.9×10^−5^). Atrial arrhythmias were more prevalent among DCM G+ (OR 2.27 [95% CI 1.52;3.31], p=8.2×10^−5^). Finally, all-cause mortality (OR 1.39 [95% CI 1.02;1.85], p=0.032) and death due to a cardiovascular cause were more prevalent in DCM G+ (OR 1.67 [95% CI 1.04;2.59], p=0.030) but did not reach statistical significance for ARVC G+ and HCM G+ (p≥0.100). **Figure I** depicts the overlap in cardiomyopathy, heart failure, ventricular arrhythmia and ischemic heart disease diagnosis. **Figure II** depicts the forest plots when excluding the more prevalent *TNNT2* and *MYBPC3* variants in HCM G+ individuals and **Figure III** shows the association between different outcomes stratified by genes for each cardiomyopathy.

### Deep phenotyping of undiagnosed pathogenic variant carriers

Next, we set out to study early signs of disease in G+ without a cardiomyopathy/heart failure diagnosis (P-) using registered ICD-10 codes, self-reported cardiac symptoms, and abnormal ECG and CMR values.

#### Diagnosis and symptoms

Ventricular arrhythmias were more prevalent in ARVC G+P- (OR 5.85 [95% CI 1.98;14.40], p=0.001) and DCM G+P- (OR 3.43 [95% CI 1.35;7.68], p=0.005) but not in HCM G+P- (OR 1.01 [95% CI 0.26;2.86], p=1.000) compared to G-P- controls. Also, atrial arrhythmias (OR 2.12 [95% CI 1.36;3.19], p=7.9×10^−4^) were more frequent in DCM G+P- compared to G-P- controls. Finally, angina pectoris occurred more often in HCM G+P- (OR 1.38 [95% CI 1.01;1.85], p=0.038), but not in ARVC G+P- and DCM G+P- (p≥0.117; **Table VII**).

#### Electrocardiography

In total, 231 out of 2,207 G+P- and 1,058 out of 9,885 G-P- had various ECG variables available. None of these ECG variables differed significantly between all undiagnosed G+ and control individuals (**Table VIII**).

#### Cardiac magnetic resonance imaging

CMR data was available in 225 G+P- of the 2,207 unique G+P- individuals: n=33 for ARVC G+P-, n=87 for DCM G+P- and n=130 for HCM G+P-) and 986 of the 9,885 G-P- controls. As shown in **Table IX**, G+P- and G-P- individuals with CMR data available were mostly comparable. Only smoking was more prevalent among DCM G+P- compared to G-P- controls (OR 1.59 [95%CI 1.00; 2.53], p=0.041). Outliers were observed in G-P- controls: 4 individuals with a median age of 64 [60-67] years had a left ventricular ejection fraction (LVEF) below 40% and 3 of them were diagnosed with hypertension and 2 with acute myocardial infarction in the past. In addition, 2 individuals with an age of 42 and 52 years old had an RVEF below 40%. They did not suffer from hypertension and did not have any cardiac diagnosis.

As shown in **Figure 5** and **Table VIII**, all RV (p≥0.546) and LV (p≥0.052) functional and structural parameters in ARVC G+P- were comparable to G-P- controls. Three ARVC G+P- individuals had an RVEDV corrected for body surface area (RVEDVi) between 100-110 mL/m^2^ for males or 90-100 mL/m^2^ for females, meeting the minor CMR task force criteria (TFC) if wall motion abnormalities were present, and two ARVC G+P- individuals had an RVEDVi larger than 110 mL/m^2^ for males or 100 mL/m^2^ for females, meeting the major CMR TFC^10^. In addition, ARVC G+P- had reduced inferior and posterolateral wall thickness compared to controls (p≤0.035).

**Figure 5:**
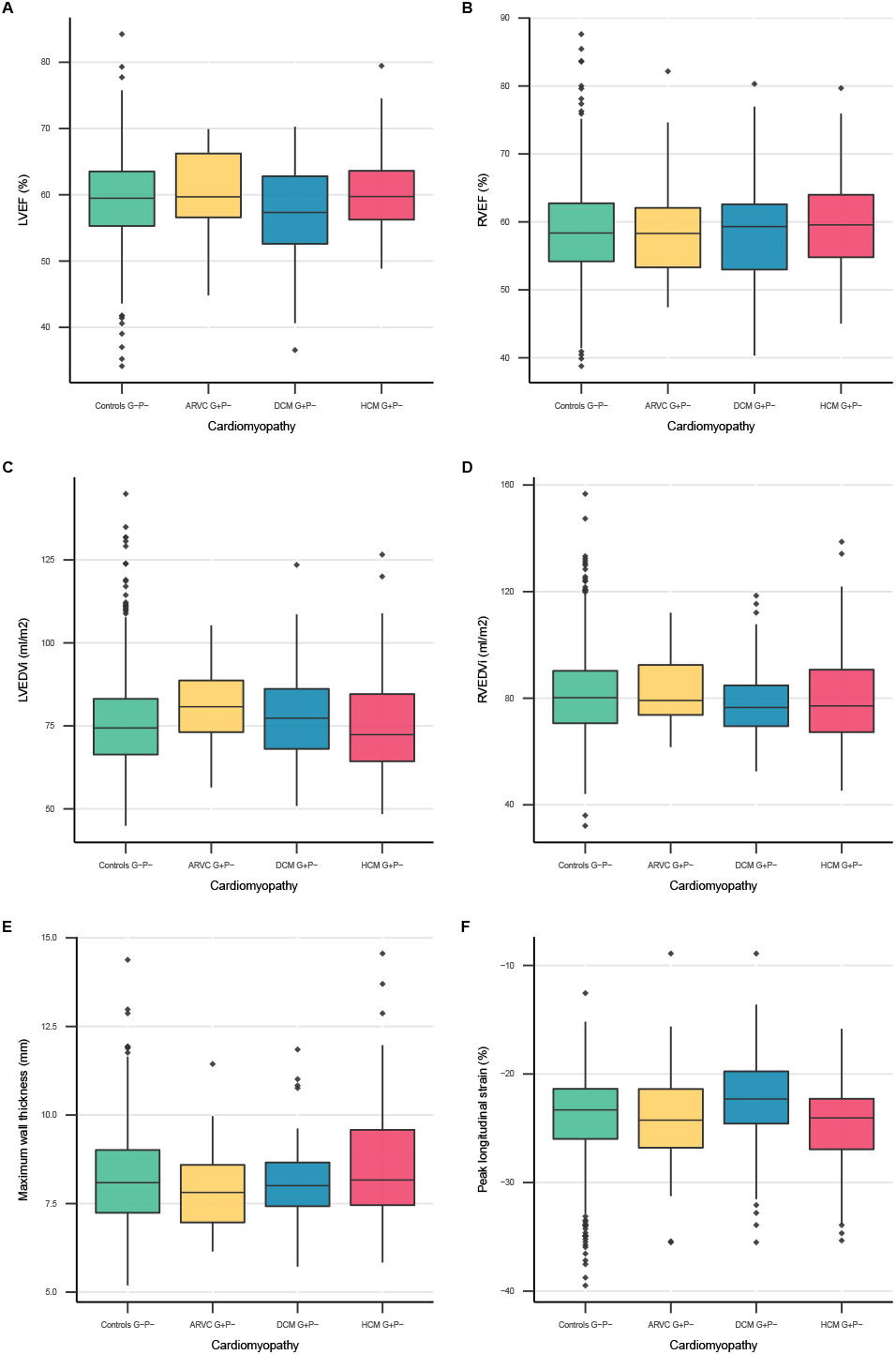
CMR parameters stratified per inherited cardiomyopathy. Boxplots of CMR parameters showing the summary statistics of CMR parameters stratified by controls and individuals with a pathogenic variant associated with ARVC, DCM, or HCM. Displayed summary statistics include the median, first and third quartile (lower and upper box edges), and the whiskers represent values within 1.5 times the interquartile range from the box edges. *Abbreviations: ARVC= arrhythmogenic right ventricular cardiomyopathy; DCM= dilated cardiomyopathy; EDVi= body surface area corrected end-diastolic volume; EF= ejection fraction; G+= pathogenic variant carrier; HCM= hypertrophic cardiomyopathy; LV= left ventricular; RV= right ventricular*.

Overall, DCM G+P- and G-P- controls had comparable RV functional and structural measures (p≥0.048). However, DCM G+P- had lower LVEF (57.3% [52.6, 62.8] vs. 59.5% [55.3, 63.5] vs, p=0.009) and less negative LV peak longitudinal strain (−22.3% [-24.6, -19.86] vs. -23.3% [-26.0, -21.4], p=0.009). Although LVEDVi was not significantly increased in DCM G+P-, the LVEDV/RVEDV ratio (0.9 [0.9, 1.0] vs 1.0 [0.9, 1.1], p=8.2×10^−4^) and LVESVi (30.0 ml/m^2^ [25.1, 35.7] vs 31.7 ml/m^2^ [26.2, 39.8], p=0.032) were increased. Six individuals had an LVEF below 45%, but none of the individuals met the Henry criteria for DCM (LVEF below 45% and LVEDVi two times the normal SD)^11^.

For HCM G+P-, most RV and LV functional and dimension parameters were comparable to G-P- controls (p≥0.051). Only RVEF was higher than controls (58.4% [54.2, 62.7] vs 59.6% [54.8, 64.0], p=0.025). Importantly, wall thickness was not significantly different between HCM G+P- without a cardiomyopathy/heart failure diagnosis and G-P-(p≥0.160). None of the G+P- individuals met HCM criteria^12^ of ≥15 mm wall thickness, but two individuals met the criteria for limited hypertrophy (13-15mm) in the presence of a positive genetic test^12^.

**Figure IV** shows a summary of all the differences tested.

#### Exclusion of the more prevalent *TNNT2* and *MYBPC3* variants

When excluding the more prevalent *TNNT2* and *MYBPC3* variants in HCM G+P- individuals, the occurrence of ventricular arrhythmias (OR 1.72 [95% CI 0.44;4.89], p=0.306) and atrial arrhythmias (OR 1.43 [95% CI 0.84;2.32], p=0.156) was comparable to G-P- controls. However, the maximum wall thickness (8.47mm [7.59, 9.94] vs. 8.09mm [7.24, 9.01], p=0.008) and basal anterior wall thickness (7.93mm [6.97, 9.11] vs. 7.65mm [6.81, 8.49], p =0.029) were significantly increased in HCM G+P- compared to controls (**Table VIII**). Two individuals had a maximum wall thickness between 13-15mm.

## Discussion

In this study we leveraged the largest European population database including WES and phenotype data to evaluate the prevalence and penetrance of previously reported pathogenic and likely pathogenic variants associated with ARVC, DCM and HCM. Our study has several interesting findings. First, we found a prevalence of 1:578, 1:251, and 1:149 for variants previously associated with ARVC, DCM and HCM respectively. Second, 1.2% of ARVC G+, 3.1% of DCM G+ and 2.6% of HCM G+ were diagnosed with a cardiomyopathy or heart failure without previous chronic ischemic heart disease. Finally, 3.2% of the undiagnosed ARVC G+, 1.8% of the undiagnosed DCM G+, 0.5% of the undiagnosed HCM G+ reported ventricular arrhythmias or had CMR abnormalities. These results confirm the low disease penetrance in G+ in the general population.

### Prevalence of pathogenic and likely pathogenic variant carriers in the general population

Since rare genetic variants are the major cause of inherited cardiomyopathies, a large dataset is needed to accurately identify the population prevalence of these variants. Prevalence of pathogenic variants in populations has been the focus of several previous studies^4,13-15^, however they were mostly limited by the number of included individuals. At time of analysis, we had access to an unprecedented number of 200,643 individuals.

Previously reported prevalence of ARVC G+ in the general population ranges between 1:143 to 1:1,706^13-15^. This variability is likely to be explained by heterogeneity in study populations and definitions of variant pathogenicity. For example, many previous studies did not include all eight curated genes with strong or moderate disease-gene association but also marked other genes (e.g. *TGFB3*) with only limited evidence as associated with ARVC^14,15^. In addition, we included both missense and LoF variants whereas prior studies only included LoF variants.

For DCM, little is known about the prevalence of DCM causing variants in the general population. Studies focusing on truncating *TTN* variants in the general population found a prevalence ranging between 1:33 and 1:526^16,17^. This wide range can partly be explained by the used definition of pathogenicity. Also, disease causing truncating *TTN* variants associated with DCM are known to be highly enriched in the A band. However, recently, truncating variants in the distal I-band region have also been implicated in DCM^18^. When solely focussing on *TTN* variants, we found a prevalence of only 1:735. This differs from the previous studies, probably because not all *TTN* variants are reported as pathogenic or likely pathogenic in ClinVar and VKGL. Including all curated DCM-associated genes, we report a prevalence of 1:251.

For HCM, we found a prevalence ranging between 1:250 and 1:149 individuals carrying a pathogenic or likely pathogenic variant, which approaches previous estimates of 1:164^19^. In a recent study, including the UKB population, a prevalence of 1:407 was reported^20^. They included 8 sarcomere-encoding genes described to be associated with HCM (*ACTC1, MYBPC3, MYH7, MYL2, MYL3, TNNI3, TNNT2* and *TPM1*) and variants that were described as pathogenic or likely pathogenic in ClinVar or annotated as pathogenic or likely pathogenic according to ACMG criteria and filtered variants for an allele frequency of 0.00004. We included additional genes (*CSRP3, JPH2* and *TNNC1*) and pathogenic and likely pathogenic variants from the VKGL database and filtered for a minor allele frequency of 0.001. Especially the latter is a driving force behind the higher prevalence in this study. When also using a minor allele frequency (MAF) <0.00004, the prevalence of our study would be 1:475, approaching the prevalence reported by de Marvao *et al*.^20^.

### Disease expression of pathogenic and likely pathogenic variants in the general population

Most information on disease penetrance in ARVC, DCM or HCM G+ is based on observations in G+ relatives of cardiomyopathy patients. Previous studies have shown that 37% of ARVC G+ relatives^21^ and up to 50% of HCM G+ relatives with sarcomeric variants^22^ show disease expression during follow-up. Our findings suggest that disease penetrance in the general population is much lower. We found that 1.2% of ARVC G+, 3.1% of DCM G+ and 2.6% of HCM G+ in the UKB were diagnosed with a cardiomyopathy or heart failure, in the absence of chronic ischemic heart disease. Our additional analysis of ventricular function and ECG in undiagnosed G+ subjects also suggests a low disease penetrance. We found significantly worse LVEF and strain parameters in DCM G+P- compared to controls, however none met the diagnostic Henry criteria (LVEF below 45% and LVEDVi two times the normal SD)^11^. Although CMR data was only available in a subgroup of undiagnosed G+ patients, these findings make it unlikely that the low penetrance found in our study arises from missed diagnoses or covert disease in the G+ cohort. Furthermore, none of the G+P- individuals met HCM criteria^12^ of ≥15 mm wall thickness, two individuals did meet the criteria for limited hypertrophy (13-15mm) in the presence of a positive genetic test^12^. Interestingly, maximum wall thickness in de Marvao *et al*.^20^ was higher compared to ours. Although this can partly be explained by the inclusion of P+ by Marvao *et al*., this may also be explained by differences in wall thickness calculation method. While Marvao *et al*. uses the absolute largest wall thickness value at a single point, we have used the AHA segment with the largest wall thickness (which is an average of all the single points within one AHA segment to reduce random outliers). In ARVC and DCM G+P- we found a low, but significantly higher prevalence of ventricular arrhythmias compared to controls (1.7% vs. 0.3% (OR 5.85 [95% CI 1.98;14.40]) and 1.0% vs. 0.3% (OR 3.43 [95% CI 1.35;7.68)) respectively). In ARVC, electrical abnormalities are known to precede structural abnormalities^23^. Therefore, these findings may suggest early disease penetrance in a small subset of undiagnosed G+ individuals. The discrepancy between the high disease penetrance found in G+ family members and the low penetrance in the G+ general population points towards the interaction of possible other (unidentified) genetic and environmental factors leading to this variation. The median age of our study population was 57 [49-63] years, however inherited cardiomyopathies are generally diagnosed at a younger age. For ARVC, Groeneweg *et al*. showed, in a cohort of 439 index-patients, that the mean age of first disease presentation is 36±14 years. Most of these patients presented with symptoms (95%), of whom 11% with sudden cardiac arrest^21^. Likewise in DCM, the mean age of presentation is mostly between 30-50 years^24^. Lastly, in HCM a mean age at presentation of 49±16 years was shown in a cohort of 4893 patients by Lorenzini *et al*.^*25*^. Interestingly, although mortality rates were low, young HCM patients showed a worse prognosis compared to their healthy peers, with 80% of mortality being caused by sudden cardiac death^25^. Therefore, it should be taken into account that younger patients with disease expression are likely underrepresented in our study. This is not only due to higher mortality and morbidity in especially ARVC and HCM, but also because individuals with a diagnosed cardiomyopathy may be less likely to participate in a large-scale biobank study such as the UKB.

Interestingly, the South Asian *MYBPC3* and the *TNNT2* variant, showed a relatively high prevalence in our cohort. In total, 19% of HCM G+ was Asian and most of these individuals carried the c.3628-41_3628-17del variant in the *MYBPC3* gene. Although this variant is indicated as likely pathogenic in ClinVar, a previous study suggests that this variant may be reclassified as benign^26^. In our study, none of these variant carriers were diagnosed with HCM. Four were diagnosed with heart failure of whom one was diagnosed with DCM. This suggests that this variant is associated with heart failure in the setting of multiple forms of cardiomyopathy, and not simply HCM^26^. Secondly, the c.862C>T p.Arg288Cys variant in *TNNT2* was previously found in HCM individuals but is often observed in patients with a mild phenotype or in combination with other variants. These observations suggest that this variant might not be a monogenic cause of severe HCM but acts in concert with other variants^27^. Interestingly, when excluding these variants from our G+P- population, a significantly higher wall thickness is measured compared to controls. These two examples emphasize that when pathogenic or likely pathogenic variants are identified as a secondary finding, other factors, such as the specific variant and the family history are crucial for follow-up decisions.

We also assessed gene-specific associations with the cardiovascular outcomes. *PKP2* variant carriers showed a stronger association with ventricular arrhythmias (OR 11.90 [95% CI 4.38; 27.86], p = 6.4×10^−6^) compared to heart failure (OR 1.50 [95% CI 0.48; 3.64], p=0.395). This is in concordance with a previous study showing sustained ventricular arrhythmias to be the first clinical presentation in 61% of ARVC patients^21^. During follow-up, sustained arrhythmias occurred in 72% of ARVC patients, highlighting sustained arrhythmias as the most important ARVC disease manifestation. On the contrary, symptomatic heart failure was seen in 13% of ARVC patients^21^. In DCM G+, ventricular arrhythmias were significantly more present compared to G- controls, especially in *TTN* (OR 4.49 [95% CI 1.15; 12.76], p=0.016), *DES* (OR 12.80 [95% CI 1.45; 52.55], p=0.013) and *LMNA* (OR 15.04 [95% CI 1.69; 62.32], p=0.009) variant carriers. A recent meta-analysis assessing predictors for sustained ventricular arrhythmias, showed *PLN* and *LMNA* to be associated with arrhythmogenic outcome^28^. Although we did not have enough power to study *PLN* G+, *LMNA* G+ did show significantly more ventricular arrhythmias compared to G- controls. Furthermore, *BAG3* variant carriers have been associated with significant risk of progressive heart failure^29^. In our study, *BAG3* variant carriers were significantly more often diagnosed with a cardiomyopathy (OR 41.18 [95% CI 4.36; 192.17], p=0.002). Even though more heart failure cases were seen compared to G- controls, this did not reach statistical significance (**Table X**).

Interestingly, self-reported health-related quality of life and psychological well-being of 89 asymptomatic HCM G+ were previously evaluated in a Dutch cohort and found to be at least similar to the general population, which suggests that reporting incidental findings will not harm psychological well-being of G+^30^. However, frequent cardiological examination of G+ and family members turning out to be carriers after cascade screening will put a burden on health care and societal costs^31^. Genetic screening and cardiological examination are necessary in family members of genetic cardiomyopathy patients since disease expression in family members is considerable. Disease expression in the general population on the other hand is low. Therefore, in case of an incidental finding, multiple factors like family history, presence of symptoms, electrical and/or structural abnormalities and gene and variant type should inform follow-up decisions. Further studies on the genotype-phenotype associations and disease penetrance will aid in facilitating these decisions.

### Limitations

Several variants are associated with more than one cardiomyopathy. This is mainly due to phenotypic heterogeneity but may also be partly explained by misdiagnosis. Information is submitted to ClinVar by laboratories, not by clinicians. Phenotype description might therefore be less reliable. To avoid selection bias, we included variants associated with multiple cardiomyopathies in both cardiomyopathy categories, possibly leading to increased prevalence estimates. Although the prevalence of cardiomyopathy variants is slightly affected by including or excluding overlapping variants, this did not substantially affect the results and conclusions (**Table X**). Future studies should focus on reaching consensus on variant-phenotype associations for the variants described in multiple cardiomyopathies to avoid variation in prevalence caused by the use of different definitions. Despite recent efforts to harmonize knowledge on genes associated with inherited cardiomyopathies^5-7^, and guidelines for variant classification^31^, the adjudication of the clinical significance of single variants can still differ between diagnostic laboratories^31^, which has led to interpretation differences and difficulties to compare results among studies using different criteria. This highlights the importance of a single set of criteria to ascertain clinical significance of a single variant. Furthermore, not all pathogenic or likely pathogenic variants are reported in these databases, especially family-specific variants and pathogenic variants in non-Caucasian populations are underreported.

Lastly, G+P- and G-P- individuals with CMR data available were age, sex and ethnicity matched and comparable in the presence of cardiovascular risk factors and diseases. Interestingly, outliers in CMR values were also present in G-P- controls, which could be partly explained by the presence of past myocardial infarctions. Therefore, differences in cardiac function and structure between G+P+ and G-P- could be underestimated.

## Conclusion

In a cohort of 200,643 individuals with WES and phenotype data we identified a prevalence of pathogenic variants associated with ARVC, DCM and HCM of 1:578, 1:251 and 1:149 respectively. Among the identified G+ individuals, cardiomyopathy, heart failure and ventricular arrhythmias were more common compared to G-. However, overall disease penetrance was low (1.2-3.1%). Therefore, in case of incidental findings, decisions on application of cascade screening and frequency of cardiological examination should be based on multiple factors besides variant and gene type, such as family history and disease expression.

## Data Availability

All data produced are available online at https://github.com/CirculatoryHealth/Inherited-cardiomyopathies.

## Non-standard Abbreviations and Acronyms

ACMG: American College of Medical Genetics and Genomics
ACTC1: Actin Alpha Cardiac Muscle 1
ACTN1: Alpha-actinin 2
AHA: American Heart Association
ARVC: Arrhythmogenic right ventricular cardiomyopathy
BAG3: BAG Cochaperone 3
BMI: Body mass index
CI: Confidence interval
ClinGen: Clinical Genome Resource
CMR: Cardiac magnetic resonance imaging
CSRP3: Cysteine And Glycine Rich Protein 3
DCM: Dilated cardiomyopathy
DES: Desmin
DSC2: Desmocollin 2
DSG2: Desmoglein 2
DSP: Desmoplakin
ECG: Electrocardiography
EDM: End diastolic mass
EDV: End diastolic volume
EF: Ejection fraction
ESV: End systolic volume
FLNC: Filamin-C
G+: Genotype positive (variant carriers)
G-: Genotype negative (controls)
i: Indexed, corrected for body surface area
JPH2: Junctophilin 2
JUP: Junction Plakoglobin
HCM: Hypertrophic cardiomyopathy
LMNA: Lamin A/C
LoF: Loss of function
LV: Left ventricular
MAF: Minor allele frequency
MET: Metabolic equivalent of task minutes
MVR: Mass to EDV ratio
MYBPC3: Myosin Binding Protein C3
MYH7: Myosin Heavy Chain 7
MYL2: Myosin Light Chain 2
MYL3: Myosin Light Chain 3
NEXN: Nexilin F-Actin Binding Protein
NGS: Next generation sequencing
OR: Odds ratio
P+: Phenotype positive
P-: Phenotype negative
PKP2: Plakophilin 2
PLN: Phospholamban
RBM20: RNA Binding Motif Protein 20
RV: Right ventricular
SCN5A: Sodium Voltage-Gated Channel Alpha Subunit 5
SD: Standard deviation
SV: Stroke volume
TFC: Task force criteria
TMEM43: Transmembrane Protein 43
TNNC1: Troponin C1, Slow Skeletal And Cardiac Type
TNNI3: Troponin I3, Cardiac Type
TNNT2: Troponin T2, Cardiac Type
TPM1: Tropomyosin 1
TTN: Titin
UKB: UK Biobank
VKGL: Vereniging Klinische Genetische Laboratoriumdiagnostiek
VCL: Vinculin
WES: Whole exome sequencing

## Acknowledgements

This research has been conducted using the UK Biobank Resource under Application Number 24711.

## Supplemental Material

Supplemental Methods

Tables I-XI

Figures I-IV

References^32-38^

