## Supplemental Material for "Prevalence and disease expression of pathogenic and likely pathogenic variants associated with inherited cardiomyopathies in the general population"

**Content**

Supplemental Methods……………………………………………………………………………………………………..2

Supplemental Tables………………………………………………………………………………………………………..5

Supplemental Figures……………………………………………………………………………………………………….30

**Supplemental Methods**

*Genetic variants in the study population*

We identified carriers of a pathogenic or likely pathogenic variant associated with ARVC, DCM or HCM in individuals from the UKB who underwent whole exome sequencing (WES, n=200,643 at time of analysis). For each inherited cardiomyopathy we selected curated genes classified to have definite, strong or moderate evidence of pathogenicity as defined by the standardized evidence-based framework of Clinical Genome Resource (ClinGen)[32] and curated by James *et al*.[5] for ARVC, Jordan *et al.*[6] for DCM and Ingles *et al*.[7] for HCM. For ARVC we included *DES, DSC2, DSG2, DSP, JUP, PKP2, PLN* and *TMEM43;* for DCM we included *ACTC1, ACTN2, BAG3, DES, DSP, FLNC, JPH2, LMNA, MYH7, NEXN, PLN, RBM20, SCN5A, TNNC1, TNNI3, TNNT2, TPM1, TTN* and *VCL;* and for HCM we included *ACTC1, CSRP3, JPH2, MYBPC3, MYH7, MYL2, MYL3, TNNC1, TNNI3, TNNT2* and *TPM1* (**Figure 1** and **Table I**). Some genetic variants are associated with two cardiomyopathies (**Table IV**). Individuals carrying these variants were included in the G+ groups of both cardiomyopathies. Next, likely pathogenic and pathogenic variants in these genes were identified using the ClinVar NCBI-NIH database[8] and the Dutch Society for Clinical Genetic Laboratory Diagnostics (Vereniging Klinische Genetische Laboratoriumdiagnostiek, VKGL) database. Laboratories submitting information to these databases use the criteria for variant classification as defined by the American College of Medical Genetics and Genomics and the Association for Molecular Pathology (ACMG-AMP)[33]. An elaborate overview of the ClinVar and VKGL search criteria is given in **Figure 2**. In short, ClinVar was queried using the disease name(s) and filtered for pathogenic and likely pathogenic variants in the curated genes. For variants mentioned in the VKGL database, which does not specify disease associations, association with one of the cardiomyopathies was confirmed in ClinVar. The minor allele frequency

(MAF) cut-off was defined at 0.001, which is the recommended cut-off for including rare variants[34] and still include any potentially at-risk variant carriers. Variants were classified as missense or loss of function (LoF), with LoF being defined as frameshift, stop gain, start lost and canonical splice site variants.

We matched G+ individuals in a 1:4 ratio to UKB individuals without a pathogenic or likely pathogenic variant associated with one of the cardiomyopathies (G-). Matching of this G-control group was based on age, sex, ethnicity and presence of cardiac magnetic resonance imaging (CMR) measurements. Controls are referred to as G- throughout this study.

*Data extraction UKB*

Disease definitions

An elaborate overview of the disease definitions used in this study is available in **Table II**. In short, individuals were defined to be phenotype positive (P+) if they had an ICD-10 or self-reporting code for cardiomyopathy, DCM, HCM or heart failure, without a diagnosis of chronic ischemic heart disease. No ICD-10 or self-reporting code was available for ARVC in the UKB.

CMR and ECG data analysis

We investigated disease expression on cardiac magnetic resonance imaging (CMR, n = 225 unique individuals) and electrocardiography (ECG, n = 231 unique individuals) of G+P- individuals. The full CMR protocol of the UKB has been described in detail[35]. In short, all CMR examinations were performed on a 1.5 Tesla scanner (Magnetom Aera, Syngo Platform VD13A, Siemens Healthcare, Erlangen, Germany). We used a previously developed and validated deep-learning methodology (AI-CMR$^{QC}$) to extract left (LV) and right ventricular (RV) CMR measurements[36]. In short, cine images of short-axis and 2- and 4-chamber long-axis views were used to automatically calculate LV and RV functional measures (ejection fraction [EF], stroke volume [SV]) and structural measures (end diastolic volume [EDV], end-systolic volume [ESV], LV end diastolic mass [EDM], LV mass to EDV ratio [LVMVR] and LV maximal and regional [16 segments model according the American Heart Association[37]] wall thickness).

The electrocardiography (ECG) variables P duration, P axis, PQ interval, QRS duration, R axis, QTc interval and T axis were extracted from the UKB for G+P- individuals.

*Statistical analysis*

Statistical analysis was performed using R version 4.0.2[38]. Continuous values are presented as median [interquartile range] and for comparisons of two groups, Mann-Whitney-U test was used. Categorical data was displayed as absolute frequency (n) and percentages (%) and Fisher's exact test was used to test for differences. The strength of the association between cardiac outcomes and G+ ARVC, DCM and HCM was calculated by the odds ratio with 95% confidence intervals. The latter was also performed stratifying by genes.

A number of included variants are associated with more than one cardiomyopathy. To investigate the effect of the inclusion of individuals in more than one cardiomyopathy, we removed overlapping variants as described in **Table IV** and calculated the odds ratios of the cardiovascular risk factors and diagnoses for these G+ compared to G-.

A p-value of less than 0.05 was considered significant.

| **Supplementary Table I: Included curated genes per cardiomyopathy** | | | |
|---|---|---|---|
| **Gene*** | **ARVC** | **DCM** | **HCM** |
| *ACTC1* | | Moderate | Definitive |
| *ACTN2* | | Moderate | |
| *BAG3* | | Definitive | |
| *CSRP3* | | | Moderate |
| *DES* | Moderate | Definitive | |
| *DSC2* | Definitive | | |
| *DSG2* | Definitive | | |
| *DSP* | Definitive | Strong | |
| *FLNC* | | Definitive | |
| *JPH2* | | Moderate | Moderate |
| *JUP* | Definitive | | |
| *LMNA* | | Definitive | |
| *MYBPC3* | | | Definitive |
| *MYH7* | | Definitive | Definitive |
| *MYL2* | | | Definitive |
| *MYL3* | | | Definitive |
| *NEXN* | | Moderate | |
| *PKP2* | Definitive | | |
| *PLN* | Moderate | Definitive | |
| *RBM20* | | Definitive | |
| *SCN5A* | | Definitive | |
| *TMEM43* | Definitive | | |
| *TNNC1* | | Definitive | Moderate |
| *TNNI3* | | Moderate | Definitive |
| *TNNT2* | | Definitive | Definitive |
| *TPM1* | | Moderate | Definitive |
| *TTN* | | Definitive | |
| *VCL* | | Moderate | |

* ARVC genes are curated by ref 5, DCM genes by ref 6 and HCM genes by ref 7.
Pathogenicity is classified as moderate, strong and definitive.

Abbreviations:
*ACTC1* : Actin Alpha Cardiac Muscle 1; *ACTN2* : Alpha-actinin 2;
ARVC: Arrhythmogenic right ventricular cardiomyopathy; *BAG3* : BAG Cochaperone 3;
*CSRP3* : Cysteine And Glycine Rich Protein 3; DCM: Dilated cardiomyopathy; *DES* : Desmin;
*DSC2* : Desmocollin 2; *DSG2* : Desmoglein 2; *DSP* : Desmoplakin; *FLNC* : Filamin-C;
HCM: Hypertrophic cardiomyopathy; *JPH2* : Junctophilin 2; *JUP* : Junction Plakoglobin;
*LMNA* : Lamin A/C; *MYBPC3* : Myosin Binding Protein C3; *MYH7* : Myosin Heavy Chain 7;
*MYL2* : Myosin Light Chain 2; *MYL3* : Myosin Light Chain 3; *NEXN* : Nexilin F-Actin Binding Protein;
*PKP2* : Plakophilin 2; *PLN* : phospholamban; *RBM20* : RNA Binding Motif Protein 20;
*SCN5A* : Sodium Voltage-Gated Channel Alpha Subunit 5; *TMEM43* :Transmembrane Protein 43;
*TNNC1* : Troponin C1, Slow Skeletal And Cardiac Type; *TNNI3* : Troponin I3, Cardiac Type;
*TNNT2* : Troponin T2, Cardiac Type; *TPM1* : Tropomyosin 1; *TTN* : Titin; *VCL* : Vinculin.

**Supplementary Table II: Disease definitions**

| Phenotype | Field names | Values (ICD or other coding) |
|---|---|---|
| **Diabetes** | Diagnoses ICD10<br>Underlying (primary) cause of death: ICD10<br>Contributory (secondary) causes of death: ICD10<br>External causes ICD10<br>Diagnoses main ICD10<br>Diagnoses secondary ICD10 | E10*; E11*; E12*; E13*; E14* |
| | Non-cancer illness code self-reported | 1220; 1222; 1223 |
| | Diabetes diagnosed by doctor | 1 |
| | Medication for cholesterol blood pressure or diabetes(, or take exogenous hormones) | 3 |
| **Hypertension** | Diagnoses ICD10<br>Underlying (primary) cause of death: ICD10<br>Contributory (secondary) causes of death: ICD10<br>External causes ICD10<br>Diagnoses main ICD10<br>Diagnoses secondary ICD10 | I10; I15* |
| | Non-cancer illness code self-reported | 1065; 1072 |
| | Medication for cholesterol blood pressure or diabetes(, or take exogenous hormones) | 2 |
| **Hypercholesterolaemia** | Diagnoses ICD10<br>Underlying (primary) cause of death: ICD10<br>Contributory (secondary) causes of death: ICD10<br>External causes ICD10<br>Diagnoses main ICD10<br>Diagnoses secondary ICD10 | E780 |
| | Non-cancer illness code self-reported | 1473 |
| | Medication for cholesterol blood pressure or diabetes(, or take exogenous hormones) | 1 |
| **Ever smoked** | Smoking status | 1; 2 |
| **Family heart disease** | Illnesses of father<br>Illnesses of mother<br>Illnesses of siblings | 1 |
| **Cardiac problem** | Non-cancer illness code self-reported | 1066 |
| **Heart failure** | Diagnoses ICD10<br>Underlying (primary) cause of death: ICD10<br>Contributory (secondary) causes of death: ICD10<br>External causes ICD10<br>Diagnoses main ICD10<br>Diagnoses secondary ICD10 | I110; I130; I132; I50* |
| | Non-cancer illness code self-reported | 1076 |
| **Cardiomyopathy** | Diagnoses ICD10<br>Underlying (primary) cause of death: ICD10<br>Contributory (secondary) causes of death: ICD10<br>External causes ICD10<br>Diagnoses main ICD10<br>Diagnoses secondary ICD10 | I42* |
| | Non-cancer illness code self-reported | 1079 |
| **Dilated cardiomyopathy** | Diagnoses ICD10<br>Underlying (primary) cause of death: ICD10<br>Contributory (secondary) causes of death: ICD10<br>External causes ICD10<br>Diagnoses main ICD10<br>Diagnoses secondary ICD10 | I420 |
| **Hypertrophic cardiomyopathy** | Diagnoses ICD10<br>Underlying (primary) cause of death: ICD10<br>Contributory (secondary) causes of death: ICD10<br>External causes ICD10<br>Diagnoses main ICD10<br>Diagnoses secondary ICD10 | I421; I422 |
| | Non-cancer illness code self-reported | 1588 |
| **Ventricular arrhythmias** | Diagnoses ICD10<br>Underlying (primary) cause of death: ICD10<br>Contributory (secondary) causes of death: ICD10<br>External causes ICD10<br>Diagnoses main ICD10<br>Diagnoses secondary ICD10 | I470; I472; I490; I493 |
| **Atrial arrhythmias** | Diagnoses ICD10<br>Underlying (primary) cause of death: ICD10<br>Contributory (secondary) causes of death: ICD10<br>External causes ICD10<br>Diagnoses main ICD10<br>Diagnoses secondary ICD10 | I48*; I471; I491 |
| | Non-cancer illness code self-reported | 1471; 1483; 1487 |
| **Heart arrhythmia** | Non-cancer illness code self-reported | 1077 |
| **Chronic ischemic heart disease** | Diagnoses ICD10<br>Underlying (primary) cause of death: ICD10<br>Contributory (secondary) causes of death: ICD10<br>External causes ICD10<br>Diagnoses main ICD10<br>Diagnoses secondary ICD10 | I25* |

| Phenotype | Field names | Values (ICD or other coding) |
|---|---|---|
| **Acute myocardial infarction** | Diagnoses ICD10<br>Underlying (primary) cause of death: ICD10<br>Contributory (secondary) causes of death: ICD10<br>External causes ICD10<br>Diagnoses main ICD10<br>Diagnoses secondary ICD10 | I21*; I22*; I248; I249 |
| | Non-cancer illness code self-reported | 1075 |
| **Cardiac arrest** | Diagnoses ICD10<br>Underlying (primary) cause of death: ICD10<br>Contributory (secondary) causes of death: ICD10<br>External causes ICD10<br>Diagnoses main ICD10<br>Diagnoses secondary ICD10 | I46* |
| **Angina pectoris** | Non-cancer illness code self-reported | 1074 |
| **Conduction disorders** | Diagnoses ICD10<br>Underlying (primary) cause of death: ICD10<br>Contributory (secondary) causes of death: ICD10<br>External causes ICD10<br>Diagnoses main ICD10<br>Diagnoses secondary ICD10 | I44*; I45* |
| **Valvular disease** | Diagnoses ICD10<br>Underlying (primary) cause of death: ICD10<br>Contributory (secondary) causes of death: ICD10<br>External causes ICD10<br>Diagnoses main ICD10<br>Diagnoses secondary ICD10 | I34*; I35*; I36*; I37*; I05*; I06*; I07*; I08* |
| | Non-cancer illness code self-reported | 1078; 1488; 1489; 1490; 1584; 1585; 1586; 1587 |
| **Congenital heart disease** | Diagnoses ICD10<br>Underlying (primary) cause of death: ICD10<br>Contributory (secondary) causes of death: ICD10<br>External causes ICD10<br>Diagnoses main ICD10<br>Diagnoses secondary ICD10 | Q20*; Q21*; Q22*; Q23*; Q24*; Q25*; Q26* |
| **Pulmonary obstructive disease** | Diagnoses ICD10<br>Underlying (primary) cause of death: ICD10<br>Contributory (secondary) causes of death: ICD10<br>External causes ICD10<br>Diagnoses main ICD10<br>Diagnoses secondary ICD10 | J44*; J43*; I26*; I27* |
| | Non-cancer illness code self-reported | 1112; 1113; 1114; 1115; 1121 |
| **All-cause mortality** | Date of Death | Any non-missing value |
| **Cardiovascular death** | Underlying (primary) cause of death: ICD10<br>Contributory (secondary) causes of death: ICD10 | I* |

* indicates starting with previously indicated code.

**Supplementary Table III: Extensive baseline table**

| | Controls G- | | | | ARVC G+ | | | |
|---|---|---|---|---|---|---|---|---|
| | Overall | Diagnosed | Non-Diagnosed | Missing | Overall | Diagnosed | Non-Diagnosed | Missing |
| n (%) | 9,972 (100) | 87 (0.8) | 9,885 (99.2) | | 347 (100) | 4 (1.2) | 343 (98.8) | |
| **Sex = Female (%)** | 5,436 (54.5) | 35 (40.2) | 5,401 (54.6) | 0 | 187 (53.9) | 2 (50.0) | 185 (53.9) | 0 |
| **Age (median [IQR])** | 57.00 [49.00, 63.00] | 62.00 [56.00, 66.00] | 57.00 [49.00, 63.00] | 0 | 57.00 [50.00, 64.00] | 55.50 [52.25, 58.50] | 57.00 [50.00, 64.00] | 0 |
| **Ethnicity (%)** | | | | 1 | | | | 0.6 |
| Asian | 1,076 (10.9) | 5 (5.8) | 1071 (10.9) | | 10 (2.9) | 0 (0.0) | 10 (2.9) | |
| Black | 164 (1.7) | 3 (3.5) | 161 (1.6) | | 7 (2.0) | 1 (25.0) | 6 (1.8) | |
| Chinese | 56 (0.6) | 1 (1.2) | 55 (0.6) | | 11 (3.2) | 0 (0.0) | 11 (3.2) | |
| Mixed | 132 (1.3) | 2 (2.3) | 130 (1.3) | | 1 (0.3) | 0 (0.0) | 1 (0.3) | |
| Other | 160 (1.6) | 1 (1.2) | 159 (1.6) | | 5 (1.4) | 0 (0.0) | 5 (1.5) | |
| White | 8,288 (83.9) | 74 (86.0) | 8,214 (83.9) | | 311 (90.1) | 3 (75.0) | 308 (90.3) | |
| CARDIOVASCULAR RISK FACTORS | | | | | | | | |
| **BMI (median [IQR])** | 26.73 [24.15, 29.82] | 28.83 [25.33, 32.20] | 26.71 [24.15, 29.80] | 0.5 | 26.40 [24.03, 30.32] | 29.29 [27.22, 32.45] | 26.36 [24.02, 30.18] | 0.3 |
| **Diabetes (%)** | 914 (9.2) | 21 (24.1) | 893 (9.0) | 0 | 35 (10.1) | 1 (25.0) | 34 (9.9) | 0 |
| **Hypertension (%)** | 3,420 (34.3) | 60 (69.0) | 3,360 (34.0) | 0 | 116 (33.4) | 3 (75.0) | 113 (32.9) | 0 |
| **Mean systolic blood pressure (median [IQR])** | 135.50 [124.00, 149.00] | 144.00 [130.00, 155.50] | 135.50 [124.00, 149.00] | 0.1 | 136.00 [124.00, 147.50] | 128.25 [120.62, 134.38] | 136.25 [124.00, 147.50] | 0.3 |
| **Mean diastolic blood pressure (median [IQR])** | 82.00 [75.00, 88.50] | 84.00 [76.50, 91.50] | 81.50 [75.00, 88.50] | 0.1 | 81.50 [75.00, 87.88] | 82.00 [77.38, 85.25] | 81.50 [75.00, 88.00] | 0.3 |
| **Hypercholesterolaemia (%)** | 2,416 (24.2) | 34 (39.1) | 2,382 (24.1) | 0 | 86 (24.8) | 3 (75.0) | 83 (24.2) | 0 |
| **Total cholesterol (median [IQR])** | 5.61 [4.86, 6.38] | 5.23 [4.39, 6.10] | 5.61 [4.87, 6.38] | 4.3 | 5.51 [4.82, 6.40] | 4.97 [4.50, 5.78] | 5.51 [4.83, 6.40] | 4.6 |
| **HDL (median [IQR])** | 1.38 [1.16, 1.65] | 1.29 [1.13, 1.56] | 1.38 [1.16, 1.65] | 11.3 | 1.38 [1.16, 1.64] | 1.48 [1.30, 1.65] | 1.38 [1.16, 1.64] | 9.2 |
| **LDL (median [IQR])** | 3.50 [2.92, 4.09] | 3.22 [2.54, 3.81] | 3.50 [2.92, 4.09] | 4.6 | 3.39 [2.87, 4.04] | 3.06 [2.71, 3.58] | 3.40 [2.88, 4.04] | 4.6 |
| **Ever Smoked (%)** | 4,132 (41.4) | 50 (57.5) | 4,082 (41.3) | 0 | 161 (46.4) | 2 (50.0) | 159 (46.4) | 0 |
| **MET minutes per week for walking (median [IQR])** | 693.00 [297.00, 1,386.00] | 528.00 [255.75, 1,608.75] | 693.00 [297.00, 1,386.00] | 19.8 | 693.00 [297.00, 1,386.00] | 569.25 [247.50, 952.88] | 693.00 [297.00, 1,386.00] | 19.3 |
| **MET minutes per week for moderate activity (median [IQR])** | 480.00 [120.00, 1,200.00] | 480.00 [100.00, 1,680.00] | 480.00 [120.00, 1,200.00] | 19.8 | 480.00 [120.00, 1,200.00] | 900.00 [640.00, 1,140.00] | 480.00 [120.00, 1,200.00] | 19.3 |
| **MET minutes per week for vigorous activity (median [IQR])** | 240.00 [0.00, 960.00] | 0.00 [0.00, 820.00] | 240.00 [0.00, 960.00] | 19.8 | 320.00 [0.00, 960.00] | 1,060.00 [540.00, 1,410.00] | 320.00 [0.00, 960.00] | 19.3 |
| **Total MET minutes per week (median [IQR])** | 1,773.00 [810.00, 3,452.50] | 1,367.50 [478.50, 3,834.00] | 1,776.50 [810.00, 3,450.00] | 19.8 | 2,001.00 [922.50, 3,550.50] | 2,529.25 [1,517.50, 3,412.88 | 1,969.00 [922.50, 3,550.50] | 19.3 |
| **Family heart disease (%)** | 4,458 (44.7) | 34 (39.1) | 4,424 (44.8) | 0 | 179 (51.6) | 4 (100.0) | 175 (51.0) | 0 |
| CARDIAC DISEASES/OUTCOMES | | | | | | | | |
| **Cardiac problem (%)** | 41 (0.4) | 2 (2.3) | 39 (0.4) | 0 | 3 (0.9) | 0 (0.0) | 3 (0.9) | 0 |
| **Heart failure (%)** | 182 (1.8) | 74 (85.1) | 108 (1.1) | 0 | 9 (2.6) | 2 (50.0) | 7 (2.0) | 0 |
| **Cardiomyopathy (%)** | 37 (0.4) | 26 (29.9) | 11 (0.1) | 0 | 3 (0.9) | 3 (75.0) | 0 (0.0) | 0 |
| **Dilated cardiomyopathy (%)** | 14 (0.1) | 8 (9.2) | 6 (0.1) | 0 | 2 (0.6) | 2 (50.0) | 0 (0.0) | 0 |
| **Hypertrophic cardiomyopathy (%)** | 8 (0.1) | 7 (8.0) | 1 (0.0) | 0 | 1 (0.3) | 1 (25.0) | 0 (0.0) | 0 |
| **Ventricular arrhythmias (%)** | 33 (0.3) | 3 (3.4) | 30 (0.3) | 0 | 7 (2.0) | 1 (25.0) | 6 (1.7) | 0 |
| **Atrial arrhythmias (%)** | 191 (1.9) | 19 (21.8) | 172 (1.7) | 0 | 7 (2.0) | 0 (0.0) | 7 (2.0) | 0 |
| **Heart arrhythmia (%)** | 54 (0.5) | 2 (2.3) | 52 (0.5) | 0 | 6 (1.7) | 1 (25.0) | 5 (1.5) | 0 |
| **Chronic ischemic heart disease (%)** | 725 (7.3) | 0 (0.0) | 725 (7.3) | 0 | 35 (10.1) | 0 (0.0) | 35 (10.2) | 0 |
| **Acute myocardial infarction (%)** | 298 (3.0) | 1 (1.1) | 297 (3.0) | 0 | 15 (4.3) | 0 (0.0) | 15 (4.4) | 0 |
| **Cardiac arrest (%)** | 34 (0.3) | 1 (1.1) | 33 (0.3) | 0 | 0 (0.0) | 0 (0.0) | 0 (0.0) | 0 |
| **Angina pectoris (%)** | 312 (3.1) | 2 (2.3) | 310 (3.1) | 0 | 16 (4.6) | 0 (0.0) | 16 (4.7) | 0 |
| **Conduction disorders (%)** | 151 (1.5) | 10 (11.5) | 141 (1.4) | 0 | 8 (2.3) | 0 (0.0) | 8 (2.3) | 0 |
| **Valvular disease (%)** | 241 (2.4) | 23 (26.4) | 218 (2.2) | 0 | 11 (3.2) | 1 (25.0) | 10 (2.9) | 0 |
| **Congenital heart disease (%)** | 28 (0.3) | 3 (3.4) | 25 (0.3) | 0 | 2 (0.6) | 0 (0.0) | 2 (0.6) | 0 |
| **Pulmonary obstructive disease (%)** | 494 (5.0) | 24 (27.6) | 470 (4.8) | 0 | 25 (7.2) | 0 (0.0) | 25 (7.3) | 0 |
| **Cardiovascular death (%)** | 181 (1.8) | 13 (14.9) | 168 (1.7) | 0 | 11 (3.2) | 0 (0.0) | 11 (3.2) | 0 |
| **All-cause mortality (%)** | 513 (5.1) | 27 (31.0) | 486 (4.9) | 0 | 19 (5.5) | 0 (0.0) | 19 (5.5) | 0 |
| ECG MEASUREMENTS | | | | | | | | |
| **n (%)** | 1,062 (10.6) | 4 (4.6) | 1,058 (10.7) | | 32 (9.2) | 0 (0.0) | 32 (9.3) | |
| **P duration (median [IQR])** | 100.00 [90.00, 108.00] | 90.00 [82.00, 111.00] | 100.00 [90.00, 108.00] | 89.8 | 100.00 [93.50, 111.50] | NA | 100.00 [93.50, 111.50] | 90.8 |
| **P axis (median [IQR])** | 55.00 [40.25, 67.00] | 47.00 [47.00, 47.00] | 55.00 [40.00, 67.00] | 93.1 | 54.00 [42.25, 61.50] | NA | 54.00 [42.25, 61.50] | 94.8 |
| **PQ interval (median [IQR])** | 160.00 [145.50, 178.00] | 188.00 [188.00, 188.00] | 160.00 [145.00, 178.00] | 93.1 | 171.00 [147.00, 183.00] | NA | 171.00 [147.00, 183.00] | 94.8 |
| **QRS duration (median [IQR])** | 86.00 [80.00, 94.00] | 93.00 [89.50, 97.00] | 86.00 [80.00, 94.00] | 89.4 | 88.00 [81.50, 96.00] | NA | 88.00 [81.50, 96.00] | 90.8 |
| **R axis (median [IQR])** | 34.00 [7.00, 58.00] | -48.00 [-48.00, -48.00] | 34.00 [7.50, 58.00] | 92.9 | 23.50 [-1.75, 50.00] | NA | 23.50 [-1.75, 50.00] | 94.8 |
| **QTC interval (median [IQR])** | 417.00 [402.00, 433.00] | 511.00 [511.00, 511.00] | 417.00 [402.00, 432.50] | 92.9 | 429.50 [403.25, 440.00] | NA | 429.50 [403.25, 440.00] | 94.8 |
| **T axis (median [IQR])** | 40.00 [23.00, 55.25] | 92.00 [92.00, 92.00] | 40.00 [23.00, 55.00] | 92.9 | 35.50 [20.25, 54.25] | NA | 35.50 [20.25, 54.25] | 94.8 |
| CMR MEASUREMENTS | | | | | | | | |
| **n (%)** | 990 (9.9) | 4 (4.6) | 986 (10.0) | | 33 (9.5) | 0 (0.0) | 33 (9.6) | |
| **RVEDVi (median [IQR])** | 80.18 [70.62, 90.27] | 79.28 [79.28, 79.28] | 80.19 [70.61, 90.27] | 91 | 79.14 [73.73, 92.49] | NA | 79.14 [73.73, 92.49] | 91.1 |
| **RVESVi (median [IQR])** | 32.89 [27.38, 39.68] | 20.93 [20.93, 20.93] | 32.90 [27.42, 39.70] | 91 | 35.16 [29.98, 38.70] | NA | 35.16 [29.98, 38.70] | 91.1 |
| **RVSVi (median [IQR])** | 46.55 [40.92, 52.84] | 58.35 [58.35, 58.35] | 46.53 [40.91, 52.81] | 91 | 48.22 [41.97, 52.24] | NA | 48.22 [41.97, 52.24] | 91.1 |
| **RVEF (median [IQR])** | 58.38 [54.19, 62.76] | 73.60 [73.60, 73.60] | 58.37 [54.19, 62.74] | 91 | 58.30 [53.30, 62.06] | NA | 58.30 [53.30, 62.06] | 91.1 |
| **RVPER (median [IQR])** | 388.72 [316.56, 465.82] | 446.55 [446.55, 446.55] | 388.66 [316.54, 465.97] | 91 | 405.50 [291.73, 489.37] | NA | 405.50 [291.73, 489.37] | 91.1 |
| **RVPFR (median [IQR])** | 300.61 [245.17, 364.00] | 373.24 [373.24, 373.24] | 300.34 [245.12, 363.44] | 91 | 302.85 [225.65, 375.82] | NA | 302.85 [225.65, 375.82] | 91.1 |
| **RVPAFR (median [IQR])** | 282.95 [222.68, 360.32] | 547.72 [547.72, 547.72] | 282.86 [222.56, 360.07] | 91 | 274.74 [213.71, 343.90] | NA | 274.74 [213.71, 343.90] | 91.1 |
| **LVEDVi (median [IQR])** | 74.33 [66.34, 83.11] | 64.68 [59.39, 69.33] | 74.37 [66.38, 83.15] | 91.9 | 80.77 [73.11, 88.68] | NA | 80.77 [73.11, 88.68] | 91.6 |
| **LVESVi (median [IQR])** | 30.02 [25.12, 35.70] | 26.43 [25.24, 28.39] | 30.02 [25.13, 35.72] | 91.9 | 31.74 [25.91, 39.55] | NA | 31.74 [25.91, 39.55] | 91.6 |
| **LVSVi (median [IQR])** | 44.03 [39.34, 50.28] | 38.25 [31.00, 44.09] | 44.05 [39.37, 50.30] | 91.9 | 46.82 [43.25, 50.82] | NA | 46.82 [43.25, 50.82] | 91.6 |
| **LVEF (median [IQR])** | 59.47 [55.29, 63.52] | 59.14 [51.51, 63.31] | 59.48 [55.29, 63.52] | 91.9 | 59.69 [56.59, 66.23] | NA | 59.69 [56.59, 66.23] | 91.6 |
| **LVPER (median [IQR])** | 373.80 [302.29, 452.71] | 284.40 [256.83, 364.56] | 373.81 [302.44, 453.27] | 91.9 | 407.32 [307.20, 455.45] | NA | 407.32 [307.20, 455.45] | 91.6 |
| **LVPFR (median [IQR])** | 321.31 [259.23, 385.16] | 201.74 [189.85, 229.92] | 321.49 [259.95, 385.63] | 91.9 | 346.24 [290.79, 422.04] | NA | 346.24 [290.79, 422.04] | 91.6 |
| **LVPAFR (median [IQR])** | 233.66 [167.35, 306.30] | 363.87 [208.97, 466.86] | 233.50 [167.46, 305.08] | 91.9 | 208.66 [158.63, 298.40] | NA | 208.66 [158.63, 298.40] | 91.6 |
| **LVEDMi (median [IQR])** | 41.88 [36.52, 48.62] | 45.76 [34.02, 48.75] | 41.85 [36.55, 48.61] | 91.9 | 42.81 [36.04, 48.38] | NA | 42.81 [36.04, 48.38] | 91.6 |
| **LVMVR (median [IQR])** | 0.56 [0.50, 0.62] | 0.70 [0.56, 0.70] | 0.56 [0.50, 0.62] | 91.9 | 0.55 [0.49, 0.60] | NA | 0.55 [0.49, 0.60] | 91.6 |
| **LVEDV/RVEDV (median [IQR])** | 0.93 [0.86, 1.03] | 0.93 [0.93, 0.93] | 0.93 [0.86, 1.03] | 92.2 | 0.94 [0.90, 1.05] | NA | 0.94 [0.90, 1.05] | 91.6 |
| **LVESV/RVESV (median [IQR])** | 0.91 [0.80, 1.04] | 1.15 [1.15, 1.15] | 0.91 [0.80, 1.04] | 92.2 | 0.91 [0.82, 1.00] | NA | 0.91 [0.82, 1.00] | 91.6 |
| **peakEcc (median [IQR])** | -22.70 [-24.98, -20.43] | -21.60 [-21.61, -21.59] | -22.72 [-24.98, -20.42] | 93.7 | -22.87 [-26.90, -21.63] | NA | -22.87 [-26.90, -21.63] | 94.2 |
| **TPKEcc (median [IQR])** | 331.31 [309.83, 354.66] | 325.97 [321.27, 330.67] | 331.31 [309.75, 354.68] | 93.7 | 326.90 [318.45, 363.83] | NA | 326.90 [318.45, 363.83] | 94.2 |
| **peakEll2Ch (median [IQR])** | -21.19 [-23.34, -18.93] | -23.89 [-23.89, -23.89] | -21.17 [-23.32, -18.93] | 93.8 | -21.37 [-23.84, -19.31] | NA | -21.37 [-23.84, -19.31] | 93.9 |
| **TPKEll2Ch (median [IQR])** | 349.86 [321.84, 379.10] | 388.50 [388.50, 388.50] | 349.86 [321.81, 379.08] | 93.9 | 346.80 [321.44, 370.50] | NA | 346.80 [321.44, 370.50] | 93.9 |
| **peakEll4Ch (median [IQR])** | -23.30 [-25.97, -21.37] | -21.40 [-21.40, -21.40] | -23.30 [-25.99, -21.37] | 93.9 | -24.25 [-26.79, -21.39] | NA | -24.25 [-26.79, -21.39] | 93.7 |
| **TPKEll4Ch (median [IQR])** | 357.53 [327.07, 397.64] | 360.78 [360.78, 360.78] | 357.30 [326.96, 397.81] | 93.9 | 354.30 [328.00, 406.56] | NA | 354.30 [328.00, 406.56] | 93.9 |
| **Wall thickness segment 1 (median [IQR])** | 7.65 [6.81, 8.50] | 7.04 [5.94, 8.14] | 7.65 [6.81, 8.49] | 93.2 | 7.05 [6.26, 8.57] | NA | 7.05 [6.26, 8.57] | 94.2 |
| **Wall thickness segment 2 (median [IQR])** | 6.75 [5.75, 7.91] | 7.50 [6.70, 8.31] | 6.75 [5.74, 7.90] | 93.2 | 6.81 [5.24, 7.75] | NA | 6.81 [5.24, 7.75] | 94.2 |
| **Wall thickness segment 3 (median [IQR])** | 6.05 [5.17, 6.96] | 7.60 [6.78, 8.42] | 6.05 [5.17, 6.95] | 93.2 | 5.58 [4.74, 7.16] | NA | 5.58 [4.74, 7.16] | 94.2 |
| **Wall thickness segment 4 (median [IQR])** | 6.54 [5.82, 7.22] | 8.31 [7.66, 8.97] | 6.54 [5.81, 7.21] | 93.2 | 6.06 [5.49, 6.76] | NA | 6.06 [5.49, 6.76] | 94.2 |
| **Wall thickness segment 5 (median [IQR])** | 6.20 [5.62, 6.96] | 6.33 [5.27, 7.39] | 6.20 [5.62, 6.96] | 93.2 | 5.96 [5.48, 6.40] | NA | 5.96 [5.48, 6.40] | 94.2 |
| **Wall thickness segment 6 (median [IQR])** | 6.55 [5.97, 7.31] | 6.61 [5.42, 7.79] | 6.55 [5.98, 7.31] | 93.2 | 6.45 [6.15, 6.79] | NA | 6.45 [6.15, 6.79] | 94.2 |
| **Wall thickness segment 7 (median [IQR])** | 5.73 [5.28, 6.31] | 5.48 [4.90, 6.07] | 5.73 [5.29, 6.31] | 93.2 | 5.52 [5.21, 6.06] | NA | 5.52 [5.21, 6.06] | 94.2 |
| **Wall thickness segment 8 (median [IQR])** | 7.01 [6.28, 7.76] | 8.52 [7.77, 9.27] | 7.01 [6.28, 7.75] | 93.2 | 6.97 [6.44, 7.28] | NA | 6.97 [6.44, 7.28] | 94.2 |
| **Wall thickness segment 9 (median [IQR])** | 7.38 [6.48, 8.25] | 9.47 [8.05, 10.89] | 7.38 [6.48, 8.25] | 93.2 | 6.96 [6.18, 7.46] | NA | 6.96 [6.18, 7.46] | 94.2 |
| **Wall thickness segment 10 (median [IQR])** | 6.23 [5.59, 6.96] | 7.07 [6.08, 8.06] | 6.23 [5.60, 6.96] | 93.2 | 5.88 [5.33, 6.23] | NA | 5.88 [5.33, 6.23] | 94.2 |
| **Wall thickness segment 11 (median [IQR])** | 5.62 [5.09, 6.32] | 5.57 [5.19, 5.95] | 5.62 [5.10, 6.32] | 93.2 | 5.42 [4.96, 5.81] | NA | 5.42 [4.96, 5.81] | 94.2 |
| **Wall thickness segment 12 (median [IQR])** | 5.60 [5.22, 6.25] | 5.56 [5.53, 5.60] | 5.60 [5.22, 6.26] | 93.2 | 5.38 [5.17, 6.12] | NA | 5.38 [5.17, 6.12] | 94.2 |
| **Wall thickness segment 13 (median [IQR])** | 5.48 [5.10, 5.91] | 8.01 [7.03, 9.00] | 5.48 [5.10, 5.91] | 93.2 | 5.38 [5.24, 5.80] | NA | 5.38 [5.24, 5.80] | 94.2 |
| **Wall thickness segment 14 (median [IQR])** | 6.00 [5.36, 6.66] | 8.33 [6.87, 9.79] | 6.00 [5.36, 6.66] | 93.2 | 5.92 [5.41, 6.60] | NA | 5.92 [5.41, 6.60] | 94.2 |
| **Wall thickness segment 15 (median [IQR])** | 5.09 [4.48, 5.69] | 7.21 [5.80, 8.63] | 5.09 [4.48, 5.69] | 93.2 | 5.12 [4.78, 5.37] | NA | 5.12 [4.78, 5.37] | 94.2 |
| **Wall thickness segment 16 (median [IQR])** | 5.26 [4.80, 5.72] | 8.31 [6.87, 9.75] | 5.26 [4.80, 5.72] | 93.2 | 5.32 [4.86, 5.67] | NA | 5.32 [4.86, 5.67] | 94.2 |
| **Global wall thickness (median [IQR])** | 6.30 [5.73, 6.84] | 7.33 [7.28, 7.38] | 6.30 [5.72, 6.84] | 93.2 | 6.07 [5.57, 6.29] | NA | 6.07 [5.57, 6.29] | 94.2 |
| **Septal wall thickness (median [IQR])** | 6.68 [5.88, 7.35] | 8.28 [7.88, 8.69] | 6.67 [5.88, 7.34] | 93.2 | 6.43 [5.69, 7.01] | NA | 6.43 [5.69, 7.01] | 94.2 |
| **Maximum wall thickness (median [IQR])** | 8.10 [7.24, 9.02] | 10.96 [10.29, 11.64] | 8.09 [7.24, 9.01] | 93.2 | 7.81 [6.97, 8.59] | NA | 7.81 [6.97, 8.59] | 94.2 |

Abbreviations:
ARVC: arrythmogenic right ventricular cardiomyopathy; BMI: body mass index; CMR: cardiac magnetic resonance imaging; DCM: dilated cardiomyopathy; ECG: Electrocardiography; EDVi: indexed end-diastolic volume; EDMi: indexed end-diastolic mass; EF: ejection fraction; ESVi: indexed end-systolic volume; G+: carriers of likely pathogenic and pathogenic variants associated with one of the cardiomyopathies; HCM: hypertrophic cardiomyopathy; HDL: high-density lipoprotein; IQR: interquartile range; LDL: low-density lipoprotein; LV: left ventricular; MET: metabolic equivalent of task; MVR: mass to volume ratio; PAFR: peak atrial filling rate; peakEcc: peak circumferential strain; peakEll2Ch: longitudinal strain analyzed in 2-chamber view; peakEll4Ch: longitudinal strain analyzed in 4-chamber view; PER: peak ejection rate; PFR: peak filling rate; RV: right ventricular; SVi: indexed stroke volume; TPKEcc: global time to peak circumferential strain; TPKEll2Ch: global time to longitudinal strain analyzed in 2-chamber view; TPKEll4Ch: global time to longitudinal strain analyzed in 4-chamber view.

**Supplementary Table III: Extensive baseline table**

| | Controls G- | | | | DCM G+ | | | |
|---|---|---|---|---|---|---|---|---|
| | Overall | Diagnosed | Non-Diagnosed | Missing | Overall | Diagnosed | Non-Diagnosed | Missing |
| n (%) | 9,972 (100) | 87 (0.8) | 9,885 (99.2) | | 800 (100) | 25 (3.1) | 775 (96.9) | |
| **Sex = Female (%)** | 5,436 (54.5) | 35 (40.2) | 5,401 (54.6) | 0 | 450 (56.2) | 15 (60.0) | 435 (56.1) | 0 |
| **Age (median [IQR])** | 57.00 [49.00, 63.00] | 62.00 [56.00, 66.00] | 57.00 [49.00, 63.00] | 0 | 58.00 [50.75, 64.00] | 62.00 [53.00, 66.00] | 57.00 [50.00, 63.00] | 0 |
| **Ethnicity (%)** | | | | 1 | | | | 0.6 |
| Asian | 1,076 (10.9) | 5 (5.8) | 1071 (10.9) | | 8 (1.0) | 1 (4.0) | 7 (0.9) | |
| Black | 164 (1.7) | 3 (3.5) | 161 (1.6) | | 12 (1.5) | 1 (4.0) | 11 (1.4) | |
| Chinese | 56 (0.6) | 1 (1.2) | 55 (0.6) | | 2 (0.3) | 0 (0.0) | 2 (0.3) | |
| Mixed | 132 (1.3) | 2 (2.3) | 130 (1.3) | | 4 (0.5) | 0 (0.0) | 4 (0.5) | |
| Other | 160 (1.6) | 1 (1.2) | 159 (1.6) | | 9 (1.1) | 0 (0.0) | 9 (1.2) | |
| White | 8,288 (83.9) | 74 (86.0) | 8,214 (83.9) | | 760 (95.6) | 23 (92.0) | 737 (95.7) | |
| **CARDIOVASCULAR RISK FACTORS** | | | | | | | | |
| **BMI (median [IQR])** | 26.73 [24.15, 29.82] | 28.83 [25.33, 32.20] | 26.71 [24.15, 29.80] | 0.5 | 26.92 [24.06, 29.90] | 27.69 [24.21, 30.90] | 26.92 [24.06, 29.88] | 0.4 |
| **Diabetes (%)** | 914 (9.2) | 21 (24.1) | 893 (9.0) | 0 | 62 (7.8) | 1 (4.0) | 61 (7.9) | 0 |
| **Hypertension (%)** | 3,420 (34.3) | 60 (69.0) | 3,360 (34.0) | 0 | 287 (35.9) | 18 (72.0) | 269 (34.7) | 0 |
| **Mean systolic blood pressure (median [IQR])** | 135.50 [124.00, 149.00] | 144.00 [130.00, 155.50] | 135.50 [124.00, 149.00] | 0.1 | 136.00 [124.00, 149.00] | 136.50 [129.50, 149.50] | 135.75 [124.00, 149.00] | 0.1 |
| **Mean diastolic blood pressure (median [IQR])** | 82.00 [75.00, 88.50] | 84.00 [76.50, 91.50] | 81.50 [75.00, 88.50] | 0.1 | 81.50 [74.50, 89.00] | 80.50 [77.00, 88.50] | 81.50 [74.50, 89.00] | 0.1 |
| **Hypercholesterolaemia (%)** | 2,416 (24.2) | 34 (39.1) | 2,382 (24.1) | 0 | 211 (26.4) | 13 (52.0) | 198 (25.5) | 0 |
| **Total cholesterol (median [IQR])** | 5.61 [4.86, 6.38] | 5.23 [4.39, 6.10] | 5.61 [4.87, 6.38] | 4.3 | 5.61 [4.90, 6.33] | 5.20 [4.79, 6.24] | 5.62 [4.93, 6.34] | 4.2 |
| **HDL (median [IQR])** | 1.38 [1.16, 1.65] | 1.29 [1.13, 1.56] | 1.38 [1.16, 1.65] | 11.3 | 1.40 [1.17, 1.65] | 1.47 [1.29, 1.69] | 1.40 [1.17, 1.65] | 11.9 |
| **LDL (median [IQR])** | 3.50 [2.92, 4.09] | 3.22 [2.54, 3.81] | 3.50 [2.92, 4.09] | 4.6 | 3.46 [2.93, 4.09] | 3.23 [2.65, 4.02] | 3.47 [2.94, 4.09] | 4.2 |
| **Ever Smoked (%)** | 4,132 (41.4) | 50 (57.5) | 4,082 (41.3) | 0 | 371 (46.4) | 11 (44.0) | 360 (46.5) | 0 |
| **MET minutes per week for walking (median [IQR])** | 693.00 [297.00, 1,386.00] | 528.00 [255.75, 1,608.75] | 693.00 [297.00, 1,386.00] | 19.8 | 577.50 [255.75, 1,386.00] | 478.50 [206.25, 767.25] | 577.50 [264.00, 1,386.00] | 21.1 |
| **MET minutes per week for moderate activity (median [IQR])** | 480.00 [120.00, 1,200.00] | 480.00 [100.00, 1,680.00] | 480.00 [120.00, 1,200.00] | 19.8 | 480.00 [120.00, 1,200.00] | 400.00 [50.00, 795.00] | 480.00 [120.00, 1,200.00] | 21.1 |
| **MET minutes per week for vigorous activity (median [IQR])** | 240.00 [0.00, 960.00] | 0.00 [0.00, 820.00] | 240.00 [0.00, 960.00] | 19.8 | 240.00 [0.00, 960.00] | 40.00 [0.00, 540.00] | 240.00 [0.00, 960.00] | 21.1 |
| **Total MET minutes per week (median [IQR])** | 1,773.00 [810.00, 3,452.50] | 1,367.50 [478.50, 3,834.00] | 1,776.50 [810.00, 3,450.00] | 19.8 | 1,695.00 [783.75, 3,536.00] | 1,050.50 [681.75, 2,846.38] | 1,706.00 [795.00, 3,539.00] | 21.1 |
| **Family heart disease (%)** | 4,458 (44.7) | 34 (39.1) | 4,424 (44.8) | 0 | 380 (47.5) | 9 (36.0) | 371 (47.9) | 0 |
| **CARDIAC DISEASES/OUTCOMES** | | | | | | | | |
| **Cardiac problem (%)** | 41 (0.4) | 2 (2.3) | 39 (0.4) | 0 | 3 (0.4) | 0 (0.0) | 3 (0.4) | 0 |
| **Heart failure (%)** | 182 (1.8) | 74 (85.1) | 108 (1.1) | 0 | 36 (4.5) | 16 (64.0) | 20 (2.6) | 0 |
| **Cardiomyopathy (%)** | 37 (0.4) | 26 (29.9) | 11 (0.1) | 0 | 22 (2.8) | 16 (64.0) | 6 (0.8) | 0 |
| **Dilated cardiomyopathy (%)** | 14 (0.1) | 8 (9.2) | 6 (0.1) | 0 | 9 (1.1) | 7 (28.0) | 2 (0.3) | 0 |
| **Hypertrophic cardiomyopathy (%)** | 8 (0.1) | 7 (8.0) | 1 (0.0) | 0 | 7 (0.9) | 6 (24.0) | 1 (0.1) | 0 |
| **Ventricular arrhythmias (%)** | 33 (0.3) | 3 (3.4) | 30 (0.3) | 0 | 13 (1.6) | 5 (20.0) | 8 (1.0) | 0 |
| **Atrial arrhythmias (%)** | 191 (1.9) | 19 (21.8) | 172 (1.7) | 0 | 34 (4.2) | 6 (24.0) | 28 (3.6) | 0 |
| **Heart arrhythmia (%)** | 54 (0.5) | 2 (2.3) | 52 (0.5) | 0 | 12 (1.5) | 2 (8.0) | 10 (1.3) | 0 |
| **Chronic ischemic heart disease (%)** | 725 (7.3) | 0 (0.0) | 725 (7.3) | 0 | 73 (9.1) | 0 (0.0) | 73 (9.4) | 0 |
| **Acute myocardial infarction (%)** | 298 (3.0) | 1 (1.1) | 297 (3.0) | 0 | 27 (3.4) | 1 (4.0) | 26 (3.4) | 0 |
| **Cardiac arrest (%)** | 34 (0.3) | 1 (1.1) | 33 (0.3) | 0 | 6 (0.8) | 1 (4.0) | 5 (0.6) | 0 |
| **Angina pectoris (%)** | 312 (3.1) | 2 (2.3) | 310 (3.1) | 0 | 30 (3.8) | 0 (0.0) | 30 (3.9) | 0 |
| **Conduction disorders (%)** | 151 (1.5) | 10 (11.5) | 141 (1.4) | 0 | 18 (2.2) | 3 (12.0) | 15 (1.9) | 0 |
| **Valvular disease (%)** | 241 (2.4) | 23 (26.4) | 218 (2.2) | 0 | 37 (4.6) | 9 (36.0) | 28 (3.6) | 0 |
| **Congenital heart disease (%)** | 28 (0.3) | 3 (3.4) | 25 (0.3) | 0 | 3 (0.4) | 1 (4.0) | 2 (0.3) | 0 |
| **Pulmonary obstructive disease (%)** | 494 (5.0) | 24 (27.6) | 470 (4.8) | 0 | 47 (5.9) | 7 (28.0) | 40 (5.2) | 0 |
| **Cardiovascular death (%)** | 181 (1.8) | 13 (14.9) | 168 (1.7) | 0 | 24 (3.0) | 5 (20.0) | 19 (2.5) | 0 |
| **All-cause mortality (%)** | 513 (5.1) | 27 (31.0) | 486 (4.9) | 0 | 56 (7.0) | 8 (32.0) | 48 (6.2) | 0 |
| **ECG MEASUREMENTS** | | | | | | | | |
| **n (%)** | 1,062 (10.6) | 4 (4.6) | 1,058 (10.7) | | 87 (10.9) | 0 (0.0) | 87 (11.2) | |
| **P duration (median [IQR])** | 100.00 [90.00, 108.00] | 90.00 [82.00, 111.00] | 100.00 [90.00, 108.00] | 89.8 | 99.00 [86.00, 106.00] | NA | 99.00 [86.00, 106.00] | 90 |
| **P axis (median [IQR])** | 55.00 [40.25, 67.00] | 47.00 [47.00, 47.00] | 55.00 [40.00, 67.00] | 93.1 | 49.00 [36.50, 61.00] | NA | 49.00 [36.50, 61.00] | 92.6 |
| **PQ interval (median [IQR])** | 160.00 [145.50, 178.00] | 188.00 [188.00, 188.00] | 160.00 [145.00, 178.00] | 93.1 | 164.00 [145.00, 176.00] | NA | 164.00 [145.00, 176.00] | 92.6 |
| **QRS duration (median [IQR])** | 86.00 [80.00, 94.00] | 93.00 [89.50, 97.00] | 86.00 [80.00, 94.00] | 89.4 | 84.00 [78.00, 92.00] | NA | 84.00 [78.00, 92.00] | 89.1 |
| **R axis (median [IQR])** | 34.00 [7.00, 58.00] | -48.00 [-48.00, -48.00] | 34.00 [7.50, 58.00] | 92.9 | 26.00 [-3.50, 50.00] | NA | 26.00 [-3.50, 50.00] | 92.1 |
| **QTC interval (median [IQR])** | 417.00 [402.00, 433.00] | 511.00 [511.00, 511.00] | 417.00 [402.00, 432.50] | 92.9 | 420.00 [404.00, 435.00] | NA | 420.00 [404.00, 435.00] | 92.1 |
| **T axis (median [IQR])** | 40.00 [23.00, 55.25] | 92.00 [92.00, 92.00] | 40.00 [23.00, 55.00] | 92.9 | 42.00 [25.50, 57.00] | NA | 42.00 [25.50, 57.00] | 92.1 |
| **CMR MEASUREMENTS** | | | | | | | | |
| **n (%)** | 990 (9.9) | 4 (4.6) | 986 (10.0) | | 87 (10.9) | 0 (0.0) | 87 (11.2) | |
| **RVEDVi (median [IQR])** | 80.18 [70.62, 90.27] | 79.28 [79.28, 79.28] | 80.19 [70.61, 90.27] | 91 | 76.54 [69.50, 84.81] | NA | 76.54 [69.50, 84.81] | 90.2 |
| **RVESVi (median [IQR])** | 32.89 [27.38, 39.68] | 20.93 [20.93, 20.93] | 32.90 [27.42, 39.70] | 91 | 32.21 [27.10, 37.43] | NA | 32.21 [27.10, 37.43] | 90.2 |
| **RVSVi (median [IQR])** | 46.55 [40.92, 52.84] | 58.35 [58.35, 58.35] | 46.53 [40.91, 52.81] | 91 | 44.50 [40.74, 51.29] | NA | 44.50 [40.74, 51.29] | 90.2 |
| **RVEF (median [IQR])** | 58.38 [54.19, 62.76] | 73.60 [73.60, 73.60] | 58.37 [54.19, 62.74] | 91 | 59.31 [52.99, 62.59] | NA | 59.31 [52.99, 62.59] | 90.2 |
| **RVPER (median [IQR])** | 388.72 [316.56, 465.82] | 446.55 [446.55, 446.55] | 388.66 [316.54, 465.97] | 91 | 361.23 [290.07, 443.94] | NA | 361.23 [290.07, 443.94] | 90.2 |
| **RVPFR (median [IQR])** | 300.61 [245.17, 364.00] | 373.24 [373.24, 373.24] | 300.34 [245.12, 363.44] | 91 | 295.78 [220.42, 343.17] | NA | 295.78 [220.42, 343.17] | 90.2 |
| **RVPAFR (median [IQR])** | 282.95 [222.68, 360.32] | 547.72 [547.72, 547.72] | 282.86 [222.56, 360.07] | 91 | 275.01 [224.94, 344.91] | NA | 275.01 [224.94, 344.91] | 90.2 |
| **LVEDVi (median [IQR])** | 74.33 [66.34, 83.11] | 64.68 [59.39, 69.33] | 74.37 [66.38, 83.15] | 91.9 | 77.32 [68.06, 86.15] | NA | 77.32 [68.06, 86.15] | 91.4 |
| **LVESVi (median [IQR])** | 30.02 [25.12, 35.70] | 26.43 [25.24, 28.39] | 30.02 [25.13, 35.72] | 91.9 | 31.69 [26.19, 39.84] | NA | 31.69 [26.19, 39.84] | 91.4 |
| **LVSVi (median [IQR])** | 44.03 [39.34, 50.28] | 38.25 [31.00, 44.09] | 44.05 [39.37, 50.30] | 91.9 | 43.18 [37.50, 49.11] | NA | 43.18 [37.50, 49.11] | 91.4 |
| **LVEF (median [IQR])** | 59.47 [55.29, 63.52] | 59.14 [51.51, 63.31] | 59.48 [55.29, 63.52] | 91.9 | 57.34 [52.60, 62.80] | NA | 57.34 [52.60, 62.80] | 91.4 |
| **LVPER (median [IQR])** | 373.80 [302.29, 452.71] | 284.40 [256.83, 364.56] | 373.81 [302.44, 453.27] | 91.9 | 339.21 [258.97, 430.85] | NA | 339.21 [258.97, 430.85] | 91.4 |
| **LVPFR (median [IQR])** | 321.31 [259.23, 385.16] | 201.74 [189.85, 229.92] | 321.49 [259.95, 385.63] | 91.9 | 314.20 [258.81, 366.77] | NA | 314.20 [258.81, 366.77] | 91.4 |
| **LVPAFR (median [IQR])** | 233.66 [167.35, 306.30] | 363.87 [208.97, 466.86] | 233.50 [167.46, 305.08] | 91.9 | 253.35 [178.90, 330.36] | NA | 253.35 [178.90, 330.36] | 91.4 |
| **LVEDMi (median [IQR])** | 41.88 [36.52, 48.62] | 45.76 [34.02, 48.75] | 41.85 [36.55, 48.61] | 91.9 | 42.96 [36.56, 46.70] | NA | 42.96 [36.56, 46.70] | 91.4 |
| **LVMVR (median [IQR])** | 0.56 [0.50, 0.62] | 0.70 [0.56, 0.70] | 0.56 [0.50, 0.62] | 91.9 | 0.54 [0.49, 0.59] | NA | 0.54 [0.49, 0.59] | 91.4 |
| **LVEDV/RVEDV (median [IQR])** | 0.93 [0.86, 1.03] | 0.93 [0.93, 0.93] | 0.93 [0.86, 1.03] | 92.2 | 1.00 [0.91, 1.08] | NA | 1.00 [0.91, 1.08] | 91.8 |
| **LVESV/RVESV (median [IQR])** | 0.91 [0.80, 1.04] | 1.15 [1.15, 1.15] | 0.91 [0.80, 1.04] | 92.2 | 1.02 [0.89, 1.19] | NA | 1.02 [0.89, 1.19] | 91.8 |
| **peakEcc (median [IQR])** | -22.70 [-24.98, -20.43] | -21.60 [-21.61, -21.59] | -22.72 [-24.98, -20.42] | 93.7 | -22.67 [-24.40, -19.13] | NA | -22.67 [-24.40, -19.13] | 92.8 |
| **TPKEcc (median [IQR])** | 331.31 [309.83, 354.66] | 325.97 [321.27, 330.67] | 331.31 [309.75, 354.68] | 93.7 | 334.71 [320.31, 360.48] | NA | 334.71 [320.31, 360.48] | 92.8 |
| **peakEll2Ch (median [IQR])** | -21.19 [-23.34, -18.93] | -23.89 [-23.89, -23.89] | -21.17 [-23.32, -18.93] | 93.8 | -20.29 [-22.24, -17.98] | NA | -20.29 [-22.24, -17.98] | 93 |
| **TPKEll2Ch (median [IQR])** | 349.86 [321.84, 379.10] | 388.50 [388.50, 388.50] | 349.86 [321.81, 379.08] | 93.9 | 353.29 [331.00, 381.68] | NA | 353.29 [331.00, 381.68] | 93 |
| **peakEll4Ch (median [IQR])** | -23.30 [-25.97, -21.37] | -21.40 [-21.40, -21.40] | -23.30 [-25.99, -21.37] | 93.9 | -22.30 [-24.57, -19.76] | NA | -22.30 [-24.57, -19.76] | 93 |
| **TPKEll4Ch (median [IQR])** | 357.53 [327.07, 397.64] | 360.78 [360.78, 360.78] | 357.30 [326.96, 397.81] | 93.9 | 354.80 [325.41, 392.55] | NA | 354.80 [325.41, 392.55] | 93.1 |
| **Wall thickness segment 1 (median [IQR])** | 7.65 [6.81, 8.50] | 7.04 [5.94, 8.14] | 7.65 [6.81, 8.49] | 93.2 | 7.44 [6.78, 8.21] | NA | 7.44 [6.78, 8.21] | 93.4 |
| **Wall thickness segment 2 (median [IQR])** | 6.75 [5.75, 7.91] | 7.50 [6.70, 8.31] | 6.75 [5.74, 7.90] | 93.2 | 6.03 [5.31, 7.39] | NA | 6.03 [5.31, 7.39] | 93.4 |
| **Wall thickness segment 3 (median [IQR])** | 6.05 [5.17, 6.96] | 7.60 [6.78, 8.42] | 6.05 [5.17, 6.95] | 93.2 | 6.10 [4.85, 6.66] | NA | 6.10 [4.85, 6.66] | 93.4 |
| **Wall thickness segment 4 (median [IQR])** | 6.54 [5.82, 7.22] | 8.31 [7.66, 8.97] | 6.54 [5.81, 7.21] | 93.2 | 6.57 [5.89, 6.99] | NA | 6.57 [5.89, 6.99] | 93.4 |
| **Wall thickness segment 5 (median [IQR])** | 6.20 [5.62, 6.96] | 6.33 [5.27, 7.39] | 6.20 [5.62, 6.96] | 93.2 | 6.08 [5.57, 6.50] | NA | 6.08 [5.57, 6.50] | 93.4 |
| **Wall thickness segment 6 (median [IQR])** | 6.55 [5.97, 7.31] | 6.61 [5.42, 7.79] | 6.55 [5.98, 7.31] | 93.2 | 6.58 [6.05, 7.01] | NA | 6.58 [6.05, 7.01] | 93.4 |
| **Wall thickness segment 7 (median [IQR])** | 5.73 [5.28, 6.31] | 5.48 [4.90, 6.07] | 5.73 [5.29, 6.31] | 93.2 | 5.56 [5.28, 6.14] | NA | 5.56 [5.28, 6.14] | 93.4 |
| **Wall thickness segment 8 (median [IQR])** | 7.01 [6.28, 7.76] | 8.52 [7.77, 9.27] | 7.01 [6.28, 7.75] | 93.2 | 6.90 [6.27, 7.47] | NA | 6.90 [6.27, 7.47] | 93.4 |
| **Wall thickness segment 9 (median [IQR])** | 7.38 [6.48, 8.25] | 9.47 [8.05, 10.89] | 7.38 [6.48, 8.25] | 93.2 | 7.18 [6.64, 7.85] | NA | 7.18 [6.64, 7.85] | 93.4 |
| **Wall thickness segment 10 (median [IQR])** | 6.23 [5.59, 6.96] | 7.07 [6.08, 8.06] | 6.23 [5.60, 6.96] | 93.2 | 6.28 [5.90, 6.89] | NA | 6.28 [5.90, 6.89] | 93.4 |
| **Wall thickness segment 11 (median [IQR])** | 5.62 [5.09, 6.32] | 5.57 [5.19, 5.95] | 5.62 [5.10, 6.32] | 93.2 | 5.56 [5.07, 6.08] | NA | 5.56 [5.07, 6.08] | 93.4 |
| **Wall thickness segment 12 (median [IQR])** | 5.60 [5.22, 6.25] | 5.56 [5.53, 5.60] | 5.60 [5.22, 6.26] | 93.2 | 5.50 [5.14, 6.02] | NA | 5.50 [5.14, 6.02] | 93.4 |
| **Wall thickness segment 13 (median [IQR])** | 5.48 [5.10, 5.91] | 8.01 [7.03, 9.00] | 5.48 [5.10, 5.91] | 93.2 | 5.44 [5.13, 5.91] | NA | 5.44 [5.13, 5.91] | 93.4 |
| **Wall thickness segment 14 (median [IQR])** | 6.00 [5.36, 6.66] | 8.33 [6.87, 9.79] | 6.00 [5.36, 6.66] | 93.2 | 5.90 [5.44, 6.28] | NA | 5.90 [5.44, 6.28] | 93.4 |
| **Wall thickness segment 15 (median [IQR])** | 5.09 [4.48, 5.69] | 7.21 [5.80, 8.63] | 5.09 [4.48, 5.69] | 93.2 | 4.99 [4.56, 5.40] | NA | 4.99 [4.56, 5.40] | 93.4 |
| **Wall thickness segment 16 (median [IQR])** | 5.26 [4.80, 5.72] | 8.31 [6.87, 9.75] | 5.26 [4.80, 5.72] | 93.2 | 5.13 [4.82, 5.67] | NA | 5.13 [4.82, 5.67] | 93.4 |
| **Global wall thickness (median [IQR])** | 6.30 [5.73, 6.84] | 7.33 [7.28, 7.38] | 6.30 [5.72, 6.84] | 93.2 | 6.17 [5.90, 6.59] | NA | 6.17 [5.90, 6.59] | 93.4 |
| **Septal wall thickness (median [IQR])** | 6.68 [5.88, 7.35] | 8.28 [7.88, 8.69] | 6.67 [5.88, 7.34] | 93.2 | 6.37 [6.00, 6.90] | NA | 6.37 [6.00, 6.90] | 93.4 |
| **Maximum wall thickness (median [IQR])** | 8.10 [7.24, 9.02] | 10.96 [10.29, 11.64] | 8.09 [7.24, 9.01] | 93.2 | 8.01 [7.43, 8.66] | NA | 8.01 [7.43, 8.66] | 93.4 |

Abbreviations:
ARVC: arrythmogenic right ventricular cardiomyopathy; BMI: body mass index; CMR: cardiac magnetic resonance imaging; DCM: dilated cardiomyopathy; ECG: Electrocardiography; EDVi: indexed end-diastolic volume; EDMi: indexed end-diastolic mass; EF: ejection fraction; ESVi: indexed end-systolic volume; G+: carriers of likely pathogenic and pathogenic variants associated with one of the cardiomyopathies; HCM: hypertrophic cardiomyopathy; HDL: high-density lipoprotein; IQR: interquartile range; LDL: low-density lipoprotein; LV: left ventricular; MET: metabolic equivalent of task; MVR: mass to volume ratio; PAFR: peak atrial filling rate; peakEcc: peak circumferential strain; peakEll2Ch: longitudinal strain analyzed in 2-chamber view; peakEll4Ch: longitudinal strain analyzed in 4-chamber view; PER: peak ejection rate; PFR: peak filling rate; RV: right ventricular; SVi: indexed stroke volume; TPKEcc: global time to peak circumferential strain; TPKEll2Ch: global time to longitudinal strain analyzed in 2-chamber view; TPKEll4Ch: global time to longitudinal strain analyzed in 4-chamber view.

**Supplementary Table III: Extensive baseline table**

| | Controls G- | | | | HCM G+ | | | |
|---|---|---|---|---|---|---|---|---|
| | Overall | Diagnosed | Non-Diagnosed | Missing | Overall | Diagnosed | Non-Diagnosed | Missing |
| n (%) | 9,972 (100) | 87 (0.8) | 9,885 (99.2) | | 1,346 (100) | 35 (2.6) | 1,311 (97.4) | |
| **Sex = Female (%)** | 5,436 (54.5) | 35 (40.2) | 5,401 (54.6) | 0 | 720 (53.5) | 20 (57.1) | 700 (53.4) | 0 |
| **Age (median [IQR])** | 57.00 [49.00, 63.00] | 62.00 [56.00, 66.00] | 57.00 [49.00, 63.00] | 0 | 56.00 [49.00, 63.00] | 59.00 [51.00, 66.00] | 56.00 [49.00, 63.00] | 0 |
| **Ethnicity (%)** | | | | 1 | | | | 1.3 |
| Asian | 1,076 (10.9) | 5 (5.8) | 1071 (10.9) | | 251 (18.9) | 3 (8.6) | 248 (19.2) | |
| Black | 164 (1.7) | 3 (3.5) | 161 (1.6) | | 22 (1.7) | 0 (0.0) | 22 (1.7) | |
| Chinese | 56 (0.6) | 1 (1.2) | 55 (0.6) | | 1 (0.1) | 0 (0.0) | 1 (0.1) | |
| Mixed | 132 (1.3) | 2 (2.3) | 130 (1.3) | | 28 (2.1) | 0 (0.0) | 28 (2.2) | |
| Other | 160 (1.6) | 1 (1.2) | 159 (1.6) | | 26 (2.0) | 0 (0.0) | 26 (2.0) | |
| White | 8,288 (83.9) | 74 (86.0) | 8,214 (83.9) | | 1001 (75.3) | 32 (91.4) | 969 (74.9) | |
| **CARDIOVASCULAR RISK FACTORS** | | | | | | | | |
| **BMI (median [IQR])** | 26.73 [24.15, 29.82] | 28.83 [25.33, 32.20] | 26.71 [24.15, 29.80] | 0.5 | 26.56 [23.88, 29.76] | 26.54 [23.30, 31.10] | 26.56 [23.90, 29.72] | 1 |
| **Diabetes (%)** | 914 (9.2) | 21 (24.1) | 893 (9.0) | 0 | 154 (11.4) | 4 (11.4) | 150 (11.4) | 0 |
| **Hypertension (%)** | 3,420 (34.3) | 60 (69.0) | 3,360 (34.0) | 0 | 475 (35.3) | 23 (65.7) | 452 (34.5) | 0 |
| **Mean systolic blood pressure (median [IQR])** | 135.50 [124.00, 149.00] | 144.00 [130.00, 155.50] | 135.50 [124.00, 149.00] | 0.1 | 135.00 [123.50, 148.50] | 135.50 [125.00, 154.25] | 135.00 [123.50, 148.00] | 0.2 |
| **Mean diastolic blood pressure (median [IQR])** | 82.00 [75.00, 88.50] | 84.00 [76.50, 91.50] | 81.50 [75.00, 88.50] | 0.1 | 81.50 [75.50, 89.00] | 81.50 [72.50, 91.25] | 81.50 [75.50, 89.00] | 0.2 |
| **Hypercholesterolaemia (%)** | 2,416 (24.2) | 34 (39.1) | 2,382 (24.1) | 0 | 369 (27.4) | 12 (34.3) | 357 (27.2) | 0 |
| **Total cholesterol (median [IQR])** | 5.61 [4.86, 6.38] | 5.23 [4.39, 6.10] | 5.61 [4.87, 6.38] | 4.3 | 5.60 [4.80, 6.38] | 5.58 [4.73, 6.46] | 5.60 [4.80, 6.37] | 4.5 |
| **HDL (median [IQR])** | 1.38 [1.16, 1.65] | 1.29 [1.13, 1.56] | 1.38 [1.16, 1.65] | 11.3 | 1.37 [1.14, 1.63] | 1.32 [1.23, 1.58] | 1.37 [1.14, 1.63] | 12.1 |
| **LDL (median [IQR])** | 3.50 [2.92, 4.09] | 3.22 [2.54, 3.81] | 3.50 [2.92, 4.09] | 4.6 | 3.48 [2.87, 4.09] | 3.38 [2.67, 4.43] | 3.48 [2.88, 4.07] | 4.8 |
| **Ever Smoked (%)** | 4,132 (41.4) | 50 (57.5) | 4,082 (41.3) | 0 | 543 (40.3) | 12 (34.3) | 531 (40.5) | 0 |
| **MET minutes per week for walking (median [IQR])** | 693.00 [297.00, 1,386.00] | 528.00 [255.75, 1,608.75] | 693.00 [297.00, 1,386.00] | 19.8 | 693.00 [307.72, 1,386.00] | 495.00 [198.00, 1,386.00] | 693.00 [330.00, 1,386.00] | 19.6 |
| **MET minutes per week for moderate activity (median [IQR])** | 480.00 [120.00, 1,200.00] | 480.00 [100.00, 1,680.00] | 480.00 [120.00, 1,200.00] | 19.8 | 480.00 [120.00, 1,200.00] | 400.00 [160.00, 840.00] | 480.00 [120.00, 1,200.00] | 19.6 |
| **MET minutes per week for vigorous activity (median [IQR])** | 240.00 [0.00, 960.00] | 0.00 [0.00, 820.00] | 240.00 [0.00, 960.00] | 19.8 | 160.00 [0.00, 930.00] | 240.00 [0.00, 480.00] | 160.00 [0.00, 960.00] | 19.6 |
| **Total MET minutes per week (median [IQR])** | 1,773.00 [810.00, 3,452.50] | 1,367.50 [478.50, 3,834.00] | 1,776.50 [810.00, 3,450.00] | 19.8 | 1,762.00 [848.12, 3,490.25] | 1,253.00 [693.00, 3,426.00] | 1,773.00 [853.00, 3,492.00] | 19.6 |
| **Family heart disease (%)** | 4,458 (44.7) | 34 (39.1) | 4,424 (44.8) | 0 | 623 (46.3) | 15 (42.9) | 608 (46.4) | 0 |
| **CARDIAC DISEASES/OUTCOMES** | | | | | | | | |
| **Cardiac problem (%)** | 41 (0.4) | 2 (2.3) | 39 (0.4) | 0 | 5 (0.4) | 0 (0.0) | 5 (0.4) | 0 |
| **Heart failure (%)** | 182 (1.8) | 74 (85.1) | 108 (1.1) | 0 | 33 (2.5) | 15 (42.9) | 18 (1.4) | 0 |
| **Cardiomyopathy (%)** | 37 (0.4) | 26 (29.9) | 11 (0.1) | 0 | 27 (2.0) | 21 (60.0) | 6 (0.5) | 0 |
| **Dilated cardiomyopathy (%)** | 14 (0.1) | 8 (9.2) | 6 (0.1) | 0 | 1 (0.1) | 1 (2.9) | 0 (0.0) | 0 |
| **Hypertrophic cardiomyopathy (%)** | 8 (0.1) | 7 (8.0) | 1 (0.0) | 0 | 20 (1.5) | 14 (40.0) | 6 (0.5) | 0 |
| **Ventricular arrhythmias (%)** | 33 (0.3) | 3 (3.4) | 30 (0.3) | 0 | 8 (0.6) | 4 (11.4) | 4 (0.3) | 0 |
| **Atrial arrhythmias (%)** | 191 (1.9) | 19 (21.8) | 172 (1.7) | 0 | 32 (2.4) | 6 (17.1) | 26 (2.0) | 0 |
| **Heart arrhythmia (%)** | 54 (0.5) | 2 (2.3) | 52 (0.5) | 0 | 4 (0.3) | 0 (0.0) | 4 (0.3) | 0 |
| **Chronic ischemic heart disease (%)** | 725 (7.3) | 0 (0.0) | 725 (7.3) | 0 | 93 (6.9) | 0 (0.0) | 93 (7.1) | 0 |
| **Acute myocardial infarction (%)** | 298 (3.0) | 1 (1.1) | 297 (3.0) | 0 | 36 (2.7) | 2 (5.7) | 34 (2.6) | 0 |
| **Cardiac arrest (%)** | 34 (0.3) | 1 (1.1) | 33 (0.3) | 0 | 5 (0.4) | 0 (0.0) | 5 (0.4) | 0 |
| **Angina pectoris (%)** | 312 (3.1) | 2 (2.3) | 310 (3.1) | 0 | 56 (4.2) | 0 (0.0) | 56 (4.3) | 0 |
| **Conduction disorders (%)** | 151 (1.5) | 10 (11.5) | 141 (1.4) | 0 | 26 (1.9) | 6 (17.1) | 20 (1.5) | 0 |
| **Valvular disease (%)** | 241 (2.4) | 23 (26.4) | 218 (2.2) | 0 | 41 (3.0) | 8 (22.9) | 33 (2.5) | 0 |
| **Congenital heart disease (%)** | 28 (0.3) | 3 (3.4) | 25 (0.3) | 0 | 4 (0.3) | 0 (0.0) | 4 (0.3) | 0 |
| **Pulmonary obstructive disease (%)** | 494 (5.0) | 24 (27.6) | 470 (4.8) | 0 | 57 (4.2) | 4 (11.4) | 53 (4.0) | 0 |
| **Cardiovascular death (%)** | 181 (1.8) | 13 (14.9) | 168 (1.7) | 0 | 18 (1.3) | 3 (8.6) | 15 (1.1) | 0 |
| **All-cause mortality (%)** | 513 (5.1) | 27 (31.0) | 486 (4.9) | 0 | 62 (4.6) | 6 (17.1) | 56 (4.3) | 0 |
| **ECG MEASUREMENTS** | | | | | | | | |
| **n (%)** | 1,062 (10.6) | 4 (4.6) | 1,058 (10.7) | | 138 (10.3) | 5 (14.3) | 133 (10.1) | |
| **P duration (median [IQR])** | 100.00 [90.00, 108.00] | 90.00 [82.00, 111.00] | 100.00 [90.00, 108.00] | 89.8 | 100.00 [90.00, 108.00] | 100.00 [86.00, 102.00] | 100.00 [90.00, 108.00] | 90 |
| **P axis (median [IQR])** | 55.00 [40.25, 67.00] | 47.00 [47.00, 47.00] | 55.00 [40.00, 67.00] | 93.1 | 53.00 [36.00, 63.00] | 70.00 [66.00, 70.00] | 52.00 [36.00, 62.00] | 92.9 |
| **PQ interval (median [IQR])** | 160.00 [145.50, 178.00] | 188.00 [188.00, 188.00] | 160.00 [145.00, 178.00] | 93.1 | 161.00 [147.50, 172.50] | 190.00 [186.00, 199.00] | 160.00 [146.00, 172.00] | 92.9 |
| **QRS duration (median [IQR])** | 86.00 [80.00, 94.00] | 93.00 [89.50, 97.00] | 86.00 [80.00, 94.00] | 89.4 | 84.00 [80.00, 93.50] | 100.00 [82.00, 104.00] | 84.00 [80.00, 92.00] | 89.7 |
| **R axis (median [IQR])** | 34.00 [7.00, 58.00] | -48.00 [-48.00, -48.00] | 34.00 [7.50, 58.00] | 92.9 | 39.00 [10.00, 52.75] | 2.00 [-17.00, 24.00] | 39.00 [11.50, 53.50] | 92.7 |
| **QTC interval (median [IQR])** | 417.00 [402.00, 433.00] | 511.00 [511.00, 511.00] | 417.00 [402.00, 432.50] | 92.9 | 414.50 [402.00, 429.00] | 435.00 [428.50, 456.50] | 414.00 [402.00, 428.50] | 92.7 |
| **T axis (median [IQR])** | 40.00 [23.00, 55.25] | 92.00 [92.00, 92.00] | 40.00 [23.00, 55.00] | 92.9 | 45.00 [31.25, 60.75] | 49.00 [43.00, 96.00] | 45.00 [30.50, 60.50] | 92.7 |
| **CMR MEASUREMENTS** | | | | | | | | |
| **n (%)** | 990 (9.9) | 4 (4.6) | 986 (10.0) | | 134 (10.0) | 4 (11.4) | 130 (9.9) | |
| **RVEDVi (median [IQR])** | 80.18 [70.62, 90.27] | 79.28 [79.28, 79.28] | 80.19 [70.61, 90.27] | 91 | 77.30 [67.73, 90.71] | 79.56 [77.34, 90.20] | 77.12 [67.27, 90.71] | 90.4 |
| **RVESVi (median [IQR])** | 32.89 [27.38, 39.68] | 20.93 [20.93, 20.93] | 32.90 [27.42, 39.70] | 91 | 31.72 [26.22, 37.33] | 34.80 [30.85, 43.04] | 31.40 [26.20, 37.28] | 90.4 |
| **RVSVi (median [IQR])** | 46.55 [40.92, 52.84] | 58.35 [58.35, 58.35] | 46.53 [40.91, 52.81] | 91 | 46.03 [40.64, 54.04] | 47.41 [45.02, 51.27] | 45.77 [40.64, 54.04] | 90.4 |
| **RVEF (median [IQR])** | 58.38 [54.19, 62.76] | 73.60 [73.60, 73.60] | 58.37 [54.19, 62.74] | 91 | 59.56 [54.73, 63.95] | 56.21 [51.68, 60.89] | 59.56 [54.80, 63.99] | 90.5 |
| **RVPER (median [IQR])** | 388.72 [316.56, 465.82] | 446.55 [446.55, 446.55] | 388.66 [316.54, 465.97] | 91 | 389.50 [310.19, 475.79] | 391.94 [296.40, 448.92] | 389.50 [310.19, 475.79] | 90.4 |
| **RVPFR (median [IQR])** | 300.61 [245.17, 364.00] | 373.24 [373.24, 373.24] | 300.34 [245.12, 363.44] | 91 | 285.88 [232.85, 336.38] | 363.12 [334.18, 386.47] | 278.68 [230.83, 334.67] | 90.4 |
| **RVPAFR (median [IQR])** | 282.95 [222.68, 360.32] | 547.72 [547.72, 547.72] | 282.86 [222.56, 360.07] | 91 | 299.05 [236.43, 365.16] | 263.66 [225.50, 297.24] | 300.15 [236.43, 366.42] | 90.4 |
| **LVEDVi (median [IQR])** | 74.33 [66.34, 83.11] | 64.68 [59.39, 69.33] | 74.37 [66.38, 83.15] | 91.9 | 74.50 [64.57, 84.89] | 85.13 [83.78, 87.08] | 72.35 [64.32, 84.59] | 91.2 |
| **LVESVi (median [IQR])** | 30.02 [25.12, 35.70] | 26.43 [25.24, 28.39] | 30.02 [25.13, 35.72] | 91.9 | 30.31 [24.10, 35.20] | 43.58 [39.30, 47.19] | 29.37 [24.09, 34.83] | 91.2 |
| **LVSVi (median [IQR])** | 44.03 [39.34, 50.28] | 38.25 [31.00, 44.09] | 44.05 [39.37, 50.30] | 91.9 | 44.17 [38.21, 49.98] | 44.56 [42.26, 45.10] | 44.07 [38.21, 50.19] | 91.2 |
| **LVEF (median [IQR])** | 59.47 [55.29, 63.52] | 59.14 [51.51, 63.31] | 59.48 [55.29, 63.52] | 91.9 | 59.55 [55.96, 63.53] | 51.02 [47.77, 53.26] | 59.73 [56.25, 63.62] | 91.2 |
| **LVPER (median [IQR])** | 373.80 [302.29, 452.71] | 284.40 [256.83, 364.56] | 373.81 [302.44, 453.27] | 91.9 | 340.46 [266.40, 459.49] | 350.17 [311.46, 380.66] | 340.46 [264.60, 460.79] | 91.2 |
| **LVPFR (median [IQR])** | 321.31 [259.23, 385.16] | 201.74 [189.85, 229.92] | 321.49 [259.95, 385.63] | 91.9 | 320.26 [252.40, 371.17] | 315.79 [273.86, 360.98] | 320.26 [248.52, 371.17] | 91.2 |
| **LVPAFR (median [IQR])** | 233.66 [167.35, 306.30] | 363.87 [208.97, 466.86] | 233.50 [167.46, 305.08] | 91.9 | 241.27 [165.31, 316.89] | 213.93 [135.78, 272.60] | 241.27 [167.29, 321.53] | 91.2 |
| **LVEDMi (median [IQR])** | 41.88 [36.52, 48.62] | 45.76 [34.02, 48.75] | 41.85 [36.55, 48.61] | 91.9 | 42.87 [35.12, 49.47] | 49.63 [46.84, 53.17] | 42.41 [34.89, 49.27] | 91.2 |
| **LVMVR (median [IQR])** | 0.56 [0.50, 0.62] | 0.70 [0.56, 0.70] | 0.56 [0.50, 0.62] | 91.9 | 0.56 [0.50, 0.64] | 0.58 [0.55, 0.61] | 0.56 [0.50, 0.64] | 91.2 |
| **LVEDV/RVEDV (median [IQR])** | 0.93 [0.86, 1.03] | 0.93 [0.93, 0.93] | 0.93 [0.86, 1.03] | 92.2 | 0.94 [0.86, 1.03] | 1.07 [0.99, 1.08] | 0.94 [0.86, 0.99] | 91.3 |
| **LVESV/RVESV (median [IQR])** | 0.91 [0.80, 1.04] | 1.15 [1.15, 1.15] | 0.91 [0.80, 1.04] | 92.2 | 0.90 [0.82, 1.05] | 1.28 [1.14, 1.30] | 0.90 [0.82, 1.02] | 91.3 |
| **peakEcc (median [IQR])** | -22.70 [-24.98, -20.43] | -21.60 [-21.61, -21.59] | -22.72 [-24.98, -20.42] | 93.7 | -22.82 [-24.98, -20.60] | -20.32 [-20.51, -20.27] | -22.91 [-25.19, -20.88] | 93.5 |
| **TPKEcc (median [IQR])** | 331.31 [309.83, 354.66] | 325.97 [321.27, 330.67] | 331.31 [309.75, 354.68] | 93.7 | 332.34 [308.14, 354.02] | 347.73 [320.45, 364.22] | 331.96 [308.15, 353.59] | 93.5 |
| **peakEll2Ch (median [IQR])** | -21.19 [-23.34, -18.93] | -23.89 [-23.89, -23.89] | -21.17 [-23.32, -18.93] | 93.8 | -20.99 [-23.36, -19.01] | -20.63 [-20.99, -20.19] | -21.54 [-23.50, -18.88] | 93.5 |
| **TPKEll2Ch (median [IQR])** | 349.86 [321.84, 379.10] | 388.50 [388.50, 388.50] | 349.86 [321.81, 379.08] | 93.9 | 355.54 [320.85, 381.68] | 403.20 [386.90, 420.80] | 353.10 [320.40, 379.60] | 93.5 |
| **peakEll4Ch (median [IQR])** | -23.30 [-25.97, -21.37] | -21.40 [-21.40, -21.40] | -23.30 [-25.99, -21.37] | 93.9 | -23.82 [-26.50, -21.90] | -20.87 [-22.20, -19.79] | -24.05 [-26.93, -22.28] | 94.1 |
| **TPKEll4Ch (median [IQR])** | 357.53 [327.07, 397.64] | 360.78 [360.78, 360.78] | 357.30 [326.96, 397.81] | 93.9 | 350.13 [318.54, 395.01] | 403.22 [378.52, 407.61] | 349.20 [318.54, 390.16] | 94.1 |
| **Wall thickness segment 1 (median [IQR])** | 7.65 [6.81, 8.50] | 7.04 [5.94, 8.14] | 7.65 [6.81, 8.49] | 93.2 | 7.59 [6.74, 8.63] | 9.35 [9.26, 9.44] | 7.59 [6.72, 8.55] | 92.9 |
| **Wall thickness segment 2 (median [IQR])** | 6.75 [5.75, 7.91] | 7.50 [6.70, 8.31] | 6.75 [5.74, 7.90] | 93.2 | 7.10 [5.91, 8.26] | 9.50 [8.42, 10.57] | 7.06 [5.87, 8.21] | 92.9 |
| **Wall thickness segment 3 (median [IQR])** | 6.05 [5.17, 6.96] | 7.60 [6.78, 8.42] | 6.05 [5.17, 6.95] | 93.2 | 6.23 [5.07, 7.49] | 7.99 [7.50, 8.47] | 6.22 [5.04, 7.39] | 92.9 |
| **Wall thickness segment 4 (median [IQR])** | 6.54 [5.82, 7.22] | 8.31 [7.66, 8.97] | 6.54 [5.81, 7.21] | 93.2 | 6.49 [5.86, 6.99] | 7.25 [7.12, 7.38] | 6.47 [5.85, 6.92] | 92.9 |
| **Wall thickness segment 5 (median [IQR])** | 6.20 [5.62, 6.96] | 6.33 [5.27, 7.39] | 6.20 [5.62, 6.96] | 93.2 | 6.03 [5.57, 6.65] | 6.36 [6.00, 6.72] | 6.03 [5.57, 6.64] | 92.9 |
| **Wall thickness segment 6 (median [IQR])** | 6.55 [5.97, 7.31] | 6.61 [5.42, 7.79] | 6.55 [5.98, 7.31] | 93.2 | 6.56 [5.94, 7.13] | 6.72 [6.45, 6.98] | 6.56 [5.93, 7.09] | 92.9 |
| **Wall thickness segment 7 (median [IQR])** | 5.73 [5.28, 6.31] | 5.48 [4.90, 6.07] | 5.73 [5.29, 6.31] | 93.2 | 5.75 [5.22, 6.20] | 6.03 [5.92, 6.13] | 5.75 [5.21, 6.19] | 92.9 |
| **Wall thickness segment 8 (median [IQR])** | 7.01 [6.28, 7.76] | 8.52 [7.77, 9.27] | 7.01 [6.28, 7.75] | 93.2 | 6.89 [6.17, 7.50] | 8.76 [7.66, 9.86] | 6.89 [6.16, 7.49] | 92.9 |
| **Wall thickness segment 9 (median [IQR])** | 7.38 [6.48, 8.25] | 9.47 [8.05, 10.89] | 7.38 [6.48, 8.25] | 93.2 | 7.21 [6.38, 8.22] | 8.99 [7.76, 10.21] | 7.21 [6.38, 8.20] | 92.9 |
| **Wall thickness segment 10 (median [IQR])** | 6.23 [5.59, 6.96] | 7.07 [6.08, 8.06] | 6.23 [5.60, 6.96] | 93.2 | 6.18 [5.45, 7.05] | 6.48 [6.17, 6.78] | 6.18 [5.44, 7.00] | 92.9 |
| **Wall thickness segment 11 (median [IQR])** | 5.62 [5.09, 6.32] | 5.57 [5.19, 5.95] | 5.62 [5.10, 6.32] | 93.2 | 5.53 [5.04, 6.33] | 5.75 [5.62, 5.88] | 5.53 [5.02, 6.37] | 92.9 |
| **Wall thickness segment 12 (median [IQR])** | 5.60 [5.22, 6.25] | 5.56 [5.53, 5.60] | 5.60 [5.22, 6.26] | 93.2 | 5.59 [5.10, 6.11] | 5.52 [5.32, 5.73] | 5.59 [5.08, 6.13] | 92.9 |
| **Wall thickness segment 13 (median [IQR])** | 5.48 [5.10, 5.91] | 8.01 [7.03, 9.00] | 5.48 [5.10, 5.91] | 93.2 | 5.49 [5.10, 5.94] | 5.21 [4.61, 5.82] | 5.49 [5.12, 5.93] | 92.9 |
| **Wall thickness segment 14 (median [IQR])** | 6.00 [5.36, 6.66] | 8.33 [6.87, 9.79] | 6.00 [5.36, 6.66] | 93.2 | 5.95 [5.38, 6.60] | 6.05 [5.36, 6.74] | 5.95 [5.38, 6.58] | 92.9 |
| **Wall thickness segment 15 (median [IQR])** | 5.09 [4.48, 5.69] | 7.21 [5.80, 8.63] | 5.09 [4.48, 5.69] | 93.2 | 4.99 [4.45, 5.69] | 5.30 [5.05, 5.54] | 4.99 [4.44, 5.68] | 92.9 |
| **Wall thickness segment 16 (median [IQR])** | 5.26 [4.80, 5.72] | 8.31 [6.87, 9.75] | 5.26 [4.80, 5.72] | 93.2 | 5.23 [4.75, 5.68] | 5.32 [4.77, 5.87] | 5.23 [4.76, 5.65] | 92.9 |
| **Global wall thickness (median [IQR])** | 6.30 [5.73, 6.84] | 7.33 [7.28, 7.38] | 6.30 [5.72, 6.84] | 93.2 | 6.30 [5.74, 6.92] | 7.06 [6.95, 7.17] | 6.28 [5.70, 6.87] | 92.9 |
| **Septal wall thickness (median [IQR])** | 6.68 [5.88, 7.35] | 8.28 [7.88, 8.69] | 6.67 [5.88, 7.34] | 93.2 | 6.65 [6.13, 7.61] | 8.26 [7.96, 8.55] | 6.62 [6.12, 7.52] | 92.9 |
| **Maximum wall thickness (median [IQR])** | 8.10 [7.24, 9.02] | 10.96 [10.29, 11.64] | 8.09 [7.24, 9.01] | 93.2 | 8.19 [7.46, 9.65] | 11.54 [11.49, 11.59] | 8.16 [7.45, 9.58] | 92.9 |

Abbreviations:
ARVC: arrythmogenic right ventricular cardiomyopathy; BMI: body mass index; CMR: cardiac magnetic resonance imaging; DCM: dilated cardiomyopathy; ECG: Electrocardiography; EDVi: indexed end-diastolic volume; EDMi: indexed end-diastolic mass; EF: ejection fraction; ESVi: indexed end-systolic volume; G+: carriers of likely pathogenic and pathogenic variants associated with one of the cardiomyopathies; HCM: hypertrophic cardiomyopathy; HDL: high-density lipoprotein; IQR: interquartile range; LDL: low-density lipoprotein; LV: left ventricular; MET: metabolic equivalent of task; MVR: mass to volume ratio; PAFR: peak atrial filling rate; peakEcc: peak circumferential strain; peakEll2Ch: longitudinal strain analyzed in 2-chamber view; peakEll4Ch: longitudinal strain analyzed in 4-chamber view; PER: peak ejection rate; PFR: peak filling rate; RV: right ventricular; SVi: indexed stroke volume; TPKEcc: global time to peak circumferential strain; TPKEll2Ch: global time to longitudinal strain analyzed in 2-chamber view; TPKEll4Ch: global time to longitudinal strain analyzed in 4-chamber view.

**Supplementary Table III: Extensive baseline table**

| | Controls G- | | | | strict HCM G+ | | | |
|---|---|---|---|---|---|---|---|---|
| | **Overall** | **Diagnosed** | **Non-Diagnosed** | **Missing** | **Overall** | **Diagnosed** | **Non-Diagnosed** | **Missing** |
| n (%) | 9,972 (100) | 87 (0.8) | 9,885 (99.2) | | 801 (100) | 32 (4.0) | 769 (96.0) | |
| **Sex = Female (%)** | 5,436 (54.5) | 35 (40.2) | 5,401 (54.6) | 0 | 445 (55.6) | 19 (59.4) | 426 (55.4) | 0 |
| **Age (median [IQR])** | 57.00 [49.00, 63.00] | 62.00 [56.00, 66.00] | 57.00 [49.00, 63.00] | 0 | 58.00 [50.00, 63.00] | 60.00 [52.75, 66.00] | 58.00 [50.00, 63.00] | 0 |
| **Ethnicity (%)** | | | | 1 | | | | 0.6 |
| Asian | 1,076 (10.9) | 5 (5.8) | 1071 (10.9) | | 14 (1.8) | 0 (0.0) | 14 (1.8) | |
| Black | 164 (1.7) | 3 (3.5) | 161 (1.6) | | 18 (2.3) | 0 (0.0) | 18 (2.4) | |
| Chinese | 56 (0.6) | 1 (1.2) | 55 (0.6) | | 1 (0.1) | 0 (0.0) | 1 (0.1) | |
| Mixed | 132 (1.3) | 2 (2.3) | 130 (1.3) | | 6 (0.8) | 0 (0.0) | 6 (0.8) | |
| Other | 160 (1.6) | 1 (1.2) | 159 (1.6) | | 3 (0.4) | 0 (0.0) | 3 (0.4) | |
| White | 8,288 (83.9) | 74 (86.0) | 8,214 (83.9) | | 754 (94.7) | 32 (100.0) | 722 (94.5) | |
| CARDIOVASCULAR RISK FACTORS | | | | | | | | |
| **BMI (median [IQR])** | 26.73 [24.15, 29.82] | 28.83 [25.33, 32.20] | 26.71 [24.15, 29.80] | 0.5 | 26.56 [23.84, 29.85] | 26.25 [23.37, 30.57] | 26.58 [23.85, 29.79] | 0.4 |
| **Diabetes (%)** | 914 (9.2) | 21 (24.1) | 893 (9.0) | 0 | 60 (7.5) | 3 (9.4) | 57 (7.4) | 0 |
| **Hypertension (%)** | 3,420 (34.3) | 60 (69.0) | 3,360 (34.0) | 0 | 291 (36.3) | 21 (65.6) | 270 (35.1) | 0 |
| **Mean systolic blood pressure (median [IQR])** | 135.50 [124.00, 149.00] | 144.00 [130.00, 155.50] | 135.50 [124.00, 149.00] | 0.1 | 136.00 [123.50, 150.50] | 138.50 [128.12, 156.50] | 136.00 [123.50, 150.00] | 0.1 |
| **Mean diastolic blood pressure (median [IQR])** | 82.00 [75.00, 88.50] | 84.00 [76.50, 91.50] | 81.50 [75.00, 88.50] | 0.1 | 81.50 [75.00, 89.00] | 83.00 [74.88, 92.12] | 81.50 [75.00, 89.00] | 0.1 |
| **Hypercholesterolaemia (%)** | 2,416 (24.2) | 34 (39.1) | 2,382 (24.1) | 0 | 195 (24.3) | 11 (34.4) | 184 (23.9) | 0 |
| **Total cholesterol (median [IQR])** | 5.61 [4.86, 6.38] | 5.23 [4.39, 6.10] | 5.61 [4.87, 6.38] | 4.3 | 5.63 [4.86, 6.42] | 5.75 [5.03, 6.51] | 5.63 [4.85, 6.41] | 4.6 |
| **HDL (median [IQR])** | 1.38 [1.16, 1.65] | 1.29 [1.13, 1.56] | 1.38 [1.16, 1.65] | 11.3 | 1.40 [1.18, 1.68] | 1.33 [1.24, 1.64] | 1.40 [1.18, 1.68] | 13 |
| **LDL (median [IQR])** | 3.50 [2.92, 4.09] | 3.22 [2.54, 3.81] | 3.50 [2.92, 4.09] | 4.6 | 3.50 [2.90, 4.11] | 3.47 [2.98, 4.46] | 3.50 [2.90, 4.10] | 4.9 |
| **Ever Smoked (%)** | 4,132 (41.4) | 50 (57.5) | 4,082 (41.3) | 0 | 361 (45.1) | 11 (34.4) | 350 (45.5) | 0 |
| **MET minutes per week for walking (median [IQR])** | 693.00 [297.00, 1,386.00] | 528.00 [255.75, 1,608.75] | 693.00 [297.00, 1,386.00] | 19.8 | 693.00 [330.00, 1,386.00] | 495.00 [214.50, 1,386.00] | 693.00 [330.00, 1,386.00] | 18 |
| **MET minutes per week for moderate activity (median [IQR])** | 480.00 [120.00, 1,200.00] | 480.00 [100.00, 1,680.00] | 480.00 [120.00, 1,200.00] | 19.8 | 480.00 [160.00, 1,200.00] | 400.00 [140.00, 900.00] | 480.00 [160.00, 1,290.00] | 18 |
| **MET minutes per week for vigorous activity (median [IQR])** | 240.00 [0.00, 960.00] | 0.00 [0.00, 820.00] | 240.00 [0.00, 960.00] | 19.8 | 240.00 [0.00, 960.00] | 240.00 [0.00, 540.00] | 240.00 [0.00, 960.00] | 18 |
| **Total MET minutes per week (median [IQR])** | 1,773.00 [810.00, 3,452.50] | 1,367.50 [478.50, 3,834.00] | 1,776.50 [810.00, 3,450.00] | 19.8 | 1,895.00 [924.00, 3,626.00] | 1,253.00 [711.50, 3,606.00] | 1,942.50 [925.50, 3,622.50] | 18 |
| **Family heart disease (%)** | 4,458 (44.7) | 34 (39.1) | 4,424 (44.8) | 0 | 389 (48.6) | 14 (43.8) | 375 (48.8) | 0 |
| CARDIAC DISEASES/OUTCOMES | | | | | | | | |
| **Cardiac problem (%)** | 41 (0.4) | 2 (2.3) | 39 (0.4) | 0 | 4 (0.5) | 0 (0.0) | 4 (0.5) | 0 |
| **Heart failure (%)** | 182 (1.8) | 74 (85.1) | 108 (1.1) | 0 | 25 (3.1) | 13 (40.6) | 12 (1.6) | 0 |
| **Cardiomyopathy (%)** | 37 (0.4) | 26 (29.9) | 11 (0.1) | 0 | 25 (3.1) | 20 (62.5) | 5 (0.7) | 0 |
| **Dilated cardiomyopathy (%)** | 14 (0.1) | 8 (9.2) | 6 (0.1) | 0 | 0 (0.0) | 0 (0.0) | 0 (0.0) | 0 |
| **Hypertrophic cardiomyopathy (%)** | 8 (0.1) | 7 (8.0) | 1 (0.0) | 0 | 19 (2.4) | 14 (43.8) | 5 (0.7) | 0 |
| **Ventricular arrhythmias (%)** | 33 (0.3) | 3 (3.4) | 30 (0.3) | 0 | 8 (1.0) | 4 (12.5) | 4 (0.5) | 0 |
| **Atrial arrhythmias (%)** | 191 (1.9) | 19 (21.8) | 172 (1.7) | 0 | 25 (3.1) | 6 (18.8) | 19 (2.5) | 0 |
| **Heart arrhythmia (%)** | 54 (0.5) | 2 (2.3) | 52 (0.5) | 0 | 52 (6.5) | 0 (0.0) | 52 (6.8) | 0 |
| **Chronic ischemic heart disease (%)** | 725 (7.3) | 0 (0.0) | 725 (7.3) | 0 | 22 (2.7) | 2 (6.2) | 20 (2.6) | 0 |
| **Acute myocardial infarction (%)** | 298 (3.0) | 1 (1.1) | 297 (3.0) | 0 | 4 (0.5) | 0 (0.0) | 4 (0.5) | 0 |
| **Cardiac arrest (%)** | 34 (0.3) | 1 (1.1) | 33 (0.3) | 0 | 15 (1.9) | 5 (15.6) | 10 (1.3) | 0 |
| **Angina pectoris (%)** | 312 (3.1) | 2 (2.3) | 310 (3.1) | 0 | 29 (3.6) | 8 (25.0) | 21 (2.7) | 0 |
| **Conduction disorders (%)** | 151 (1.5) | 10 (11.5) | 141 (1.4) | 0 | 3 (0.4) | 0 (0.0) | 3 (0.4) | 0 |
| **Valvular disease (%)** | 241 (2.4) | 23 (26.4) | 218 (2.2) | 0 | 36 (4.5) | 3 (9.4) | 33 (4.3) | 0 |
| **Congenital heart disease (%)** | 28 (0.3) | 3 (3.4) | 25 (0.3) | 0 | 3 (0.4) | 0 (0.0) | 3 (0.4) | 0 |
| **Pulmonary obstructive disease (%)** | 494 (5.0) | 24 (27.6) | 470 (4.8) | 0 | 32 (4.0) | 0 (0.0) | 32 (4.2) | 0 |
| **Cardiovascular death (%)** | 181 (1.8) | 13 (14.9) | 168 (1.7) | 0 | 10 (1.2) | 3 (9.4) | 7 (0.9) | 0 |
| **All-cause mortality (%)** | 513 (5.1) | 27 (31.0) | 486 (4.9) | 0 | 45 (5.6) | 5 (15.6) | 40 (5.2) | 0 |
| ECG MEASUREMENTS | | | | | | | | |
| **n (%)** | 1,062 (10.6) | 4 (4.6) | 1,058 (10.7) | | 85 (10.6) | 5 (15.6) | 80 (10.4) | 0 |
| **P duration (median [IQR])** | 100.00 [90.00, 108.00] | 90.00 [82.00, 111.00] | 100.00 [90.00, 108.00] | 89.8 | 100.00 [90.00, 106.00] | 100.00 [86.00, 102.00] | 100.00 [90.50, 106.00] | 89.6 |
| **P axis (median [IQR])** | 55.00 [40.25, 67.00] | 47.00 [47.00, 47.00] | 55.00 [40.00, 67.00] | 93.1 | 50.00 [36.00, 65.50] | 70.00 [66.00, 70.00] | 48.00 [35.75, 64.25] | 92.1 |
| **PQ interval (median [IQR])** | 160.00 [145.50, 178.00] | 188.00 [188.00, 188.00] | 160.00 [145.00, 178.00] | 93.1 | 164.00 [145.00, 174.00] | 190.00 [186.00, 199.00] | 163.00 [143.50, 172.00] | 92.1 |
| **QRS duration (median [IQR])** | 86.00 [80.00, 94.00] | 93.00 [89.50, 97.00] | 86.00 [80.00, 94.00] | 89.4 | 86.00 [80.00, 92.00] | 100.00 [82.00, 104.00] | 85.00 [80.00, 92.00] | 89.4 |
| **R axis (median [IQR])** | 34.00 [7.00, 58.00] | -48.00 [-48.00, -48.00] | 34.00 [7.50, 58.00] | 92.9 | 38.50 [9.75, 55.00] | 2.00 [-17.00, 24.00] | 39.00 [13.00, 55.00] | 92 |
| **QTC interval (median [IQR])** | 417.00 [402.00, 433.00] | 511.00 [511.00, 511.00] | 417.00 [402.00, 432.50] | 92.9 | 415.00 [401.75, 429.25] | 435.00 [428.50, 456.50] | 414.00 [401.00, 429.00] | 92 |
| **T axis (median [IQR])** | 40.00 [23.00, 55.25] | 92.00 [92.00, 92.00] | 40.00 [23.00, 55.00] | 92.9 | 45.00 [30.75, 61.25] | 49.00 [43.00, 96.00] | 45.00 [30.00, 61.00] | 92 |
| CMR MEASUREMENTS | | | | | | | | |
| **n (%)** | 990 (9.9) | 4 (4.6) | 986 (10.0) | | 84 (10.5) | 4 (12.5) | 80 (10.4) | 0 |
| **RVEDVi (median [IQR])** | 80.18 [70.62, 90.27] | 79.28 [79.28, 79.28] | 80.19 [70.61, 90.27] | 91 | 77.46 [68.84, 93.43] | 79.56 [77.34, 90.20] | 77.39 [67.84, 93.43] | 89.9 |
| **RVESVi (median [IQR])** | 32.89 [27.38, 39.68] | 20.93 [20.93, 20.93] | 32.90 [27.42, 39.70] | 91 | 31.40 [26.87, 37.04] | 34.80 [30.85, 43.04] | 31.35 [26.22, 36.21] | 89.9 |
| **RVSVi (median [IQR])** | 46.55 [40.92, 52.84] | 58.35 [58.35, 58.35] | 46.53 [40.91, 52.81] | 91 | 47.18 [41.46, 55.77] | 47.41 [45.02, 51.27] | 47.18 [41.46, 55.77] | 89.9 |
| **RVEF (median [IQR])** | 58.38 [54.19, 62.76] | 73.60 [73.60, 73.60] | 58.37 [54.19, 62.74] | 91 | 59.99 [56.00, 63.95] | 56.21 [51.67, 60.89] | 59.99 [56.46, 64.04] | 90 |
| **RVPER (median [IQR])** | 388.72 [316.56, 465.82] | 446.55 [446.55, 446.55] | 388.66 [316.54, 465.97] | 91 | 398.00 [333.00, 478.60] | 391.95 [296.40, 448.95] | 398.00 [333.00, 478.60] | 89.9 |
| **RVPFR (median [IQR])** | 300.61 [245.17, 364.00] | 373.24 [373.24, 373.24] | 300.34 [245.12, 363.44] | 91 | 294.17 [243.48, 338.50] | 363.15 [334.20, 386.48] | 278.70 [243.13, 334.41] | 89.9 |
| **RVPAFR (median [IQR])** | 282.95 [222.68, 360.32] | 547.72 [547.72, 547.72] | 282.86 [222.56, 360.07] | 91 | 302.92 [251.30, 369.30] | 263.65 [225.50, 297.22] | 307.70 [254.50, 378.30] | 89.9 |
| **LVEDVi (median [IQR])** | 74.33 [66.34, 83.11] | 64.68 [59.39, 69.33] | 74.37 [66.38, 83.15] | 91.9 | 78.88 [67.38, 87.06] | 85.13 [83.78, 87.08] | 77.40 [66.41, 87.06] | 90.9 |
| **LVESVi (median [IQR])** | 30.02 [25.12, 35.70] | 26.43 [25.24, 28.39] | 30.02 [25.13, 35.72] | 91.9 | 31.28 [25.63, 37.04] | 43.58 [39.30, 47.19] | 31.22 [25.36, 35.84] | 90.9 |
| **LVSVi (median [IQR])** | 44.03 [39.34, 50.28] | 38.25 [31.00, 44.09] | 44.05 [39.37, 50.30] | 91.9 | 45.49 [39.66, 51.78] | 44.56 [42.26, 45.10] | 45.74 [39.66, 52.12] | 90.9 |
| **LVEF (median [IQR])** | 59.47 [55.29, 63.52] | 59.14 [51.51, 63.31] | 59.48 [55.29, 63.52] | 91.9 | 58.64 [55.31, 62.35] | 51.02 [47.77, 53.27] | 58.91 [55.57, 62.38] | 90.9 |
| **LVPER (median [IQR])** | 373.80 [302.29, 452.71] | 284.40 [256.83, 364.56] | 373.81 [302.44, 453.27] | 91.9 | 351.29 [253.70, 463.30] | 350.15 [311.48, 380.62] | 351.29 [253.00, 486.30] | 90.9 |
| **LVPFR (median [IQR])** | 321.31 [259.23, 385.16] | 201.74 [189.85, 229.92] | 321.49 [259.95, 385.63] | 91.9 | 336.70 [260.80, 380.00] | 315.75 [273.85, 360.95] | 336.70 [259.70, 383.50] | 90.9 |
| **LVPAFR (median [IQR])** | 233.66 [167.35, 306.30] | 363.87 [208.97, 466.86] | 233.50 [167.46, 305.08] | 91.9 | 254.63 [181.10, 323.00] | 213.90 [135.76, 272.58] | 254.63 [182.20, 323.97] | 90.9 |
| **LVEDMi (median [IQR])** | 41.88 [36.52, 48.62] | 45.76 [34.02, 48.75] | 41.85 [36.55, 48.61] | 91.9 | 45.21 [37.68, 50.86] | 49.64 [46.84, 53.17] | 44.67 [37.47, 49.86] | 90.9 |
| **LVMVR (median [IQR])** | 0.56 [0.50, 0.62] | 0.70 [0.56, 0.70] | 0.56 [0.50, 0.62] | 91.9 | 0.57 [0.50, 0.65] | 0.58 [0.55, 0.61] | 0.57 [0.50, 0.65] | 90.9 |
| **LVEDV/RVEDV (median [IQR])** | 0.93 [0.86, 1.03] | 0.93 [0.93, 0.93] | 0.93 [0.86, 1.03] | 92.2 | 0.95 [0.86, 1.05] | 1.07 [0.99, 1.08] | 0.95 [0.86, 1.03] | 91 |
| **LVESV/RVESV (median [IQR])** | 0.91 [0.80, 1.04] | 1.15 [1.15, 1.15] | 0.91 [0.80, 1.04] | 92.2 | 0.96 [0.86, 1.10] | 1.27 [1.14, 1.30] | 0.96 [0.86, 1.09] | 91 |
| **peakEcc (median [IQR])** | -22.70 [-24.98, -20.43] | -21.60 [-21.61, -21.59] | -22.72 [-24.98, -20.42] | 93.7 | -22.59 [-24.30, -20.60] | -20.32 [-20.51, -20.27] | -22.78 [-24.41, -20.88] | 92.1 |
| **TPKEcc (median [IQR])** | 331.31 [309.83, 354.66] | 325.97 [321.27, 330.67] | 331.31 [309.75, 354.68] | 93.7 | 332.34 [309.34, 354.95] | 347.70 [320.45, 364.20] | 331.96 [309.94, 354.92] | 92.1 |
| **peakEll2Ch (median [IQR])** | -21.19 [-23.34, -18.93] | -23.89 [-23.89, -23.89] | -21.17 [-23.32, -18.93] | 93.8 | -20.32 [-22.79, -18.60] | -20.63 [-20.98, -20.18] | -20.28 [-22.92, -18.55] | 92.1 |
| **TPKEll2Ch (median [IQR])** | 349.86 [321.84, 379.10] | 388.50 [388.50, 388.50] | 349.86 [321.81, 379.08] | 93.9 | 364.20 [334.60, 384.50] | 403.20 [386.90, 420.80] | 362.52 [333.25, 382.95] | 92.1 |
| **peakEll4Ch (median [IQR])** | -23.30 [-25.97, -21.37] | -21.40 [-21.40, -21.40] | -23.30 [-25.99, -21.37] | 93.9 | -23.56 [-26.34, -21.77] | -20.87 [-22.20, -19.78] | -23.57 [-26.50, -22.22] | 93 |
| **TPKEll4Ch (median [IQR])** | 357.53 [327.07, 397.64] | 360.78 [360.78, 360.78] | 357.30 [326.96, 397.81] | 93.9 | 350.13 [323.33, 395.18] | 403.25 [378.53, 407.65] | 348.81 [323.33, 390.17] | 93 |
| **Wall thickness segment 1 (median [IQR])** | 7.65 [6.81, 8.50] | 7.04 [5.94, 8.14] | 7.65 [6.81, 8.49] | 93.2 | 8.15 [7.16, 9.17] | 9.35 [9.26, 9.44] | 7.93 [6.97, 9.11] | 94.1 |
| **Wall thickness segment 2 (median [IQR])** | 6.75 [5.75, 7.91] | 7.50 [6.70, 8.31] | 6.75 [5.74, 7.90] | 93.2 | 7.21 [5.77, 8.72] | 9.50 [8.42, 10.57] | 7.12 [5.70, 8.57] | 94.1 |
| **Wall thickness segment 3 (median [IQR])** | 6.05 [5.17, 6.96] | 7.60 [6.78, 8.42] | 6.05 [5.17, 6.95] | 93.2 | 6.42 [5.21, 7.97] | 7.99 [7.50, 8.47] | 6.37 [5.20, 7.96] | 94.1 |
| **Wall thickness segment 4 (median [IQR])** | 6.54 [5.82, 7.22] | 8.31 [7.66, 8.97] | 6.54 [5.81, 7.21] | 93.2 | 6.54 [6.09, 7.27] | 7.25 [7.12, 7.38] | 6.49 [6.07, 7.26] | 94.1 |
| **Wall thickness segment 5 (median [IQR])** | 6.20 [5.62, 6.96] | 6.33 [5.27, 7.39] | 6.20 [5.62, 6.96] | 93.2 | 6.25 [5.59, 6.76] | 6.36 [6.00, 6.72] | 6.25 [5.59, 6.75] | 94.1 |
| **Wall thickness segment 6 (median [IQR])** | 6.55 [5.97, 7.31] | 6.61 [5.42, 7.79] | 6.55 [5.98, 7.31] | 93.2 | 6.64 [6.16, 7.46] | 6.72 [6.45, 6.98] | 6.64 [6.15, 7.52] | 94.1 |
| **Wall thickness segment 7 (median [IQR])** | 5.73 [5.28, 6.31] | 5.48 [4.90, 6.07] | 5.73 [5.29, 6.31] | 93.2 | 5.75 [5.35, 6.33] | 6.03 [5.92, 6.13] | 5.74 [5.32, 6.37] | 94.1 |
| **Wall thickness segment 8 (median [IQR])** | 7.01 [6.28, 7.76] | 8.52 [7.77, 9.27] | 7.01 [6.28, 7.75] | 93.2 | 7.13 [6.27, 7.59] | 8.76 [7.66, 9.86] | 7.13 [6.18, 7.54] | 94.1 |
| **Wall thickness segment 9 (median [IQR])** | 7.38 [6.48, 8.25] | 9.47 [8.05, 10.89] | 7.38 [6.48, 8.25] | 93.2 | 7.60 [6.77, 8.32] | 8.99 [7.76, 10.21] | 7.60 [6.89, 8.29] | 94.1 |
| **Wall thickness segment 10 (median [IQR])** | 6.23 [5.59, 6.96] | 7.07 [6.08, 8.06] | 6.23 [5.60, 6.96] | 93.2 | 6.47 [5.87, 7.10] | 6.48 [6.17, 6.78] | 6.47 [5.88, 7.12] | 94.1 |
| **Wall thickness segment 11 (median [IQR])** | 5.62 [5.09, 6.32] | 5.57 [5.19, 5.95] | 5.62 [5.10, 6.32] | 93.2 | 5.68 [5.33, 6.54] | 5.75 [5.62, 5.88] | 5.68 [5.32, 6.69] | 94.1 |
| **Wall thickness segment 12 (median [IQR])** | 5.60 [5.22, 6.25] | 5.56 [5.53, 5.60] | 5.60 [5.22, 6.26] | 93.2 | 5.64 [5.25, 6.00] | 5.53 [5.32, 5.73] | 5.64 [5.32, 6.02] | 94.1 |
| **Wall thickness segment 13 (median [IQR])** | 5.48 [5.10, 5.91] | 8.01 [7.03, 9.00] | 5.48 [5.10, 5.91] | 93.2 | 5.59 [5.19, 5.93] | 5.21 [4.61, 5.82] | 5.59 [5.22, 5.91] | 94.1 |
| **Wall thickness segment 14 (median [IQR])** | 6.00 [5.36, 6.66] | 8.33 [6.87, 9.79] | 6.00 [5.36, 6.66] | 93.2 | 5.94 [5.49, 6.61] | 6.05 [5.36, 6.74] | 5.94 [5.53, 6.55] | 94.1 |
| **Wall thickness segment 15 (median [IQR])** | 5.09 [4.48, 5.69] | 7.21 [5.80, 8.63] | 5.09 [4.48, 5.69] | 93.2 | 5.16 [4.60, 5.73] | 5.30 [5.05, 5.54] | 5.16 [4.58, 5.70] | 94.1 |
| **Wall thickness segment 16 (median [IQR])** | 5.26 [4.80, 5.72] | 8.31 [6.87, 9.75] | 5.26 [4.80, 5.72] | 93.2 | 5.23 [5.01, 5.93] | 5.32 [4.77, 5.87] | 5.23 [5.02, 5.91] | 94.1 |
| **Global wall thickness (median [IQR])** | 6.30 [5.73, 6.84] | 7.33 [7.28, 7.38] | 6.30 [5.72, 6.84] | 93.2 | 6.51 [5.85, 7.17] | 7.06 [6.95, 7.17] | 6.44 [5.79, 7.17] | 94.1 |
| **Septal wall thickness (median [IQR])** | 6.68 [5.88, 7.35] | 8.28 [7.88, 8.69] | 6.67 [5.88, 7.34] | 93.2 | 6.84 [6.18, 8.03] | 8.26 [7.96, 8.55] | 6.74 [6.17, 8.00] | 94.1 |
| **Maximum wall thickness (median [IQR])** | 8.10 [7.24, 9.02] | 10.96 [10.29, 11.64] | 8.09 [7.24, 9.01] | 93.2 | 8.54 [7.65, 10.15] | 11.54 [11.49, 11.59] | 8.47 [7.59, 9.94] | 94.1 |

Abbreviations:
ARVC: arrythmogenic right ventricular cardiomyopathy; BMI: body mass index; CMR: cardiac magnetic resonance imaging; DCM: dilated cardiomyopathy; ECG: Electrocardiography; EDVi: indexed end-diastolic volume; EDMi: indexed end-diastolic mass; EF: ejection fraction; ESVi: indexed end-systolic volume; G+: carriers of likely pathogenic and pathogenic variants associated with one of the cardiomyopathies; HCM: hypertrophic cardiomyopathy; HDL: high-density lipoprotein; IQR: interquartile range; LDL: low-density lipoprotein; LV: left ventricular; MET: metabolic equivalent of task; MVR: mass to volume ratio; PAFR: peak atrial filling rate; peakEcc: peak circumferential strain; peakEll2Ch: longitudinal strain analyzed in 2-chamber view; peakEll4Ch: longitudinal strain analyzed in 4-chamber view; PER: peak ejection rate; PFR: peak filling rate; RV: right ventricular; SVi: indexed stroke volume; TPKEcc: global time to peak circumferential strain; TPKEll2Ch: global time to longitudinal strain analyzed in 2-chamber view; TPKEll4Ch: global time to longitudinal strain analyzed in 4-chamber view.

**Supplementary Table IV: Detailed information of all included SNPs**

| SNP | GRCh37 | Gene | rsID | Accession ClinVar | Canonical SPDI | N ARVC | N DCM | N HCM | MAF | Origin | Molecular Consequence | Amino acid change | Nucleotide change |
|---|---|---|---|---|---|---|---|---|---|---|---|---|---|
| 11:47332274:D:25 | 11:47353825 | *MYBPC3* | rs36212066 | VCV000177677 | NC_000011.10:47332274:GAGAGGGAGGG | NA | NA | 303 | 7.66E-04 | VKGL | NA | NA | 3628-41_3628-17del |
| 1:201359245:G:A | 1:201328373 | *TNNT2* | rs121964857 | VCV000012411 | NC_000001.11:201359244:G:A | NA | NA | 242 | 6.03E-04 | VKGL | Missense | Arg278Cys | 862C>T |
| 11:47342698:G:A | 11:47364249 | *MYBPC3* | rs375882485 | VCV000042540 | NC_000011.10:47342697:G:A | NA | NA | 88 | 2.19E-04 | VKGL | Missense | Arg502Trp | 1504C>T |
| 12:32802557:C:G | 12:32955491 | *PKP2* | rs193922674 | VCV000006756 | NC_000012.12:32802556:C:G | 46 | NA | NA | 1.15E-04 | ClinVar | Splice acceptor | NA | 2014-1G>C |
| 7:128856810:G:A | 7:128496864 | *FLNC* | rs778922568 | VCV000472173 | NC_000007.14:128856809:G:A | NA | **42** | NA | 1.05E-04 | VKGL | Missense | Gly2484Ser | 7450G>A |
| 12:32802499:D:5 | 12:32955434-32955438 | *PKP2* | rs397517021 | VCV000689321 | NC_000012.12:32802499:GGTGTG:G | 40 | NA | NA | 9.97E-05 | ClinVar | Frameshift | His689fs | 2066_2070del |
| 11:47350077:C:T | 11:47371628 | *MYBPC3* | rs397516050 | VCV000042752 | NC_000011.10:47350076:C:T | NA | NA | 34 | 8.47E-05 | VKGL | Missense | Gly148Arg | 442G>A |
| 14:23417573:A:G | 14:23886782 | *MYH7* | rs727503244 | VCV000164289 | NC_000014.9:23417572:A:G | NA | 34 | 34 | 8.47E-05 | VKGL | Missense | Leu1428Ser | 4283T>C |
| 2:219425699:C:A | 2:220290421 | *DES* | rs121913005 | VCV000626715 | NC_000002.12:219425698:C:A | NA | **31** | NA | 7.73E-05 | VKGL | Missense | Thr442Asn | 1325C>A |
| 11:47342574:T:A | 11:47364125 | *MYBPC3* | rs397515916 | VCV000042556 | NC_000011.10:47342573:T:A | NA | NA | 28 | 6.98E-05 | ClinVar | NA | NA | 1624+4A>T |
| 12:32878134:C:T | 12:33031068 | *PKP2* | rs1085307949 | VCV000427088 | NC_000012.12:32878133:C:T | 28 | NA | NA | 6.98E-05 | VKGL | Missense | Ser249Asn | 746G>A |
| 11:47343314:C:T | 11:47364865 | *MYBPC3* | NA | VCV000188544 | NC_000011.10:47343313:C:T | NA | NA | 26 | 6.48E-05 | ClinVar | NA | NA | 1224-52G>A |
| 14:23414007:C:T | 14:23883216 | *MYH7* | rs753392652 | VCV000378215 | NC_000014.9:23414006:C:T | NA | NA | 24 | 6.23E-05 | ClinVar | Synonymous | Ala1885= | 5655G>A |
| 14:23422292:G:A | 14:23891501 | *MYH7* | rs45611033 | VCV000177753 | NC_000014.9:23422291:G:A | NA | 24 | 24 | 5.98E-05 | ClinVar | Missense | Arg1045Cys | 3133C>T |
| 17:41757751:C:A | 17:39914003 | *JUP* | r200327969 | VCV000180376 | NC_000017.11:41757750:C:A | 24 | NA | NA | 5.98E-05 | VKGL | Missense | Val603Leu | 1807G>T |
| 11:47343117:G:A | 11:47364668 | *MYBPC3* | rs368770848 | VCV000042516 | NC_000011.10:47343116:G:A | NA | NA | 23 | 5.73E-05 | VKGL | Missense | Arg419Cys | 1255C>T |
| 11:19188286:A:G | 11:19209833 | *CSRP3* | rs104894205 | VCV000008778 | NC_000011.10:19188285:A:G | NA | NA | 21 | 5.23E-05 | VKGL | Missense | Leu44Pro | 131T>C |
| 11:47342578:C:G | 11:47364129 | *MYBPC3* | rs121909374 | VCV000008608 | NC_000011.10:47342577:C:G | NA | NA | 21 | 5.23E-05 | ClinVar | Missense | Glu542Gln | 1624G>C |
| 11:47346276:C:T | 11:47367827 | *MYBPC3* | rs397515881 | VCV000042499 | NC_000011.10:47346275:C:T | NA | NA | 20 | 4.98E-05 | VKGL | Missense | Gly341Ser | 1021G>A |
| 2:178528273:C:T | 2:179393000 | *TTN* | rs112188483 | VCV000196723 | NC_000002.12:178528272:C:T | NA | 20 | NA | 4.98E-05 | ClinVar | Splice donor | NA | 107377+1G>A |
| 14:23428957:C:T | 14:23898166 | *MYH7* | rs397516106 | VCV000177921 | NC_000014.9:23428956:C:T | NA | NA | 19 | 4.74E-05 | VKGL | Missense | Asp469Asn | 1405G>A |
| 3:38550326:C:T | 3:38591817 | *SCN5A* | rs762981322 | VCV000201549 | NC_000003.12:38550325:C:T | NA | 19 | NA | 4.74E-05 | VKGL | Missense | Val2015Met | 6043G>A |
| 12:32896580:D:4 | 12:33049514 | *PKP2* | rs397516997 | VCV000045028 | NC_000012.12:32896580:CTGTCTG:CTG | 18 | NA | NA | 4.49E-05 | VKGL | Frameshift | Thr50fs | 148_151del |
| 11:47351507:T:C | 11:47373058 | *MYBPC3* | rs376395543 | VCV000042644 | NC_000011.10:47351506:T:C | NA | NA | 17 | 4.24E-05 | ClinVar | Splice acceptor | NA | 26-2A>G |
| 2:178579702:G:A | 2:179444429 | *TTN* | rs574660186 | VCV000180573 | NC_000002.12:178579701:G:A | NA | 17 | NA | 4.24E-05 | ClinVar | Nonsense | Arg22499Ter | 67495C>T |
| 11:47337534:C:T | 11:47359085 | *MYBPC3* | rs2856655 | VCV000008617 | NC_000011.10:47337533:C:T | NA | NA | 16 | 3.99E-05 | ClinVar | Missense | Arg820Gln | 2459G>A |
| 18:31089461:C:T | 18:28669424 | *DSC2* | rs758527425 | VCV000372720 | NC_000018.10:31089460:C:T | 15 | NA | NA | 3.74E-05 | VKGL | Missense | Arg203His | 608G>A |
| 2:178613938:C:T | 2:179478665 | *TTN* | rs869312070 | VCV000223309 | NC_000002.12:178613937:C:T | NA | 15 | NA | 3.74E-05 | ClinVar | Splice acceptor | NA | 22151-1G>A |
| 14:23427746:T:C | 14:23896955 | *MYH7* | rs727504238 | VCV000177625 | NC_000014.9:23427745:T:C | NA | 14 | 14 | 3.49E-05 | ClinVar | Missense | His576Arg | 1727A>G |
| 1:156135268:C:T | 1:156105059 | *LMNA* | rs59885338 | VCV000014498 | NC_000001.11:156135267:C:T | NA | 13 | NA | 3.24E-05 | VKGL | Missense | Arg298Cys | 892C>T |
| 11:47337729:I:1 | 11:47359280 | *MYBPC3* | rs397515963 | VCV000042619 | NC_000011.10:47337729:C:CC | NA | NA | 13 | 3.24E-05 | ClinVar | Frameshift | Trp792fs | 2373dup |
| 14:23425316:C:T | 14:23894525 | *MYH7* | rs3218716 | VCV000042901 | NC_000014.9:23425315:C:T | NA | 13 | 13 | 3.24E-05 | ClinVar | Missense | Ala797Thr | 2389G>A |
| 18:31087815:T:C | 18:28667778 | *DSC2* | rs397514042 | VCV000016850 | NC_000018.10:31087814:T:C | 13 | NA | NA | 3.24E-05 | VKGL | Splice acceptor | NA | 631-2A>G |
| 19:55154821:G:A | 19:55666189 | *TNNI3* | rs730881068 | VCV000181575 | NC_000019.10:55154820:G:A | NA | 13 | 13 | 3.24E-05 | VKGL | Nonsense | Arg98Ter | 292C>T |
| 19:55154094:C:T | 19:55665462 | *TNNI3* | rs397516354 | VCV000043389 | NC_000019.10:55154093:C:T | NA | 12 | 12 | 2.99E-05 | ClinVar | Missense | Arg162Gln | 485G>A |
| 3:38562422:C:A | 3:38603913 | *SCN5A* | rs199473220 | VCV000067838 | NC_000003.12:38562421:C:A | NA | 12 | NA | 2.99E-05 | ClinVar | Missense | Gly1318Val | 3953G>T |
| 10:119669881:C:T | 10:121429393 | *BAG3* | rs387906874 | VCV000030396 | NC_000010.11:119669880:C:T | NA | 11 | NA | 2.74E-05 | VKGL | Missense | Arg71Trp | 211C>T |
| 2:178777234:D:5 | 2:179641961 | *TTN* | rs756433029 | VCV000202501 | NC_000002.12:178777234:TTTCATTTCA:TT | NA | 11 | NA | 2.74E-05 | VKGL | Frameshift | Met1575fs | 4724_4728del |
| 1:156134823:C:T | 1:156104614 | *LMNA* | rs370134870 | VCV000264626 | NC_000001.11:156134822:C:T | NA | 10 | NA | 2.49E-05 | VKGL | Missense | Arg220Cys | 658C>T |
| 11:47342718:C:T | 11:47364269 | *MYBPC3* | rs200411226 | VCV000164113 | NC_000011.10:47342717:C:T | NA | NA | 10 | 2.49E-05 | ClinVar | Missense | Arg495Gln | 1484G>A |
| 11:47348541:C:G | 11:47370092 | *MYBPC3* | rs397516068 | VCV000042784 | NC_000011.10:47348540:C:G | NA | NA | 10 | 2.49E-05 | ClinVar | Missense | Val219Leu | 655G>C |
| 2:178620285:G:T | 2:179485012 | *TTN* | rs368200299 | VCV000223308 | NC_000002.12:178620284:G:T | NA | 10 | NA | 2.49E-05 | ClinVar | Nonsense | Cys15412Ter | 46236C>A |
| 3:38613773:G:A | 3:38655264 | *SCN5A* | rs199473072 | VCV000068032 | NC_000003.12:38613772:G:A | NA | 10 | NA | 2.49E-05 | ClinVar | Missense | Arg225Trp | 673C>T |
| 6:7585760:C:G | 6:7585993 | *DSP* | NA | VCV000924608 | NC_000006.12:7585759:C:G | NA | 10 | NA | 2.49E-05 | VKGL | Missense | Ser2833Cys | 8498C>G |
| 11:47342854:G:A | 11:47364405 | *MYBPC3* | rs730880540 | VCV000180935 | NC_000011.10:47342853:G:A | NA | NA | 9 | 2.24E-05 | VKGL | Missense | Ser478Leu | 1433C>T |
| 11:47348424:C:T | 11:47369975 | *MYBPC3* | rs397516074 | VCV000042792 | NC_000011.10:47348423:C:T | NA | NA | 9 | 2.24E-05 | ClinVar | Missense | Glu258Lys | 772G>A |
| 12:110913140:G:A | 12:111350944 | *MYL2* | rs397516404 | VCV000043471 | NC_000012.12:110913139:G:A | NA | NA | 9 | 2.24E-05 | VKGL | Missense | Arg120Trp | 358C>T |
| 19:55154095:G:A | 19:55665463 | *TNNI3* | rs368861241 | VCV000161396 | NC_000019.10:55154094:G:A | NA | NA | 9 | 2.24E-05 | ClinVar | Missense | Arg162Trp | 484C>T |
| 2:178534401:A:G | 2:179399128 | *TTN* | rs375159973 | VCV000405075 | NC_000002.12:178534400:A:G | NA | 9 | NA | 2.24E-05 | VKGL | Missense | Trp34072Arg | 102214T>C |
| 2:178579850:T:G | 2:179444577 | *TTN* | rs753948675 | VCV000242425 | NC_000002.12:178579849:T:G | NA | 9 | NA | 2.24E-05 | ClinVar | Splice acceptor | NA | 67349-2A>C |
| 1:156135956:G:A | 1:156105747 | *LMNA* | rs59301204 | VCV000048098 | NC_000001.11:156135955:G:A | NA | 8 | NA | 1.99E-05 | VKGL | Missense | Arg331Gln | 992G>A |
| 1:201364327:G:A | 1:201333455 | *TNNT2* | rs483352832 | VCV000132943 | NC_000001.11:201364326:G:A | NA | 8 | 7 | 1.99E-05 | VKGL | Missense | Arg154Trp | 460C>T |
| 1:201365620:D:2 | 1:201334748 | *TNNT2* | NA | VCV000925600 | NC_000001.11:201365620:CTCTCTCTC:CTCT | NA | 8 | 8 | 1.99E-05 | VKGL | Frameshift | Arg94fs | 282_283del |
| 11:47342719:G:C | 11:47364270 | *MYBPC3* | rs397515905 | VCV000042537 | NC_000011.10:47342718:G:C | NA | NA | 8 | 1.99E-05 | ClinVar | Missense | Arg495Gly | 1483C>G |
| 12:110919133:C:T | 12:111356937 | *MYL2* | rs104894368 | VCV000014065 | NC_000012.12:110919132:C:T | NA | NA | 8 | 1.99E-05 | ClinVar | Missense | Glu22Lys | 64G>A |
| 12:32878545:T:A | 12:33031479 | *PKP2* | rs786204389 | VCV000188654 | NC_000012.12:32878544:T:A | 8 | NA | NA | 1.99E-05 | ClinVar | Splice acceptor | NA | 337-2A>T |
| 2:178531668:G:A | 2:179396395 | *TTN* | rs991187915 | VCV000667024 | NC_000002.12:178531667:G:A | NA | 8 | NA | 1.99E-05 | ClinVar | Nonsense | Gln34983Ter | 104947C>T |
| 3:38551513:G:A | 3:38593004 | *SCN5A* | rs199473282 | VCV000067932 | NC_000003.12:38551512:G:A | NA | 8 | NA | 1.99E-05 | ClinVar | Missense | Thr1619Met | 4856C>T |
| 3:52453993:G:A | 3:52488009 | *TNNC1* | rs267607125 | VCV000012443 | NC_000003.12:52453992:G:A | NA | 7 | 7 | 1.75E-05 | ClinVar | Missense | Ala8Val | 23C>T |

| SNP | GRCh37 | Gene | rsID | Accession ClinVar | Canonical SPDI | N ARVC | N DCM | N HCM | MAF | Origin | Molecular Consequence | Amino acid change | Nucleotide change |
|---|---|---|---|---|---|---|---|---|---|---|---|---|---|
| 11:47346379:C:T | 11:47367930 | *MYBPC3* | rs397516083 | VCV000042807 | NC_000011.10:47346378:C:T | NA | NA | 7 | 1.74E-05 | ClinVar | NA | NA | 927-9G>A |
| 2:178562716:G:A | 2:179427443 | *TTN* | NA | VCV000864799 | NC_000002.12:178562715:G:A | NA | 7 | NA | 1.74E-05 | ClinVar | Nonsense | Arg27806Ter | 83416C>T |
| 1:236727715:C:T | 1:236891015 | *ACTN2* | rs1253211384 | VCV000660714 | NC_000001.11:236727714:C:T | NA | **6** | NA | 1.50E-05 | VKGL | Nonsense | Arg192Ter | 574C>T |
| 11:47339792:T:C | 11:47361343 | *MYBPC3* | rs397515937 | VCV000042585 | NC_000011.10:47339791:T:C | NA | NA | 6 | 1.50E-05 | ClinVar | Splice acceptor | NA | 1928-2A>G |
| 12:32796108:C:T | 12:32949042 | *PKP2* | rs111517471 | VCV000006757 | NC_000012.12:32796107:C:T | 6 | NA | NA | 1.50E-05 | ClinVar | Splice donor | NA | 2357+1G>A |
| 12:32822616:I:1 | 12:32975550-32975551 | *PKP2* | rs397517010 | VCV000045047 | NC_000012.12:32822616:A:AA | 6 | NA | NA | 1.50E-05 | ClinVar | Frameshift | Val564fs | 1689dup |
| 12:32878981:A:T | 12:33031915 | *PKP2* | rs763639737 | VCV000202026 | NC_000012.12:32878980:A:T | 6 | NA | NA | 1.50E-05 | ClinVar | Nonsense | Leu92Ter | 275T>A |
| 14:23416057:G:A | 14:23885266 | *MYH7* | rs397516232 | VCV000043043 | NC_000014.9:23416056:G:A | NA | NA | 6 | 1.50E-05 | VKGL | Missense | Arg1634Cys | 4900C>T |
| 14:23424840:G:A | 14:23894049 | *MYH7* | rs138049878 | VCV000161326 | NC_000014.9:23424839:G:A | NA | 6 | 6 | 1.50E-05 | ClinVar | Missense | Arg870Cys | 2608C>T |
| 14:23428631:C:T | 14:23897840 | *MYH7* | rs121913651 | VCV000014119 | NC_000014.9:23428630:C:T | NA | NA | 6 | 1.50E-05 | VKGL | Missense | Glu483Lys | 1447G>A |
| 18:31070724:A:G | 18:28650690 | *DSC2* | rs1064793731 | VCV000419220 | NC_000018.10:31070723:A:G | 6 | NA | NA | 1.50E-05 | ClinVar | Splice donor | NA | 2250+2T>C |
| 2:178582209:C:G | 2:179446936 | *TTN* | rs1553627403 | VCV000466651 | NC_000002.12:178582208:C:G | NA | 6 | NA | 1.50E-05 | ClinVar | Splice acceptor | NA | 66161-1G>C |
| 2:178684990:C:T | 2:179549717 | *TTN* | rs371725574 | VCV000194146 | NC_000002.12:178684989:C:T | NA | 6 | NA | 1.50E-05 | VKGL | Splice acceptor | NA | 32471-1G>A |
| 20:44160293:G:A | 20:42788933 | *JPH2* | rs387906898 | VCV000030457 | NC_000020.11:44160292:G:A | NA | NA | 6 | 1.50E-05 | ClinVar | Missense | Ser165Phe | 494C>T |
| 11:19188281:T:G | 11:19209828 | *CSRP3* | rs137852765 | VCV000008781 | NC_000011.10:19188280:T:G | NA | NA | 5 | 1.25E-05 | VKGL | Missense | Ser46Arg | 136A>C |
| 11:47333189:C:G | 11:47354740 | *MYBPC3* | rs373746463 | VCV000042707 | NC_000011.10:47333188:C:G | NA | NA | 5 | 1.25E-05 | ClinVar | NA | NA | 3330+5G>T |
| 11:47341219:C:T | 11:47362770 | *MYBPC3* | rs368482358 | VCV000180951 | NC_000011.10:47341218:C:T | NA | NA | 5 | 1.25E-05 | VKGL | Missense | Val606Ile | 1816G>A |
| 11:47341991:C:T | 11:47363542 | *MYBPC3* | rs727503195 | VCV000164098 | NC_000011.10:47341990:C:T | NA | NA | 5 | 1.25E-05 | VKGL | Missense | Arg597Gln | 1790G>A |
| 11:47342611:C:G | 11:47364162 | *MYBPC3* | rs397515912 | VCV000042550 | NC_000011.10:47342610:C:G | NA | NA | 5 | 1.25E-05 | ClinVar | Missense | Gly531Arg | 1591G>C |
| 14:23417598:G:A | 14:23886807 | *MYH7* | rs145213771 | VCV000043003 | NC_000014.9:23417597:G:A | NA | 5 | 5 | 1.25E-05 | ClinVar | Missense | Arg1420Trp | 4258C>T |
| 14:23418304:G:A | 14:23887513 | *MYH7* | rs45451303 | VCV000178082 | NC_000014.9:23418303:G:A | NA | 5 | 5 | 1.25E-05 | VKGL | Missense | Arg1359Cys | 4075C>T |
| 14:23424112:T:C | 14:23893321 | *MYH7* | rs267606908 | VCV000014125 | NC_000014.9:23424111:T:C | NA | 5 | 5 | 1.25E-05 | ClinVar | Missense | Asp906Gly | 2717A>G |
| 14:23429037:C:T | 14:23898246 | *MYH7* | rs730880870 | VCV000181342 | NC_000014.9:23429036:C:T | NA | 5 | 5 | 1.25E-05 | VKGL | Missense | Arg442His | 1325G>A |
| 19:55156638:I:1 | 19:55668006 | *TNNI3* | rs772607683 | VCV000419596 | NC_000019.10:55156638:TTTTTT:TTTTTTT | NA | 5 | 5 | 1.25E-05 | VKGL | Frameshift | Ser39fs | 114dup |
| 2:178560865:G:A | 2:179425592 | *TTN* | NA | VCV000853671 | NC_000002.12:178560864:G:A | NA | 5 | NA | 1.25E-05 | ClinVar | Nonsense | Arg28423Ter | 85267C>T |
| 2:178574530:G:A | 2:179439257 | *TTN* | rs397517689 | VCV000047301 | NC_000002.12:178574529:G:A | NA | 5 | NA | 1.25E-05 | ClinVar | Nonsense | Arg23868Ter | 71602C>T |
| 2:178589849:G:A | 2:179454576 | *TTN* | rs72646846 | VCV000047175 | NC_000002.12:178589848:G:A | NA | 5 | NA | 1.25E-05 | ClinVar | Nonsense | Arg20626Ter | 61876C>T |
| 2:178767782:G:A | 2:179632509 | *TTN* | rs146572907 | VCV000282852 | NC_000002.12:178767781:G:A | NA | 5 | NA | 1.25E-05 | VKGL | Nonsense | Arg3150Ter | 9448C>T |
| 2:219423821:G:A | 2:220288543 | *DES* | rs112224037 | VCV000639517 | NC_000002.12:219423820:G:A | 5 | 5 | NA | 1.25E-05 | ClinVar | Splice donor | NA | 1288+1G>A |
| 6:7569211:G:A | 6:7569444 | *DSP* | NA | VCV000956247 | NC_000006.12:7569210:G:A | 5 | 5 | NA | 1.25E-05 | VKGL | Missense | Cys482Tyr | 1445G>A |
| 6:7579922:I:7 | 6:7580155-7580156 | *DSP* | rs1554108152 | VCV000199923 | NC_000006.12:7579922:GAAAATCGA:GAA/ | 5 | NA | NA | 1.25E-05 | ClinVar | Frameshift | Asp1248fs | 3735_3741dup |
| 1:156134454:C:T | 1:156104245 | *LMNA* | rs267607626 | VCV000066906 | NC_000001.11:156134453:C:T | NA | 4 | NA | 9.97E-06 | VKGL | Missense | Arg189Trp | 565C>T |
| 1:156136311:C:T | 1:156106102 | *LMNA* | rs1064793731 | VCV000242002 | NC_000001.11:156136310:C:T | NA | 4 | NA | 9.97E-06 | VKGL | Missense | Arg419Cys | 1255C>T |
| 1:201361317:A:C | 1:201330445 | *TNNT2* | rs730881110 | VCV000181645 | NC_000001.11:201361316:A:C | NA | 4 | 4 | 9.97E-06 | VKGL | Missense | Phe258Val | 772T>G |
| 10:110812459:C:T | 10:112572217 | *RBM20* | rs794729150 | VCV000202065 | NC_000010.11:110812458:C:T | NA | 4 | NA | 9.97E-06 | VKGL | Nonsense | Arg688Ter | 2062C>T |
| 11:47337544:G:A | 11:47359095 | *MYBPC3* | rs727503188 | VCV000164078 | NC_000011.10:47337543:G:A | NA | NA | 4 | 9.97E-06 | ClinVar | Missense | Arg817Trp | 2449C>T |
| 11:47342750:C:T | 11:47364301 | *MYBPC3* | rs375347534 | VCV000042533 | NC_000011.10:47342749:C:T | NA | NA | 4 | 9.97E-06 | VKGL | Na | NA | 1458-6G>A |
| 12:110911176:C:G | 12:111348980 | *MYL2* | rs199474813 | VCV000031768 | NC_000012.12:110911175:C:G | NA | NA | 4 | 9.97E-06 | ClinVar | Splice acceptor | NA | 403-1G>C |
| 12:32878217:G:T | 12:33031151 | *PKP2* | rs767987619 | VCV000201976 | NC_000012.12:32878216:G:T | 4 | NA | NA | 9.97E-06 | ClinVar | Nonsense | Tyr221Ter | 663C>A |
| 14:23415651:C:T | 14:23884860 | *MYH7* | rs193922390 | VCV000036642 | NC_000014.9:23415650:C:T | NA | 4 | 4 | 9.97E-06 | ClinVar | Missense | Arg1712Gln | 5135G>A |
| 14:23418348:C:T | 14:23887557 | *MYH7* | rs797045097 | VCV000208597 | NC_000014.9:23418347:C:T | NA | NA | 4 | 9.97E-06 | VKGL | Missense | Arg1344Gln | 4031G>A |
| 14:23424876:G:A | 14:23894085 | *MYH7* | rs2754158 | VCV000164324 | NC_000014.9:23424875:G:A | NA | 4 | 4 | 9.97E-06 | ClinVar | Missense | Arg858Cys | 2572C>T |
| 18:31086694:G:A | 18:28666657 | *DSC2* | rs397517404 | VCV000222557 | NC_000018.10:31086693:G:A | 4 | NA | NA | 9.97E-06 | VKGL | Missense | Thr275Met | 824C>T |
| 18:31498254:G:A | 18:29078217 | *DSG2* | rs1021457619 | VCV000657863 | NC_000018.10:31498253:G:A | 4 | NA | NA | 9.97E-06 | ClinVar | Missense | Met1Ile | 3G>A |
| 19:55151881:C:T | 19:55663249 | *TNNI3* | rs104894727 | VCV000012422 | NC_000019.10:55151880:C:T | NA | NA | 4 | 9.97E-06 | ClinVar | Missense | Asp196Asn | 586G>A |
| 2:178539559:G:A | 2:179404286 | *TTN* | rs869312085 | VCV000223329 | NC_000002.12:178539558:G:A | NA | 4 | NA | 9.97E-06 | ClinVar | Nonsense | Arg32836Ter | 98506C>T |
| 2:178584726:G:A | 2:179449453 | *TTN* | rs1432889079 | VCV000466649 | NC_000002.12:178584725:G:A | NA | 4 | NA | 9.97E-06 | ClinVar | Nonsense | Arg21639Ter | 64915C>T |
| 2:178590170:G:A | 2:179454897 | *TTN* | NA | VCV000202397 | NC_000002.12:178590169:G:A | NA | 4 | NA | 9.97E-06 | ClinVar | Nonsense | Arg20519Ter | 61555C>T |
| 2:219418784:G:T | 2:220283506 | *DES* | rs62636490 | VCV000804737 | NC_000002.12:219418783:G:T | 4 | 4 | NA | 9.97E-06 | ClinVar | Nonsense | Glu108Ter | 322G>T |
| 3:38597737:C:T | 3:38639228 | *SCN5A* | rs199473153 | VCV000067723 | NC_000003.12:38597736:C:T | NA | 4 | NA | 9.97E-06 | ClinVar | Missense | Gly752Arg | 2254G>A |
| 3:38603999:G:A | 3:38645490 | *SCN5A* | rs1417036453 | VCV000517279 | NC_000003.12:38603998:G:A | NA | 4 | NA | 9.97E-06 | ClinVar | Nonsense | Arg535Ter | 1603C>T |
| 6:118558947:G:A | 6:118880110 | *PLN* | rs754782171 | VCV000202037 | NC_000006.12:118558946:G:A | NA | 4 | NA | 9.97E-06 | VKGL | Missense | Arg9His | 26G>A |
| 6:7565521:G:A | 6:7565754 | *DSP* | rs727504443 | VCV000178282 | NC_000006.12:7565520:G:A | 4 | 4 | NA | 9.97E-06 | ClinVar | Splice donor | NA | 939+1G>A |
| 6:7583758:C:T | 6:7583991 | *DSP* | rs141026028 | VCV000199903 | NC_000006.12:7583757:C:T | 4 | 4 | NA | 9.97E-06 | ClinVar | Nonsense | Arg2166Ter | 6496C>T |
| 6:7585028:D:4 | 6:7585261 | *DSP* | NA | VCV000923199 | NC_000006.12:7585028:AGTAAGTAAG:AG | 4 | NA | NA | 9.97E-06 | VKGL | Frameshift | Ser2591fs | 7773_7776del |
| 12:32843181:C:T | 12:32996115 | *PKP2* | rs1332615728 | VCV000640418 | NC_000012.12:32843180:C:T | 3 | NA | NA | 7.50E-06 | ClinVar | Splice donor | NA | 1379-1976G>A |
| 1:201359636:C:T | 1:201328764 | *TNNT2* | rs121964861 | VCV000012417 | NC_000001.11:201359635:C:T | NA | 3 | NA | 7.48E-06 | ClinVar | Missense | Asp280Asn | 838G>A |
| 1:201361970:A:G | 1:201331098 | *TNNT2* | rs863225120 | VCV000217496 | NC_000001.11:201361969:A:G | NA | NA | 3 | 7.48E-06 | ClinVar | Missense | Ile221Thr | 662T>C |
| 11:47332075:G:A | 11:47353626 | *MYBPC3* | rs397516042 | VCV000042744 | NC_000011.10:47332074:G:A | NA | NA | 3 | 7.48E-06 | ClinVar | Nonsense | Arg1271Ter | 3811C>T |
| 11:47341204:C:T | 11:47362755 | *MYBPC3* | rs397515937 | VCV000180955 | NC_000011.10:47341203:C:T | NA | NA | 3 | 7.48E-06 | VKGL | Missense | Glu611Lys | 1831G>A |

| SNP | GRCh37 | Gene | rsID | Accession ClinVar | Canonical SPDI | N ARVC | N DCM | N HCM | MAF | Origin | Molecular Consequence | Amino acid change | Nucleotide change |
|---|---|---|---|---|---|---|---|---|---|---|---|---|---|
| 11:47342611:C:T | 11:47364162 | *MYBPC3* | rs397515912 | VCV000164109 | NC_000011.10:47342610:C:T | NA | NA | 3 | 7.48E-06 | VKGL | Missense | Gly531Arg | 1591G>A |
| 11:47348486:T:G | 11:47370037 | *MYBPC3* | rs397516070 | VCV000042787 | NC_000011.10:47348485:T:G | NA | NA | 3 | 7.48E-06 | ClinVar | Missense | Tyr237Ser | 710A>C |
| 12:32850907:G:A | 12:33003841 | *PKP2* | rs372827156 | VCV000045016 | NC_000012.12:32850906:G:A | 3 | NA | NA | 7.48E-06 | ClinVar | Nonsense | Arg413Ter | 1237C>T |
| 14:23424817:C:A | 14:23894026 | *MYH7* | rs1060505018 | VCV000417718 | NC_000014.9:23424816:C:A | NA | NA | 3 | 7.48E-06 | ClinVar | Missense | Met877Ile | 2631G>T |
| 14:23424839:C:T | 14:23894048 | *MYH7* | rs36211715 | VCV000014120 | NC_000014.9:23424838:C:T | NA | 3 | 3 | 7.48E-06 | ClinVar | Missense | Arg870His | 2609G>A |
| 14:23424854:T:A | 14:23894063 | *MYH7* | rs758891557 | VCV000454358 | NC_000014.9:23424853:T:A | NA | NA | 3 | 7.48E-06 | ClinVar | Missense | Lys865Met | 2594A>T |
| 14:23425814:G:A | 14:23895023 | *MYH7* | rs121913630 | VCV000014095 | NC_000014.9:23425813:G:A | NA | 3 | 3 | 7.48E-06 | ClinVar | Missense | Arg723Cys | 2167C>T |
| 14:23427723:C:T | 14:23896932 | *MYH7* | rs121913626 | VCV000042862 | NC_000014.9:23427722:C:T | NA | 3 | 3 | 7.48E-06 | ClinVar | Missense | Gly584Ser | 1750G>A |
| 14:23429038:G:A | 14:23898247 | *MYH7* | rs148808089 | VCV000177897 | NC_000014.9:23429037:G:A | NA | 3 | 3 | 7.48E-06 | ClinVar | Missense | Arg442Cys | 1324C>T |
| 14:23429850:C:T | 14:23899059 | *MYH7* | rs397516088 | VCV000042820 | NC_000014.9:23429849:C:T | NA | 3 | 3 | 7.48E-06 | ClinVar | Missense | Ala355Thr | 1063G>A |
| 18:31074908:C:G | 18:28654874 | *DSC2* | NA | VCV000860937 | NC_000018.10:31074907:C:G | 3 | NA | NA | 7.48E-06 | ClinVar | Splice acceptor | NA | 1664-1G>C |
| 18:31521213:I:1 | 18:29101176-29101177 | *DSG2* | rs781532110 | VCV000280230 | NC_000018.10:31521213:TT:TTT | 3 | NA | NA | 7.48E-06 | ClinVar | Frameshift | Gly166fs | 495dup |
| 18:31521233:G:T | 18:29101196 | *DSG2* | rs199926617 | VCV000577605 | NC_000018.10:31521232:G:T | 3 | NA | NA | 7.48E-06 | VKGL | Missense | Leu171Phe | 513G>T |
| 18:31524549:T:A | 18:29104512 | *DSG2* | rs869025388 | VCV000222562 | NC_000018.10:31524548:T:A | 3 | NA | NA | 7.48E-06 | ClinVar | Missense | Asp264Glu | 792T>A |
| 18:31524744:I:1 | 18:29104707-29104708 | *DSG2* | rs759944835 | VCV000639905 | NC_000018.10:31524744:A:AA | 3 | NA | NA | 7.48E-06 | ClinVar | Frameshift | Thr291fs | 871dup |
| 18:31541191:A:G | 18:29121154 | *DSG2* | rs397514038 | VCV000016817 | NC_000018.10:31541190:A:G | 3 | NA | NA | 7.48E-06 | ClinVar | Splice acceptor | NA | 1880-2A>G |
| 2:178546041:G:A | 2:179410768 | *TTN* | rs753334568 | VCV000132137 | NC_000002.12:178546040:G:A | NA | 3 | NA | 7.48E-06 | ClinVar | Missense | Pro31732Leu | 95195C>T |
| 2:178548460:G:A | 2:179413187 | *TTN* | rs72648250 | VCV000223326 | NC_000002.12:178548459:G:A | NA | 3 | NA | 7.48E-06 | ClinVar | Nonsense | Arg31056Ter | 93166C>T |
| 2:178563493:C:A | 2:179428220 | *TTN* | rs779874042 | VCV000202416 | NC_000002.12:178563492:C:A | NA | 3 | NA | 7.48E-06 | ClinVar | Nonsense | Glu27547Ter | 82639G>T |
| 2:178569267:C:T | 2:179433994 | *TTN* | rs756552975 | VCV000379555 | NC_000002.12:178569266:C:T | NA | 3 | NA | 7.48E-06 | ClinVar | Nonsense | Trp25622Ter | 76865G>A |
| 2:178569478:G:A | 2:179434205 | *TTN* | rs545954490 | VCV000404828 | NC_000002.12:178569477:G:A | NA | 3 | NA | 7.48E-06 | ClinVar | Nonsense | Arg25552Ter | 76654C>T |
| 2:178570804:G:A | 2:179435531 | *TTN* | rs794729382 | VCV000202521 | NC_000002.12:178570803:G:A | NA | 3 | NA | 7.48E-06 | ClinVar | Nonsense | Arg25110Ter | 75328C>T |
| 2:178575970:G:A | 2:179440697 | *TTN* | rs781540455 | VCV000202402 | NC_000002.12:178575969:G:A | NA | 3 | NA | 7.48E-06 | ClinVar | Nonsense | Arg23388Ter | 70162C>T |
| 2:178585291:G:A | 2:179450018 | *TTN* | rs768345594 | VCV000223315 | NC_000002.12:178585290:G:A | NA | 3 | NA | 7.48E-06 | ClinVar | Nonsense | Arg21485Ter | 64453C>T |
| 2:178588700:G:A | 2:179453427 | *TTN* | rs368452607 | VCV000202518 | NC_000002.12:178588699:G:A | NA | 3 | NA | 7.48E-06 | ClinVar | Nonsense | Arg21009Ter | 63025C>T |
| 2:178593338:G:A | 2:179458065 | *TTN* | rs1553649171 | VCV000466646 | NC_000002.12:178593337:G:A | NA | 3 | NA | 7.48E-06 | ClinVar | Nonsense | Arg19624Ter | 58870C>T |
| 2:178609756:G:A | 2:179474483 | *TTN* | NA | VCV001066907 | NC_000002.12:178609755:G:A | NA | 3 | NA | 7.48E-06 | VKGL | Nonsense | Arg17223Ter | 51667C>T |
| 2:178733497:G:A | 2:179598224 | *TTN* | rs372277017 | VCV000130662 | NC_000002.12:178733496:G:A | NA | 3 | NA | 7.48E-06 | VKGL | Nonsense | Arg5266Ter | 15796C>T |
| 2:219421560:G:A | 2:220286282 | *DES* | rs1262288015 | VCV000626714 | NC_000002.12:219421559:G:A | NA | **3** | NA | 7.48E-06 | VKGL | Missense | Arg415Gln | 1244G>A |
| 2:219423817:C:T | 2:220288539 | *DES* | rs150974575 | VCV000177872 | NC_000002.12:219423816:C:T | 3 | 3 | NA | 7.48E-06 | ClinVar | Nonsense | Arg429Ter | 1285C>T |
| 6:118558994:C:T | 6:118880157 | *PLN* | rs761056344 | VCV000202040 | NC_000006.12:118558993:C:T | NA | 3 | NA | 7.48E-06 | VKGL | Missense | Arg25Cys | 73C>T |
| 6:7580388:C:T | 6:7580621 | *DSP* | rs770873593 | VCV000199884 | NC_000006.12:7580387:C:T | 3 | 3 | NA | 7.48E-06 | ClinVar | Nonsense | Arg1400Ter | 4198C>T |
| 6:7580547:C:T | 6:7580780 | *DSP* | rs1561698750 | VCV000576091 | NC_000006.12:7580546:C:T | 3 | 3 | NA | 7.48E-06 | ClinVar | Nonsense | Gln1453Ter | 4357C>T |
| 7:128845989:G:C | 7:128486043 | *FLNC* | rs781135153 | VCV000420146 | NC_000007.14:128845988:G:C | NA | **3** | NA | 7.48E-06 | ClinVar | Splice acceptor | NA | 3791-1G>C |
| 7:128846444:C:T | 7:128486498 | *FLNC* | NA | VCV000842060 | NC_000007.14:128846443:C:T | NA | **3** | NA | 7.48E-06 | ClinVar | Nonsense | Arg1370Ter | 4108C>T |
| 12:32802499:G:A | 12:32955433 | *PKP2* | rs121434421 | VCV000006755 | NC_000012.12:32802498:G:A | 2 | NA | NA | 4.99E-06 | ClinVar | Nonsense | Arg691Ter | 2071C>T |
| 1:156135913:G:A | 1:156105704 | *LMNA* | rs56816490 | VCV000048093 | NC_000001.11:156135912:G:A | NA | 2 | NA | 4.98E-06 | ClinVar | Missense | Glu317Lys | 949G>A |
| 1:201359217:C:T | 1:201328345 | *TNNT2* | rs727504247 | VCV000177636 | NC_000001.11:201359216:C:T | NA | 2 | 2 | 4.98E-06 | ClinVar | Nonsense | Trp297Ter | 890G>A |
| 1:201363352:C:A | 1:201332480 | *TNNT2* | rs730881097 | VCV000181612 | NC_000001.11:201363351:C:A | NA | 2 | 2 | 4.98E-06 | VKGL | Missense | Ala182Ser | 544G>T |
| 1:77926827:G:T | 1:78392512 | *NEXN* | rs771262904 | VCV000599095 | NC_000001.11:77926826:G:T | NA | 2 | NA | 4.98E-06 | ClinVar | Nonsense | Glu267Ter | 799G>T |
| 1:77942736:C:G | 1:78408421 | *NEXN* | rs794729086 | VCV000201935 | NC_000001.11:77942735:C:G | NA | 2 | NA | 4.98E-06 | ClinVar | Missense | Phe645Leu | 1935C>G |
| 10:119670037:C:T | 10:121429549 | *BAG3* | rs387906875 | VCV000030397 | NC_000010.11:119670036:C:T | NA | 2 | NA | 4.98E-06 | ClinVar | Nonsense | Arg123Ter | 367C>T |
| 10:119676479:C:T | 10:121435991 | *BAG3* | rs869248137 | VCV000228322 | NC_000010.11:119676478:C:T | NA | 2 | NA | 4.98E-06 | ClinVar | Nonsense | Arg309Ter | 925C>T |
| 11:47332105:C:T | 11:47353656 | *MYBPC3* | rs730880141 | VCV000180414 | NC_000011.10:47332104:C:T | NA | NA | 2 | 4.98E-06 | VKGL | Missense | Glu1261Lys | 3781G>A |
| 11:47332894:D:3 | 11:47354445 | *MYBPC3* | rs730880674 | VCV000181102 | NC_000011.10:47332894:AGTAGTAG:AGTA | NA | NA | 2 | 4.98E-06 | VKGL | NA | Tyr1136del | 3404ACT[1] |
| 11:47333552:C:T | 11:47355103 | *MYBPC3* | rs587782958 | VCV000155808 | NC_000011.10:47333551:C:T | NA | NA | 2 | 4.98E-06 | ClinVar | NA | NA | 3190+5G>A |
| 11:47335041:C:T | 11:47356592 | *MYBPC3* | rs397515991 | VCV000042666 | NC_000011.10:47335040:C:T | NA | NA | 2 | 4.98E-06 | VKGL | Splice donor | NA | 2905+1G>A |
| 11:47335120:G:A | 11:47356671 | *MYBPC3* | rs387907267 | VCV000037039 | NC_000011.10:47335119:G:A | NA | NA | 2 | 4.98E-06 | ClinVar | Nonsense | Arg943Ter | 2827C>T |
| 11:47335165:D:2 | 11:47356716 | *MYBPC3* | rs727504265 | VCV000177660 | NC_000011.10:47335165:TGTGTG:TGTG | NA | NA | 2 | 4.98E-06 | ClinVar | Frameshift | Thr927fs | 2780_2781del |
| 11:47336003:D:1 | 11:47357554 | *MYBPC3* | rs397515979 | VCV000181083 | NC_000011.10:47336003:GGGGGG:GGGG | NA | NA | 2 | 4.98E-06 | ClinVar | Frameshift | Ser871fs | 2610del |
| 11:47342096:G:A | 11:47363647 | *MYBPC3* | rs730880694 | VCV000656085 | NC_000011.10:47342095:G:A | NA | NA | 2 | 4.98E-06 | VKGL | Missense | Ala562Val | 1685C>T |
| 11:47342697:C:T | 11:47364248 | *MYBPC3* | rs397515907 | VCV000042541 | NC_000011.10:47342696:C:T | NA | NA | 2 | 4.98E-06 | ClinVar | Missense | Arg502Gln | 1505G>A |
| 11:47347856:C:T | 11:47369407 | *MYBPC3* | rs397516073 | VCV000042791 | NC_000011.10:47347855:C:T | NA | NA | 2 | 4.98E-06 | ClinVar | Splice donor | NA | 821+1G>A |
| 11:47352622:C:T | 11:47374173 | *MYBPC3* | rs113709679 | VCV000810740 | NC_000011.10:47352621:C:T | NA | NA | 2 | 4.98E-06 | ClinVar | Splice donor | NA | 25+1G>A |
| 12:32802402:C:T | 12:32955336 | *PKP2* | rs794729116 | VCV000202005 | NC_000012.12:32802401:C:T | 2 | NA | NA | 4.98E-06 | ClinVar | Splice donor | NA | 2167+1G>A |
| 12:32821502:C:A | 12:32974436 | *PKP2* | rs397517015 | VCV000045054 | NC_000012.12:32821501:C:A | 2 | NA | NA | 4.98E-06 | ClinVar | Nonsense | Glu623Ter | 1867G>T |
| 12:32822502:I:19 | 12:32822502-32822503 | *PKP2* | rs1555142971 | VCV000523703 | NC_000012.12:32822502:AATACTTTTGTTG | 2 | NA | NA | 4.98E-06 | ClinVar | Nonsense | Gly602Ter | 1785_1803dup |
| 12:32868965:G:A | 12:33021899 | *PKP2* | rs397516986 | VCV000045010 | NC_000012.12:32868964:G:A | 2 | NA | NA | 4.98E-06 | ClinVar | Nonsense | Gln378Ter | 1132C>T |
| 14:23415385:G:A | 14:23884594 | *MYH7* | rs727505294 | VCV000180024 | NC_000014.9:23415384:G:A | NA | 2 | 2 | 4.98E-06 | VKGL | Missense | Thr1760Met | 5279C>T |
| 14:23417670:G:A | 14:23886879 | *MYH7* | rs730880793 | VCV000181246 | NC_000014.9:23417669:G:A | NA | NA | 2 | 4.98E-06 | VKGL | Missense | Arg1396Trp | 4186C>T |

| SNP | GRCh37 | Gene | rsID | Accession ClinVar | Canonical SPDI | N ARVC | N DCM | N HCM | MAF | Origin | Molecular Consequence | Amino acid change | Nucleotide change |
|---|---|---|---|---|---|---|---|---|---|---|---|---|---|
| 14:23418244:C:T | 14:23887453 | *MYH7* | rs397516202 | VCV000042993 | NC_000014.9:23418243:C:T | NA | 2 | 2 | 4.98E-06 | ClinVar | Missense | Ala1379Thr | 4135G>A |
| 14:23418313:C:T | 14:23887522 | *MYH7* | rs727503246 | VCV000164294 | NC_000014.9:23418312:C:T | NA | NA | 2 | 4.98E-06 | ClinVar | Missense | Glu1356Lys | 4066G>A |
| 14:23424107:G:C | 14:23893316 | *MYH7* | rs121913631 | VCV000014097 | NC_000014.9:23424106:G:C | NA | 2 | 2 | 4.98E-06 | ClinVar | Missense | Leu908Val | 2722C>G |
| 14:23424909:T:C | 14:23894118 | *MYH7* | rs727504310 | VCV000177757 | NC_000014.9:23424908:T:C | NA | 2 | 2 | 4.98E-06 | ClinVar | Missense | Lys847Glu | 2539A>G |
| 14:23426045:C:T | 14:23895254 | *MYH7* | rs886039030 | VCV000264068 | NC_000014.9:23426044:C:T | NA | 2 | 2 | 4.98E-06 | ClinVar | Missense | Arg694His | 2081G>A |
| 14:23429255:C:T | 14:23898464 | *MYH7* | rs730880868 | VCV000181339 | NC_000014.9:23429254:C:T | NA | 2 | 2 | 4.98E-06 | VKGL | Missense | Val411Ile | 1231G>A |
| 14:23431602:C:T | 14:23900811 | *MYH7* | rs397516264 | VCV000043100 | NC_000014.9:23431601:C:T | NA | 2 | 2 | 4.98E-06 | ClinVar | Missense | Asp239Asn | 715G>A |
| 14:23431790:G:A | 14:23900999 | *MYH7* | rs397516259 | VCV000181315 | NC_000014.9:23431789:G:A | NA | NA | 2 | 4.98E-06 | VKGL | Missense | Arg204Cys | 610C>T |
| 15:63060899:G:A | 15:63353098 | *TPM1* | rs104894503 | VCV000012456 | NC_000015.10:63060898:G:A | NA | 2 | 2 | 4.98E-06 | ClinVar | Missense | Asp175Asn | 523G>A |
| 15:63061723:G:A | 15:63353922 | *TPM1* | rs199476315 | VCV000031882 | NC_000015.10:63061722:G:A | NA | NA | 2 | 4.98E-06 | ClinVar | Missense | Glu192Lys | 574G>A |
| 15:63061751:C:T | 15:63353950 | *TPM1* | rs730881141 | VCV000181668 | NC_000015.10:63061750:C:T | NA | NA | 2 | 4.98E-06 | VKGL | Missense | Thr201Met | 602C>T |
| 18:31519867:G:A | 18:29099830 | *DSG2* | rs121913006 | VCV000016810 | NC_000018.10:31519866:G:A | 2 | NA | NA | 4.98E-06 | ClinVar | Missense | Arg49His | 146G>A |
| 18:31524752:I:1 | 18:29104715-29104716 | *DSG2* | rs1187924885 | VCV000691669 | NC_000018.10:31524752:AAAA:AAAAA | 2 | NA | NA | 4.98E-06 | ClinVar | Frameshift | Val295fs | 882dup |
| 18:31545783:T:G | 18:29125746 | *DSG2* | NA | VCV000943833 | NC_000018.10:31545782:T:G | 2 | NA | NA | 4.98E-06 | ClinVar | Nonsense | Tyr799Ter | 2397T>G |
| 19:55154082:G:A | 19:55665450 | *TNNI3* | rs727504242 | VCV000177630 | NC_000019.10:55154081:G:A | NA | NA | 2 | 4.98E-06 | ClinVar | Missense | Ser166Phe | 497C>T |
| 19:55154145:C:T | 19:55665513 | *TNNI3* | rs397516349 | VCV000043384 | NC_000019.10:55154144:C:T | NA | 2 | 2 | 4.98E-06 | ClinVar | Missense | Arg145Gln | 434G>A |
| 19:55154146:G:A | 19:55665514 | *TNNI3* | rs104894724 | VCV000012426 | NC_000019.10:55154145:G:A | NA | 2 | 2 | 4.98E-06 | ClinVar | Missense | Arg145Trp | 433C>T |
| 2:178531788:G:A | 2:179396515 | *TTN* | NA | VCV000954597 | NC_000002.12:178531787:G:A | NA | 2 | NA | 4.98E-06 | ClinVar | Nonsense | Arg34943Ter | 104827C>T |
| 2:178531962:G:A | 2:179396689 | *TTN* | rs1057518003 | VCV000372824 | NC_000002.12:178531961:G:A | NA | 2 | NA | 4.98E-06 | ClinVar | Nonsense | Arg34885Ter | 104653C>T |
| 2:178532670:G:A | 2:179397397 | *TTN* | rs995029896 | VCV000570433 | NC_000002.12:178532669:G:A | NA | 2 | NA | 4.98E-06 | ClinVar | Nonsense | Arg34649Ter | 103945C>T |
| 2:178532910:T:A | 2:179397637 | *TTN* | rs1553490574 | VCV000499641 | NC_000002.12:178532909:T:A | NA | 2 | NA | 4.98E-06 | ClinVar | Nonsense | Lys34569Ter | 103705A>T |
| 2:178534092:G:A | 2:179398819 | *TTN* | rs752697861 | VCV000464497 | NC_000002.12:178534091:G:A | NA | 2 | NA | 4.98E-06 | ClinVar | Nonsense | Arg34175Ter | 102523C>T |
| 2:178535508:G:A | 2:179400235 | *TTN* | rs766265889 | VCV000625156 | NC_000002.12:178535507:G:A | NA | 2 | NA | 4.98E-06 | ClinVar | Nonsense | Arg33703Ter | 101107C>T |
| 2:178535729:C:T | 2:179400456 | *TTN* | rs1260821931 | VCV000488972 | NC_000002.12:178535728:C:T | NA | 2 | NA | 4.98E-06 | ClinVar | Nonsense | Trp33629Ter | 100886G>A |
| 2:178542263:C:G | 2:179406990 | *TTN* | rs727505319 | VCV000180058 | NC_000002.12:178542262:C:G | NA | 2 | NA | 4.98E-06 | ClinVar | Splice donor | NA | 97492+1G>C |
| 2:178547518:D:5 | 2:179412245 | *TTN* | rs769488730 | VCV000202493 | NC_000002.12:178547518:TTTAATTT:TTT | NA | 2 | NA | 4.98E-06 | VKGL | Frameshift | p.Ile31368fs | 94103_94107del |
| 2:178568057:G:T | 2:179432784 | *TTN* | rs1553597198 | VCV000535021 | NC_000002.12:178568056:G:T | NA | 2 | NA | 4.98E-06 | ClinVar | Missense | Tyr26025Ter | 78075C>A |
| 2:178575154:G:A | 2:179439881 | *TTN* | rs1553612386 | VCV000466655 | NC_000002.12:178575153:G:A | NA | 2 | NA | 4.98E-06 | ClinVar | Nonsense | Arg23660Ter | 70978C>T |
| 2:178576691:G:A | 2:179441418 | *TTN* | rs878854328 | VCV000238830 | NC_000002.12:178576690:G:A | NA | 2 | NA | 4.98E-06 | ClinVar | Nonsense | Arg23185Ter | 69553C>T |
| 2:178592916:D:2 | 2:179457644-179457645 | *TTN* | rs752948913 | VCV000419310 | NC_000002.12:178592916:AG: | NA | 2 | NA | 4.98E-06 | ClinVar | Frameshift | Pro19734fs | 59201_59202del |
| 2:178609289:G:A | 2:179474016 | *TTN* | rs926741242 | VCV000405082 | NC_000002.12:178609288:G:A | NA | 2 | NA | 4.98E-06 | ClinVar | Nonsense | Arg17341Ter | 52021C>T |
| 2:178612430:G:A | 2:179477157 | *TTN* | NA | VCV000862652 | NC_000002.12:178612429:G:A | NA | 2 | NA | 4.98E-06 | ClinVar | Nonsense | Gln16699Ter | 50095C>T |
| 2:178617857:G:A | 2:179482584 | *TTN* | rs751746401 | VCV000264517 | NC_000002.12:178617856:G:A | NA | 2 | NA | 4.98E-06 | ClinVar | Nonsense | Arg15832Ter | 47494C>T |
| 2:178740646:G:T | 2:179605373 | *TTN* | rs370912401 | VCV000047827 | NC_000002.12:178740645:G:T | NA | 2 | NA | 4.98E-06 | VKGL | Nonsense | Ser4196Ter | 12587C>A |
| 2:178766474:G:A | 2:179631201 | *TTN* | rs757836789 | VCV000288998 | NC_000002.12:178766473:G:A | NA | 2 | NA | 4.98E-06 | VKGL | Nonsense | Arg3204Ter | 9610C>T |
| 2:219418497:C:T | 2:220283219 | *DES* | rs267607495 | VCV000066412 | NC_000002.12:219418496:C:T | 2 | 2 | NA | 4.98E-06 | ClinVar | Missense | Ser12Phe | 35C>T |
| 3:38566426:C:T | 3:38607917 | *SCN5A* | rs137854618 | VCV000009401 | NC_000003.12:38566425:C:T | NA | 2 | NA | 4.98E-06 | ClinVar | Missense | Asp1274Asn | 3820G>A |
| 6:7568443:C:T | 6:7568676 | *DSP* | rs397516915 | VCV000044856 | NC_000006.12:7568442:C:T | 2 | 2 | NA | 4.98E-06 | ClinVar | Nonsense | Arg425Ter | 1273C>T |
| 6:7579527:C:T | 6:7579760 | *DSP* | rs746877365 | VCV000405247 | NC_000006.12:7579526:C:T | 2 | 2 | NA | 4.98E-06 | ClinVar | Nonsense | Arg1113Ter | 3337C>T |
| 6:7579995:C:T | 6:7580228 | *DSP* | rs767643821 | VCV000199881 | NC_000006.12:7579994:C:T | 2 | 2 | NA | 4.98E-06 | ClinVar | Nonsense | Arg1269Ter | 3805C>T |
| 6:7582690:C:T | 6:7582923 | *DSP* | rs397516946 | VCV000044928 | NC_000006.12:7582689:C:T | 2 | NA | NA | 4.98E-06 | ClinVar | Nonsense | Gln1810Ter | 5428C>T |
| 7:128844045:C:T | 7:128484099 | *FLNC* | rs886037830 | VCV000267288 | NC_000007.14:128844044:C:T | NA | **2** | NA | 4.98E-06 | ClinVar | Nonsense | Arg991Ter | 2971C>T |
| 7:128846136:C:T | 7:128486190 | *FLNC* | rs766330686 | VCV000579589 | NC_000007.14:128846135:C:T | NA | **2** | NA | 4.98E-06 | ClinVar | Nonsense | Arg1313Ter | 3937C>T |
| 3:46860702:C:T | 3:46902192 | *MYL3* | rs199474703 | VCV000031777 | NC_000003.12:46860701:C:T | NA | NA | 1 | 2.49E-06 | ClinVar | Missense | Arg94His | 281G>A |
| 6:7584359:G:A | 6:7584592 | *DSP* | rs387906618 | VCV000029672 | NC_000006.12:7584358:G:A | NA | 1 | NA | 2.49E-06 | ClinVar | Missense | Arg2366His | 7097G>A |
| 2:178774206:C:T | 2:179638933 | *TTN* | NA | VCV000873434 | NC_000002.12:178774205:C:T | NA | 1 | NA | 2.49E-06 | VKGL | Splice donor | NA | 7057+1G>A |
| 1:156134458:G:A | 1:156104249 | *LMNA* | rs267607571 | VCV000066910 | NC_000001.11:156134457:G:A | NA | 1 | NA | 2.49E-06 | ClinVar | Missense | Arg190Gln | 569G>A |
| 1:201361989:G:A | 1:201331117 | *TNNT2* | rs45586240 | VCV000180554 | NC_000001.11:201361988:G:A | NA | 1 | 1 | 2.49E-06 | VKGL | Missense | Arg215Trp | 643C>T |
| 1:201362016:G:A | 1:201331144 | *TNNT2* | NA | VCV000181625 | NC_000001.11:201362015:G:A | NA | 1 | 1 | 2.49E-06 | ClinVar | Missense | Arg206Trp | 616C>T |
| 1:201363349:G:A | 1:201332477 | *TNNT2* | rs727503512 | VCV000228409 | NC_000001.11:201363348:G:A | NA | 1 | 1 | 2.49E-06 | ClinVar | Missense | Arg183Trp | 547C>T |
| 1:201365291:C:A | 1:201334419 | *TNNT2* | rs397516457 | VCV000043629 | NC_000001.11:201365290:C:A | NA | 1 | 1 | 2.49E-06 | ClinVar | Missense | Arg104Leu | 311G>T |
| 1:201365638:A:T | 1:201334766 | *TNNT2* | rs121964855 | VCV000012408 | NC_000001.11:201365637:A:T | NA | 1 | 1 | 2.49E-06 | ClinVar | Missense | Ile89Asn | 266T>A |
| 10:110812573:C:T | 10:112572331 | *RBM20* | rs1393804220 | VCV000538028 | NC_000010.11:110812572:C:T | NA | 1 | NA | 2.49E-06 | VKGL | Nonsense | Arg726Ter | 2176C>T |
| 11:19188245:A:C | 11:19209792 | *CSRP3* | rs104894204 | VCV000008777 | NC_000011.10:19188244:A:C | NA | NA | 1 | 2.49E-06 | ClinVar | Missense | Cys58Gly | 172T>G |
| 11:47332189:G:A | 11:47353740 | *MYBPC3* | rs397516037 | VCV000042735 | NC_000011.10:47332188:G:A | NA | NA | 1 | 2.49E-06 | ClinVar | Nonsense | Gln1233Ter | 3697C>T |
| 11:47333923:T:C | 11:47355474 | *MYBPC3* | rs727503177 | VCV000164052 | NC_000011.10:47333922:T:C | NA | NA | 1 | 2.49E-06 | VKGL | Missense | Gln998Arg | 2993A>G |
| 11:47335081:D:2 | 11:47356632 | *MYBPC3* | rs397515990 | VCV000042663 | NC_000011.10:47335081:AG: | NA | NA | 1 | 2.49E-06 | ClinVar | Frameshift | Pro955fs | 2864_2865del |
| 11:47337535:G:A | 11:47359086 | *MYBPC3* | rs775404728 | VCV000195850 | NC_000011.10:47337534:G:A | NA | NA | 1 | 2.49E-06 | VKGL | Missense | Arg820Trp | 2458C>T |
| 11:47341230:G:A | 11:47362781 | *MYBPC3* | rs730880551 | VCV000180950 | NC_000011.10:47341229:G:A | NA | NA | 1 | 2.49E-06 | ClinVar | Missense | Thr602Ile | 1805C>T |
| 11:47342882:D:1 | 11:47364433 | *MYBPC3* | rs886037900 | VCV000254153 | NC_000011.10:47342882:CCCC:CCC | NA | NA | 1 | 2.49E-06 | VKGL | Frameshift | Gln469fs | 1404del |

| SNP | GRCh37 | Gene | rsID | Accession ClinVar | Canonical SPDI | N ARVC | N DCM | N HCM | MAF | Origin | Molecular Consequence | Amino acid change | Nucleotide change |
|---|---|---|---|---|---|---|---|---|---|---|---|---|---|
| 11:47343019:A:G | 11:47364570 | *MYBPC3* | rs397515897 | VCV000042525 | NC_000011.10:47343018:A:G | NA | NA | 1 | 2.49E-06 | ClinVar | Splice donor | NA | 1351+2T>C |
| 11:47343147:T:C | 11:47364698 | *MYBPC3* | rs730880531 | VCV000180925 | NC_000011.10:47343146:T:C | NA | NA | 1 | 2.49E-06 | ClinVar | Splice acceptor | NA | 1227-2A>G |
| 11:47343342:C:T | 11:47364893 | *MYBPC3* | rs1025692267 | VCV000693982 | NC_000011.10:47343341:C:T | NA | NA | 1 | 2.49E-06 | ClinVar | NA | NA | 1224-80G>A |
| 11:47343505:G:A | 11:47365056 | *MYBPC3* | rs727504329 | VCV000177796 | NC_000011.10:47343504:G:A | NA | NA | 1 | 2.49E-06 | ClinVar | Nonsense | Gln404Ter | 1210C>T |
| 11:47346217:C:G | 11:47367768 | *MYBPC3* | rs730880632 | VCV000181057 | NC_000011.10:47346216:C:G | NA | NA | 1 | 2.49E-06 | ClinVar | Missense | Lys360Asn | 1080G>C |
| 11:47347661:G:T | 11:47369212 | *MYBPC3* | rs371711564 | VCV000454335 | NC_000011.10:47347660:G:T | NA | NA | 1 | 2.49E-06 | VKGL | Synonymous | Arg281= | 841C>A |
| 11:47347854:C:A | 11:47369405 | *MYBPC3* | rs727503213 | VCV000228869 | NC_000011.10:47347853:C:A | NA | NA | 1 | 2.49E-06 | VKGL | NA | NA | 821+3G>T |
| 12:32821487:D:1 | 12:32974421 | *PKP2* | rs764817683 | VCV000202022 | NC_000012.12:32821487:GGGG:GGG | 1 | NA | NA | 2.49E-06 | VKGL | Frameshift | Lys628fs | 1881del |
| 12:32824163:C:G | 12:32977097 | *PKP2* | rs78897684 | VCV000201989 | NC_000012.12:32824162:C:G | 1 | NA | NA | 2.49E-06 | ClinVar | Splice acceptor | NA | 1557-1G>C |
| 12:32850771:D:4 | 12:33003705 | *PKP2* | rs397516993 | VCV000045020 | NC_000012.12:32850771:TTTGTTT:TTT | 1 | NA | NA | 2.49E-06 | VKGL | Nonsense | Lys456_Gln457insTer | 1369_1372del |
| 12:32869034:G:A | 12:33021968 | *PKP2* | rs754912778 | VCV000201977 | NC_000012.12:32869033:G:A | 1 | NA | NA | 2.49E-06 | ClinVar | Nonsense | Arg355Ter | 1063C>T |
| 12:32878426:G:A | 12:33031360 | *PKP2* | NA | VCV000927573 | NC_000012.12:32878425:G:A | 1 | NA | NA | 2.49E-06 | VKGL | Missense | Pro152Ser | 454C>T |
| 12:32878512:C:T | 12:33031446 | *PKP2* | rs760576804 | VCV000196395 | NC_000012.12:32878511:C:T | 1 | NA | NA | 2.49E-06 | ClinVar | Nonsense | Trp123Ter | 368G>A |
| 12:32879021:G:A | 12:33031955 | *PKP2* | rs121434420 | VCV000006754 | NC_000012.12:32879020:G:A | 1 | NA | NA | 2.49E-06 | ClinVar | Nonsense | Arg79Ter | 235C>T |
| 14:23415652:G:A | 14:23884861 | *MYH7* | rs121913650 | VCV000014118 | NC_000014.9:23415651:G:A | NA | 1 | 1 | 2.49E-06 | ClinVar | Missense | Arg1712Trp | 5134C>T |
| 14:23416129:C:T | 14:23885338 | *MYH7* | rs730880810 | VCV000312894 | NC_000014.9:23416128:C:T | NA | 1 | 1 | 2.49E-06 | VKGL | Missense | Glu1610Lys | 4828G>A |
| 14:23417174:G:A | 14:23886383 | *MYH7* | rs45544633 | VCV000164284 | NC_000014.9:23417173:G:A | NA | 1 | 1 | 2.49E-06 | ClinVar | Missense | Arg1500Trp | 4498C>T |
| 14:23417209:T:C | 14:23886418 | *MYH7* | NA | VCV000920179 | NC_000014.9:23417208:T:C | NA | 1 | 1 | 2.49E-06 | VKGL | Missense | Tyr1488Cys | 4463A>G |
| 14:23418337:C:T | 14:23887546 | *MYH7* | rs1275262402 | VCV000524974 | NC_000014.9:23418336:C:T | NA | 1 | 1 | 2.49E-06 | VKGL | Missense | Glu1348Lys | 4042G>A |
| 14:23422267:C:T | 14:23891476 | *MYH7* | rs587782962 | VCV000155814 | NC_000014.9:23422266:C:T | NA | 1 | 1 | 2.49E-06 | ClinVar | Missense | Arg1053Gln | 3158G>A |
| 14:23423966:C:T | 14:23893175 | *MYH7* | rs886039204 | VCV000264608 | NC_000014.9:23423965:C:T | NA | NA | 1 | 2.49E-06 | VKGL | Missense | Asp955Asn | 2863G>A |
| 14:23426046:G:A | 14:23895255 | *MYH7* | rs727504240 | VCV000177627 | NC_000014.9:23426045:G:A | NA | NA | 1 | 2.49E-06 | VKGL | Missense | Arg694Cys | 2080C>T |
| 14:23427840:C:T | 14:23897049 | *MYH7* | rs564101364 | VCV000264607 | NC_000014.9:23427839:C:T | NA | 1 | 1 | 2.49E-06 | VKGL | Missense | Asp545Asn | 1633G>A |
| 14:23429279:G:A | 14:23898488 | *MYH7* | rs3218714 | VCV000014102 | NC_000014.9:23429278:G:A | NA | 1 | 1 | 2.49E-06 | ClinVar | Missense | Arg403Trp | 1207C>T |
| 14:23431426:A:G | 14:23900635 | *MYH7* | rs397516269 | VCV000043106 | NC_000014.9:23431425:A:G | NA | 1 | 1 | 2.49E-06 | ClinVar | Missense | Ile263Thr | 788T>C |
| 14:23431611:C:T | 14:23900820 | *MYH7* | rs397516261 | VCV000043096 | NC_000014.9:23431610:C:T | NA | 1 | 1 | 2.49E-06 | VKGL | Missense | Val236Ile | 706G>A |
| 15:34791163:C:T | 15:35083364 | *ACTC1* | rs121912673 | VCV000018323 | NC_000015.10:34791162:C:T | NA | 1 | 1 | 2.49E-06 | VKGL | Missense | Arg314His | 941G>A |
| 18:31070776:G:A | 18:28650742 | *DSC2* | rs769022411 | VCV000568186 | NC_000018.10:31070775:G:A | 1 | NA | NA | 2.49E-06 | ClinVar | Nonsense | Gln734Ter | 2200C>T |
| 18:31498297:G:A | 18:29078260 | *DSG2* | rs1568098570 | VCV000567764 | NC_000018.10:31498296:G:A | 1 | NA | NA | 2.49E-06 | ClinVar | Splice donor | NA | 45+1G>A |
| 18:31521244:G:C | 18:29101207 | *DSG2* | rs553299589 | VCV000188450 | NC_000018.10:31521243:G:C | 1 | NA | NA | 2.49E-06 | ClinVar | Splice donor | NA | 523+1G>C |
| 18:31522250:G:A | 18:29102213 | *DSG2* | rs750176752 | VCV000410373 | NC_000018.10:31522249:G:A | 1 | NA | NA | 2.49E-06 | ClinVar | Splice donor | NA | 690+1G>A |
| 18:31524554:A:G | 18:29104517 | *DSG2* | rs121913011 | VCV000016815 | NC_000018.10:31524553:A:G | 1 | NA | NA | 2.49E-06 | ClinVar | Missense | Asn266Ser | 797A>G |
| 18:31538849:C:T | 18:29118812 | *DSG2* | rs794728086 | VCV000199810 | NC_000018.10:31538848:C:T | 1 | NA | NA | 2.49E-06 | ClinVar | Nonsense | Gln584Ter | 1750C>T |
| 18:31538922:I:1 | 18:29118885-29118886 | *DSG2* | rs1039633976 | VCV000585217 | NC_000018.10:31538922:GGG:GGGG | 1 | NA | NA | 2.49E-06 | ClinVar | Frameshift | Leu610fs | 1826dup |
| 19:55154157:C:T | 19:55665525 | *TNNI3* | rs397516347 | VCV000043381 | NC_000019.10:55154156:C:T | NA | 1 | 1 | 2.49E-06 | ClinVar | Missense | Arg141Gln | 422G>A |
| 2:178528367:G:A | 2:179393094 | *TTN* | rs1477669354 | VCV000640886 | NC_000002.12:178528366:G:A | NA | 1 | NA | 2.49E-06 | ClinVar | Nonsense | Arg35762Ter | 107284C>T |
| 2:178528797:G:A | 2:179393524 | *TTN* | rs565675340 | VCV000242530 | NC_000002.12:178528796:G:A | NA | 1 | NA | 2.49E-06 | ClinVar | Nonsense | Arg35652Ter | 106954C>T |
| 2:178529959:C:T | 2:179394686 | *TTN* | rs760915007 | VCV000617573 | NC_000002.12:178529958:C:T | NA | 1 | NA | 2.49E-06 | ClinVar | Splice donor | NA | 106531+1G>A |
| 2:178532100:G:A | 2:179396827 | *TTN* | rs1553488049 | VCV000535030 | NC_000002.12:178532099:G:A | NA | 1 | NA | 2.49E-06 | ClinVar | Nonsense | Arg34839Ter | 104515C>T |
| 2:178532202:G:A | 2:179396929 | *TTN* | rs750519430 | VCV000290707 | NC_000002.12:178532201:G:A | NA | 1 | NA | 2.49E-06 | VKGL | Nonsense | Arg34805Ter | 104413C>T |
| 2:178532844:G:A | 2:179397571 | *TTN* | NA | VCV000934781 | NC_000002.12:178532843:G:A | NA | 1 | NA | 2.49E-06 | ClinVar | Nonsense | Arg34591Ter | 103771C>T |
| 2:178534619:C:T | 2:179399346 | *TTN* | rs869312068 | VCV000223304 | NC_000002.12:178534618:C:T | NA | 1 | NA | 2.49E-06 | ClinVar | Nonsense | Trp33999Ter | 101996G>A |
| 2:178535790:G:A | 2:179400517 | *TTN* | rs1057518195 | VCV000373074 | NC_000002.12:178535789:G:A | NA | 1 | NA | 2.49E-06 | ClinVar | Nonsense | Arg33609Ter | 100825C>T |
| 2:178536357:C:A | 2:179401084 | *TTN* | rs374920916 | VCV000654634 | NC_000002.12:178536356:C:A | NA | 1 | NA | 2.49E-06 | ClinVar | Nonsense | Glu33464Ter | 100390G>T |
| 2:178544357:G:A | 2:179409084 | *TTN* | NA | VCV001067228 | NC_000002.12:178544356:G:A | NA | 1 | NA | 2.49E-06 | VKGL | Nonsense | Arg31958Ter | 95872C>T |
| 2:178546102:A:G | 2:179410829 | *TTN* | rs869320740 | VCV000132133 | NC_000002.12:178546101:A:G | NA | 1 | NA | 2.49E-06 | ClinVar | Missense | Cys31712Arg | 95134T>C |
| 2:178546476:G:A | 2:179411203 | *TTN* | rs869311121 | VCV000223389 | NC_000002.12:178546475:G:A | NA | 1 | NA | 2.49E-06 | ClinVar | Nonsense | Arg31619Ter | 94855C>T |
| 2:178549309:G:A | 2:179414036 | *TTN* | rs794729301 | VCV000202424 | NC_000002.12:178549308:G:A | NA | 1 | NA | 2.49E-06 | ClinVar | Nonsense | Arg30773Ter | 92317C>T |
| 2:178553039:C:T | 2:179417766 | *TTN* | rs1060500457 | VCV000404812 | NC_000002.12:178553038:C:T | NA | 1 | NA | 2.49E-06 | ClinVar | Nonsense | Trp29954Ter | 89861G>A |
| 2:178554094:G:A | 2:179418821 | *TTN* | rs886038916 | VCV000263764 | NC_000002.12:178554093:G:A | NA | 1 | NA | 2.49E-06 | ClinVar | Nonsense | Arg29673Ter | 89017C>T |
| 2:178559309:A:T | 2:179424036 | *TTN* | rs397517735 | VCV000047458 | NC_000002.12:178559308:A:T | NA | 1 | NA | 2.49E-06 | ClinVar | Splice donor | NA | 86821+2T>A |
| 2:178560055:I:1 | 2:179424782-179424783 | *TTN* | rs1285329277 | VCV000519013 | NC_000002.12:178560055:TTTTTT:TTTTTTT | NA | 1 | NA | 2.49E-06 | ClinVar | Frameshift | Ser28693fs | 86076dup |
| 2:178560364:G:A | 2:179425091 | *TTN* | rs748689777 | VCV000488732 | NC_000002.12:178560363:G:A | NA | 1 | NA | 2.49E-06 | ClinVar | Nonsense | Arg28590Ter | 85768C>T |
| 2:178563607:G:A | 2:179428334 | *TTN* | rs1575649368 | VCV000667023 | NC_000002.12:178563606:G:A | NA | 1 | NA | 2.49E-06 | ClinVar | Nonsense | Arg27509Ter | 82525C>T |
| 2:178563892:G:A | 2:179428619 | *TTN* | rs766840243 | VCV000202415 | NC_000002.12:178563891:G:A | NA | 1 | NA | 2.49E-06 | ClinVar | Nonsense | Arg27414Ter | 82240C>T |
| 2:178566838:G:A | 2:179431565 | *TTN* | rs774411587 | VCV000466659 | NC_000002.12:178566837:G:A | NA | 1 | NA | 2.49E-06 | ClinVar | Nonsense | Arg26432Ter | 79294C>T |
| 2:178570158:C:T | 2:179434885 | *TTN* | rs1553602546 | VCV000518953 | NC_000002.12:178570157:C:T | NA | 1 | NA | 2.49E-06 | ClinVar | Nonsense | Trp25325Ter | 75974G>A |
| 2:178570882:G:A | 2:179435609 | *TTN* | rs794729286 | VCV000202406 | NC_000002.12:178570881:G:A | NA | 1 | NA | 2.49E-06 | ClinVar | Nonsense | Arg25084Ter | 75250C>T |
| 2:178573462:D:1 | 2:179438190 | *TTN* | rs727504531 | VCV000178908 | NC_000002.12:178573462:A: | NA | 1 | NA | 2.49E-06 | ClinVar | Frameshift | Asp24224fs | 72669del |
| 2:178577762:C:T | 2:179442489 | *TTN* | NA | VCV000958293 | NC_000002.12:178577761:C:T | NA | 1 | NA | 2.49E-06 | ClinVar | Nonsense | Trp22888Ter | 68664G>A |

| SNP | GRCh37 | Gene | rsID | Accession ClinVar | Canonical SPDI | N ARVC | N DCM | N HCM | MAF | Origin | Molecular Consequence | Amino acid change | Nucleotide change |
|---|---|---|---|---|---|---|---|---|---|---|---|---|---|
| 2:178584552:G:A | 2:179449279 | *TTN* | rs794729280 | VCV000202399 | NC_000002.12:178584551:G:A | NA | 1 | NA | 2.49E-06 | ClinVar | Nonsense | Arg21667Ter | 64999C>T |
| 2:178587418:C:T | 2:179452145 | *TTN* | NA | VCV000948116 | NC_000002.12:178587417:C:T | NA | 1 | NA | 2.49E-06 | ClinVar | Splice acceptor | NA | 63794-1G>A |
| 2:178590230:G:A | 2:179454957 | *TTN* | rs869312112 | VCV000223377 | NC_000002.12:178590229:G:A | NA | 1 | NA | 2.49E-06 | ClinVar | Nonsense | Arg20499Ter | 61495C>T |
| 2:178593566:A:G | 2:179458293 | *TTN* | rs869312054 | VCV000223287 | NC_000002.12:178593565:A:G | NA | 1 | NA | 2.49E-06 | ClinVar | Splice donor | NA | 58732+2T>C |
| 2:178594198:G:A | 2:179458925 | *TTN* | rs768073446 | VCV000432196 | NC_000002.12:178594197:G:A | NA | 1 | NA | 2.49E-06 | VKGL | Nonsense | Arg19399Ter | 58195C>T |
| 2:178595636:G:A | 2:179460363 | *TTN* | NA | VCV000839183 | NC_000002.12:178595635:G:A | NA | 1 | NA | 2.49E-06 | ClinVar | Nonsense | Arg19240Ter | 57718C>T |
| 2:178597751:G:A | 2:179462478 | *TTN* | rs72646831 | VCV000047121 | NC_000002.12:178597750:G:A | NA | 1 | NA | 2.49E-06 | ClinVar | Nonsense | Arg19111Ter | 57331C>T |
| 2:178599145:C:T | 2:179463872 | *TTN* | rs397517624 | VCV000047113 | NC_000002.12:178599144:C:T | NA | 1 | NA | 2.49E-06 | ClinVar | Splice donor | NA | 56647+1G>A |
| 2:178601739:G:A | 2:179466466 | *TTN* | NA | VCV000958943 | NC_000002.12:178601738:G:A | NA | 1 | NA | 2.49E-06 | ClinVar | Nonsense | Arg18451Ter | 55351C>T |
| 2:178604269:G:A | 2:179468996 | *TTN* | rs747236787 | VCV000579797 | NC_000002.12:178604268:G:A | NA | 1 | NA | 2.49E-06 | ClinVar | Nonsense | Arg18140Ter | 54418C>T |
| 2:178605110:G:A | 2:179469837 | *TTN* | rs1553682168 | VCV000466638 | NC_000002.12:178605109:G:A | NA | 1 | NA | 2.49E-06 | ClinVar | Nonsense | Arg18023Ter | 54067C>T |
| 2:178605552:G:A | 2:179470279 | *TTN* | rs753333359 | VCV000534995 | NC_000002.12:178605551:G:A | NA | 1 | NA | 2.49E-06 | ClinVar | Nonsense | Arg17915Ter | 53743C>T |
| 2:178608700:I:4 | 2:179473427-179473428 | *TTN* | rs794729323 | VCV000202450 | NC_000002.12:178608700:TCAATCA:TCAAT | NA | 1 | NA | 2.49E-06 | ClinVar | Nonsense | Glu17437delinsAspTer | 52307_52310dup |
| 2:178610089:C:T | 2:179474816 | *TTN* | rs761807131 | VCV000202384 | NC_000002.12:178610088:C:T | NA | 1 | NA | 2.49E-06 | ClinVar | Splice donor | NA | 51436+1G>A |
| 2:178610090:G:A | 2:179474817 | *TTN* | rs906494713 | VCV000691694 | NC_000002.12:178610089:G:A | NA | 1 | NA | 2.49E-06 | ClinVar | Nonsense | Gln17146Ter | 51436C>T |
| 2:178612115:G:A | 2:179476842 | *TTN* | rs754866489 | VCV000202379 | NC_000002.12:178612114:G:A | NA | 1 | NA | 2.49E-06 | ClinVar | Nonsense | Arg16766Ter | 50296C>T |
| 2:178612355:G:A | 2:179477082 | *TTN* | rs794729265 | VCV000202378 | NC_000002.12:178612354:G:A | NA | 1 | NA | 2.49E-06 | ClinVar | Nonsense | Arg16724Ter | 50170C>T |
| 2:178614226:G:A | 2:179478953 | *TTN* | rs570046043 | VCV000636978 | NC_000002.12:178614225:G:A | NA | 1 | NA | 2.49E-06 | ClinVar | Nonsense | Arg16391Ter | 49171C>T |
| 2:178622684:G:A | 2:179487411 | *TTN* | rs727505350 | VCV000180102 | NC_000002.12:178622683:G:A | NA | 1 | NA | 2.49E-06 | VKGL | Nonsense | Arg14967Ter | 44899C>T |
| 2:178629441:G:A | 2:179494168 | *TTN* | rs770767998 | VCV000223369 | NC_000002.12:178629440:G:A | NA | 1 | NA | 2.49E-06 | VKGL | Nonsense | Arg14762Ter | 44284C>T |
| 2:178630250:G:A | 2:179494977 | *TTN* | rs140743001 | VCV000202367 | NC_000002.12:178630249:G:A | NA | 1 | NA | 2.49E-06 | VKGL | Nonsense | Arg14758Ter | 44272C>T |
| 2:178678746:C:T | 2:179543473 | *TTN* | rs1389908421 | VCV000522785 | NC_000002.12:178678745:C:T | NA | 1 | NA | 2.49E-06 | ClinVar | Splice donor | NA | 33826+1G>A |
| 2:178740125:G:A | 2:179604852 | *TTN* | rs267607158 | VCV000012657 | NC_000002.12:178740124:G:A | NA | 1 | NA | 2.49E-06 | ClinVar | Nonsense | Gln4370Ter | 13108C>T |
| 2:178770483:D:2 | 2:179635211-179635212 | *TTN* | rs869312037 | VCV000223266 | NC_000002.12:178770483:CACA:CA | NA | 1 | NA | 2.49E-06 | ClinVar | Frameshift | Ala2770fs | 8307_8308del |
| 2:178786129:T:A | 2:179650856 | *TTN* | rs1554023044 | VCV000518936 | NC_000002.12:178786128:T:A | NA | 1 | NA | 2.49E-06 | ClinVar | Nonsense | Lys697Ter | 2089A>T |
| 2:219418463:A:G | 2:220283185 | *DES* | rs1057523274 | VCV000388926 | NC_000002.12:219418462:A:G | 1 | 1 | NA | 2.49E-06 | ClinVar | Missense | Met1Val | 1A>G |
| 6:118559037:T:G | 6:118880200 | *PLN* | rs111033560 | VCV000013637 | NC_000006.12:118559036:T:G | 1 | **1** | NA | 2.49E-06 | ClinVar | Nonsense | Leu39Ter | 116T>G |
| 6:7555797:C:T | 6:7556030 | *DSP* | rs768521444 | VCV000451211 | NC_000006.12:7555796:C:T | 1 | 1 | NA | 2.49E-06 | ClinVar | Nonsense | Arg84Ter | 250C>T |
| 6:7558155:C:T | 6:7558388 | *DSP* | NA | VCV000853704 | NC_000006.12:7558154:C:T | 1 | 1 | NA | 2.49E-06 | ClinVar | Nonsense | Arg105Ter | 313C>T |
| 6:7559281:C:T | 6:7559514 | *DSP* | rs397516943 | VCV000044922 | NC_000006.12:7559280:C:T | 1 | 1 | NA | 2.49E-06 | ClinVar | Nonsense | Arg160Ter | 478C>T |
| 6:7568448:I:1 | 6:7568681-7568682 | *DSP* | rs1561687796 | VCV000565816 | NC_000006.12:7568448:AAAA:AAAAA | 1 | NA | NA | 2.49E-06 | ClinVar | Frameshift | Ile428fs | 1282dup |
| 6:7568458:G:T | 6:7568691 | *DSP* | NA | VCV000857798 | NC_000006.12:7568457:G:T | 1 | 1 | NA | 2.49E-06 | ClinVar | Nonsense | Glu430Ter | 1288G>T |
| 6:7568521:C:G | 6:7568754 | *DSP* | NA | VCV000948761 | NC_000006.12:7568520:C:G | NA | 1 | NA | 2.49E-06 | ClinVar | Missense | Arg451Gly | 1351C>G |
| 6:7571554:C:T | 6:7571787 | *DSP* | rs876657638 | VCV000228253 | NC_000006.12:7571553:C:T | 1 | 1 | NA | 2.49E-06 | ClinVar | Nonsense | Gln625Ter | 1873C>T |
| 6:7574797:T:C | 6:7575030 | *DSP* | rs774514264 | VCV000388661 | NC_000006.12:7574796:T:C | 1 | 1 | NA | 2.49E-06 | ClinVar | Splice donor | NA | 2436+2T>C |
| 6:7579385:C:G | 6:7579618 | *DSP* | rs886039178 | VCV000264512 | NC_000006.12:7579384:C:G | 1 | 1 | NA | 2.49E-06 | ClinVar | Nonsense | Tyr1065Ter | 3195C>G |
| 6:7580370:C:T | 6:7580603 | *DSP* | rs140474226 | VCV000162505 | NC_000006.12:7580369:C:T | 1 | 1 | NA | 2.49E-06 | ClinVar | Nonsense | Gln1394Ter | 4180C>T |
| 6:7580721:C:T | 6:7580954 | *DSP* | rs397516940 | VCV000044914 | NC_000006.12:7580720:C:T | 1 | 1 | NA | 2.49E-06 | ClinVar | Nonsense | Gln1511Ter | 4531C>T |
| 6:7581402:C:T | 6:7581635 | *DSP* | rs794728124 | VCV000199890 | NC_000006.12:7581401:C:T | 1 | 1 | NA | 2.49E-06 | ClinVar | Nonsense | Arg1738Ter | 5212C>T |
| 6:7583740:C:T | 6:7583973 | *DSP* | rs777573018 | VCV000199902 | NC_000006.12:7583739:C:T | 1 | 1 | NA | 2.49E-06 | ClinVar | Nonsense | Arg2160Ter | 6478C>T |
| 6:7584904:C:T | 6:7585137 | *DSP* | NA | VCV000984931 | NC_000006.12:7584903:C:T | 1 | 1 | NA | 2.49E-06 | ClinVar | Nonsense | Arg2548Ter | 7642C>T |
| 7:128841304:C:T | 7:128481358 | *FLNC* | rs770606675 | VCV000421215 | NC_000007.14:128841303:C:T | NA | **1** | NA | 2.49E-06 | ClinVar | Nonsense | Arg650Ter | 1948C>T |
| 7:128846396:C:T | 7:128486450 | *FLNC* | rs138193236 | VCV000070588 | NC_000007.14:128846395:C:T | NA | **1** | NA | 2.49E-06 | ClinVar | Nonsense | Arg1354Ter | 4060C>T |
| 7:128853750:C:T | 7:128493804 | *FLNC* | rs1186464414 | VCV000539432 | NC_000007.14:128853749:C:T | NA | **1** | NA | 2.49E-06 | VKGL | Missense | Arg2133Cys | 6397C>T |
| 7:128854661:C:T | 7:128494715 | *FLNC* | rs748416758 | VCV000478129 | NC_000007.14:128854660:C:T | NA | **1** | NA | 2.49E-06 | ClinVar | Nonsense | Arg2326Ter | 6976C>T |

Abbreviations:
ARVC: arrhythmogenic right ventricular cardiomyopathy; DCM: dilated cardiomyopathy; HCM: hypertrophic cardiomyopathy; MAF: minor allele frequency; N: number of carriers.

| Supplementary Table V: Prevalence of all genes per cardiomyopathy | | | | |
|---|---|---|---|---|
| **Cardiomyopathy** | **Gene** | **N** | **Proportion** | **Prevalence** |
| **ARVC** | *DES* | 15 | 4.3 | 7.48E-05 |
| | *DSC2* | 42 | 12.1 | 2.09E-04 |
| | *DSG2* | 31 | 8.9 | 1.55E-04 |
| | *DSP* | 49 | 14.1 | 2.44E-04 |
| | *JUP* | 24 | 6.9 | 1.20E-04 |
| | *PKP2* | 185 | 53.3 | 9.22E-04 |
| | *PLN* | 1 | 0.3 | 4.98E-06 |
| **DCM** | *ACTC1* | 1 | 0.1 | 4.98E-06 |
| | *ACTN2* | 6 | 0.8 | 2.99E-05 |
| | *BAG3* | 15 | 1.9 | 7.48E-05 |
| | *DES* | 49 | 6.1 | 2.44E-04 |
| | *DSP* | 49 | 6.1 | 2.44E-04 |
| | *FLNC* | 56 | 7.0 | 2.79E-04 |
| | *LMNA* | 42 | 5.3 | 2.09E-04 |
| | *MYH7* | 158 | 19.8 | 7.87E-04 |
| | *NEXN* | 4 | 0.5 | 1.99E-05 |
| | *PLN* | 8 | 1.0 | 3.99E-05 |
| | *RBM20* | 5 | 0.6 | 2.49E-05 |
| | *SCN5A* | 59 | 7.4 | 2.94E-04 |
| | *TNNC1* | 7 | 0.9 | 3.49E-05 |
| | *TNNI3* | 35 | 4.4 | 1.74E-04 |
| | *TNNT2* | 32 | 4.0 | 1.59E-04 |
| | *TPM1* | 2 | 0.3 | 9.97E-06 |
| | *TTN* | 272 | 34.0 | 1.36E-03 |
| **HCM** | *ACTC1* | 1 | 0.1 | 4.98E-06 |
| | *CSRP3* | 27 | 2.0 | 1.35E-04 |
| | *JPH2* | 6 | 0.4 | 2.99E-05 |
| | *MYBPC3* | 723 | 53.6 | 3.60E-03 |
| | *MYH7* | 232 | 17.2 | 1.16E-03 |
| | *MYL2* | 21 | 1.6 | 1.05E-04 |
| | *MYL3* | 1 | 0.1 | 4.98E-06 |
| | *TNNC1* | 7 | 0.5 | 3.49E-05 |
| | *TNNI3* | 50 | 3.7 | 2.49E-04 |
| | *TNNT2* | 274 | 20.3 | 1.37E-03 |
| | *TPM1* | 6 | 0.4 | 2.99E-05 |

Abbreviations:
*ACTC1: Actin Alpha Cardiac Muscle 1; ACTN2: Alpha-actinin 2;*
*ARVC: Arrhythmogenic right ventricular cardiomyopathy; BAG3: BAG Cochaperone 3;*
*CSRP3: Cysteine And Glycine Rich Protein 3; DCM: Dilated cardiomyopathy; DES: Desmin;*
*DSC2: Desmocollin 2; DSG2: Desmoglein 2; DSG2: Desmoglein 2; DSP: desmoplakin;*
*FLNC: Filamin-C; HCM: Hypertrophic cardiomyopathy; JPH2: Junctophilin 2;*
*JUP: Junction Plakoglobin; LMNA: Lamin A/C; MYBPC3: Myosin Binding Protein C3;*
*MYH7: Myosin Heavy Chain 7; MYL2: Myosin Light Chain 2; MYL3: Myosin Light Chain 3;*
*N: Number of individuals; NEXN: Nexilin F-Actin Binding Protein; PKP2: Plakophilin 2;*
*PLN: phospholamban; RBM20: RNA Binding Motif Protein 20;*
*SCN5A: Sodium Voltage-Gated Channel Alpha Subunit 5;*
*TNNC1: Troponin C1, Slow Skeletal And Cardiac Type; TNNI3: Troponin I3, Cardiac Type;*
*TNNT2: Troponin T2, Cardiac Type; TPM1: Tropomyosin 1; TTN: Titin.*

| Supplementary Table VI: Prevalence of variants associated with the inherited cardiomyopathies | | | |
|---|---|---|---|
| **Cardiomyopathy** | **Prevalence with overlapping genes** | **Prevalence without overlapping genes** | **Previously reported prevalence range** |
| ARVC | 1:578 | 1:712 | 1:143 - 1:1,706[13-15] |
| DCM | 1:251 | 1:289 (ARVC overlap) / 1:354 (HCM overlap) | 1:33 - 1:526[16, 17] |
| HCM | 1:149 | 1:260 | 1:164[19] |

Abbreviations:
ARVC: arrhythmogenic right ventricular cardiomyopathy; DCM: dilated cardiomyopathy; HCM: hypertrophic cardiomyopathy.

**Supplementary Table VII: Results of Fisher's Exact tests**

| | ARVC G+ vs G- | | | | DCM G+ vs G- | | | | HCM G+ vs G- | | | |
|---|---|---|---|---|---|---|---|---|---|---|---|---|
| | OR | 95% LCI | 95% UCI | p-value | OR | 95% LCI | 95% UCI | p-value | OR | 95% LCI | 95% UCI | p-value |
| CARDIOVASCULAR RISK FACTORS | | | | | | | | | | | | |
| Diabetes | 1.112 | 0.755 | 1.592 | 0.570 | 0.833 | 0.626 | 1.091 | 0.200 | 1.280 | 1.061 | 1.537 | 0.008 |
| Hypertension | 0.962 | 0.760 | 1.212 | 0.774 | 1.072 | 0.919 | 1.248 | 0.374 | 1.045 | 0.925 | 1.179 | 0.482 |
| Hypercholesterolaemia | 1.031 | 0.795 | 1.326 | 0.799 | 1.120 | 0.946 | 1.322 | 0.185 | 1.181 | 1.036 | 1.345 | 0.011 |
| Ever Smoked | 1.223 | 0.981 | 1.525 | 0.068 | 1.222 | 1.055 | 1.416 | 0.007 | 0.956 | 0.849 | 1.075 | 0.461 |
| Family heart disease | 1.318 | 1.058 | 1.643 | 0.012 | 1.119 | 0.966 | 1.296 | 0.130 | 1.066 | 0.949 | 1.197 | 0.280 |
| CARDIAC DISEASE/OUTCOME | | | | | | | | | | | | |
| Cardiac problem | 2.112 | 0.416 | 6.672 | 0.183 | 0.912 | 0.180 | 2.868 | 1.000 | 0.903 | 0.278 | 2.289 | 1.000 |
| Heart failure* | 1.432 | 0.639 | 2.812 | 0.305 | 2.534 | 1.708 | 3.671 | 5.05E-06 | 1.352 | 0.899 | 1.977 | 0.135 |
| Cardiomyopathy* | 2.341 | 0.460 | 7.452 | 0.150 | 7.590 | 4.242 | 13.283 | 6.94E-11 | 5.495 | 3.206 | 9.306 | 8.76E-10 |
| Phenotype positive† | 1.325 | 0.351 | 3.547 | 0.550 | 3.664 | 2.236 | 5.813 | 4.88E-07 | 3.033 | 1.979 | 4.560 | 5.76E-07 |
| Dilated cardiomyopathy* | 4.122 | 0.453 | 18.056 | 0.099 | 8.090 | 3.078 | 20.130 | 2.08E-05 | 0.529 | 0.013 | 3.481 | 1.000 |
| Hypertrophic cardiomyopathy* | 3.599 | 0.081 | 26.974 | 0.265 | 10.989 | 3.383 | 34.746 | 4.60E-05 | 18.775 | 7.903 | 49.429 | 3.41E-13 |
| Ventricular arrhythmias | 6.198 | 2.297 | 14.376 | 3.27E-04 | 4.974 | 2.392 | 9.752 | 1.93E-05 | 1.801 | 0.717 | 3.987 | 0.143 |
| Atrial arrhythmias | 1.054 | 0.415 | 2.239 | 0.841 | 2.273 | 1.518 | 3.314 | 8.18E-05 | 1.247 | 0.826 | 1.830 | 0.250 |
| Heart arrhythmia | 3.231 | 1.128 | 7.573 | 0.015 | 2.796 | 1.356 | 5.321 | 0.003 | 0.547 | 0.144 | 1.488 | 0.310 |
| Chronic ischemic heart disease* | 1.431 | 0.971 | 2.052 | 0.059 | 1.281 | 0.981 | 1.652 | 0.058 | 0.947 | 0.748 | 1.186 | 0.695 |
| Acute myocardial infarction | 1.467 | 0.801 | 2.494 | 0.151 | 1.134 | 0.730 | 1.697 | 0.519 | 0.892 | 0.610 | 1.270 | 0.606 |
| Cardiac arrest | 0.000 | 0.000 | 3.299 | 0.630 | 2.209 | 0.756 | 5.343 | 0.118 | 1.090 | 0.332 | 2.808 | 0.804 |
| Angina pectoris | 1.497 | 0.835 | 2.506 | 0.120 | 1.206 | 0.795 | 1.772 | 0.344 | 1.344 | 0.987 | 1.803 | 0.049 |
| Conduction disorders | 1.535 | 0.645 | 3.137 | 0.260 | 1.497 | 0.859 | 2.464 | 0.136 | 1.281 | 0.807 | 1.960 | 0.242 |
| Valvular disease | 1.322 | 0.645 | 2.439 | 0.373 | 1.958 | 1.335 | 2.801 | 4.54E-04 | 1.269 | 0.883 | 1.782 | 0.163 |
| Congenital heart disease | 2.059 | 0.237 | 8.219 | 0.267 | 1.337 | 0.260 | 4.342 | 0.499 | 1.059 | 0.269 | 3.033 | 0.788 |
| Pulmonary obstructive disease | 1.490 | 0.940 | 2.266 | 0.078 | 1.198 | 0.860 | 1.634 | 0.240 | 0.848 | 0.629 | 1.125 | 0.280 |
| Cardiovascular death | 1.771 | 0.860 | 3.286 | 0.100 | 1.673 | 1.038 | 2.588 | 0.030 | 0.733 | 0.423 | 1.197 | 0.268 |
| All-cause mortality | 1.068 | 0.629 | 1.713 | 0.712 | 1.388 | 1.023 | 1.852 | 0.032 | 0.890 | 0.668 | 1.169 | 0.428 |

* Used to define P+, therefore not included in some tests.
† Defined as diagnosis of cardiomyopathy, DCM, HCM or heart failure, in absence of chronic ischemic heart disease.

Abbreviations:
ARVC: arrhythmogenic right ventricular cardiomyopathy; DCM: dilated cardiomyopathy;
G+: carriers of likely pathogenic and pathogenic variants associated with one of the cardiomyopathies; HCM: hypertrophic cardiomyopathy;
LCI: lower limit confidence interval; UCI: upper limit confidence interval.

**Supplementary Table VII: Results of Fisher's Exact tests**

| | strict HCM G+ vs G- | | | | ARVC G+P- vs G-P- | | | | DCM G+P- vs G-P- | | | |
|---|---|---|---|---|---|---|---|---|---|---|---|---|
| | OR | 95% LCI | 95% UCI | p-value | OR | 95% LCI | 95% UCI | p-value | OR | 95% LCI | 95% UCI | p-value |
| CARDIOVASCULAR RISK FACTORS | | | | | | | | | | | | |
| Diabetes | 0.802 | 0.601 | 1.055 | 0.124 | 1.108 | 0.749 | 1.594 | 0.566 | 0.860 | 0.645 | 1.130 | 0.296 |
| Hypertension | 1.093 | 0.938 | 1.273 | 0.246 | 0.954 | 0.752 | 1.205 | 0.728 | 1.032 | 0.882 | 1.207 | 0.694 |
| Hypercholesterolaemia | 1.006 | 0.846 | 1.193 | 0.932 | 1.006 | 0.772 | 1.298 | 0.949 | 1.081 | 0.909 | 1.281 | 0.361 |
| Ever Smoked | 1.160 | 1.000 | 1.344 | 0.048 | 1.228 | 0.983 | 1.533 | 0.066 | 1.233 | 1.062 | 1.432 | 0.006 |
| Family heart disease | 1.168 | 1.008 | 1.352 | 0.035 | 1.286 | 1.031 | 1.605 | 0.024 | 1.134 | 0.976 | 1.316 | 0.099 |
| CARDIAC DISEASE/OUTCOME | | | | | | | | | | | | |
| Cardiac problem | 1.216 | 0.315 | 3.369 | 0.575 | 2.227 | 0.438 | 7.061 | 0.166 | 0.981 | 0.194 | 3.097 | 1.000 |
| Heart failure* | 1.733 | 1.086 | 2.661 | 0.015 | NA | NA | NA | NA | NA | NA | NA | NA |
| Cardiomyopathy* | 8.647 | 4.962 | 14.841 | 4.40E-13 | NA | NA | NA | NA | NA | NA | NA | NA |
| Phenotype positive† | 4.727 | 3.028 | 7.216 | 8.15E-11 | NA | NA | NA | NA | NA | NA | NA | NA |
| Dilated cardiomyopathy* | 0.000 | 0.000 | 3.756 | 0.619 | NA | NA | NA | NA | NA | NA | NA | NA |
| Hypertrophic cardiomyopathy* | 30.258 | 12.586 | 79.971 | 3.68E-16 | NA | NA | NA | NA | NA | NA | NA | NA |
| Ventricular arrhythmias | 3.038 | 1.208 | 6.738 | 0.010 | 5.846 | 1.976 | 14.398 | 0.001 | 3.426 | 1.352 | 7.682 | 0.005 |
| Atrial arrhythmias | 1.650 | 1.035 | 2.530 | 0.025 | 1.176 | 0.462 | 2.504 | 0.672 | 2.117 | 1.357 | 3.194 | 0.001 |
| Heart arrhythmia | 0.690 | 0.138 | 2.136 | 0.798 | 2.797 | 0.866 | 7.018 | 0.042 | 2.471 | 1.115 | 4.941 | 0.013 |
| Chronic ischemic heart disease* | 0.885 | 0.649 | 1.187 | 0.477 | NA | NA | NA | NA | NA | NA | NA | NA |
| Acute myocardial infarction | 0.917 | 0.562 | 1.425 | 0.828 | 1.476 | 0.806 | 2.511 | 0.149 | 1.121 | 0.715 | 1.689 | 0.586 |
| Cardiac arrest | 1.467 | 0.377 | 4.128 | 0.527 | 0.000 | 0.000 | 3.415 | 0.628 | 1.938 | 0.589 | 5.017 | 0.195 |
| Angina pectoris | 1.288 | 0.860 | 1.873 | 0.175 | 1.511 | 0.843 | 2.531 | 0.117 | 1.244 | 0.819 | 1.828 | 0.244 |
| Conduction disorders | 1.241 | 0.674 | 2.126 | 0.454 | 1.650 | 0.693 | 3.379 | 0.165 | 1.364 | 0.740 | 2.342 | 0.274 |
| Valvular disease | 1.517 | 0.987 | 2.253 | 0.045 | 1.332 | 0.624 | 2.527 | 0.351 | 1.662 | 1.072 | 2.491 | 0.017 |
| Congenital heart disease | 1.335 | 0.259 | 4.337 | 0.499 | 2.313 | 0.264 | 9.341 | 0.229 | 1.020 | 0.117 | 4.105 | 1.000 |
| Pulmonary obstructive disease | 0.903 | 0.620 | 1.279 | 0.611 | 1.575 | 0.993 | 2.398 | 0.040 | 1.090 | 0.762 | 1.522 | 0.600 |
| Cardiovascular death | 0.684 | 0.321 | 1.294 | 0.328 | 1.916 | 0.929 | 3.562 | 0.054 | 1.454 | 0.849 | 2.358 | 0.153 |
| All-cause mortality | 1.097 | 0.783 | 1.505 | 0.562 | 1.134 | 0.668 | 1.820 | 0.611 | 1.277 | 0.920 | 1.738 | 0.123 |

* Used to define P+, therefore not included in some tests.
† Defined as diagnosis of cardiomyopathy, DCM, HCM or heart failure, in absence of chronic ischemic heart disease.

Abbreviations:
ARVC: arrhythmogenic right ventricular cardiomyopathy; DCM: dilated cardiomyopathy;
G+: carriers of likely pathogenic and pathogenic variants associated with one of the cardiomyopathies; HCM: hypertrophic cardiomyopathy;
LCI: lower limit confidence interval; UCI: upper limit confidence interval.

| Supplementary Table VII: Results of Fisher's Exact tests | | | | | | | | |
|---|---|---|---|---|---|---|---|---|
| | HCM G+P- vs G-P- | | | | strict HCM G+P- vs G-P- | | | |
| | OR | 95% LCI | 95% UCI | p-value | OR | 95% LCI | 95% UCI | p-value |
| **CARDIOVASCULAR RISK FACTORS** | | | | | | | | |
| Diabetes | 1.301 | 1.075 | 1.566 | 0.006 | 0.806 | 0.599 | 1.067 | 0.148 |
| Hypertension | 1.022 | 0.903 | 1.155 | 0.733 | 1.051 | 0.897 | 1.228 | 0.528 |
| Hypercholesterolaemia | 1.179 | 1.032 | 1.344 | 0.014 | 0.991 | 0.830 | 1.179 | 0.965 |
| Ever Smoked | 0.968 | 0.859 | 1.090 | 0.591 | 1.187 | 1.022 | 1.380 | 0.023 |
| Family heart disease | 1.068 | 0.949 | 1.200 | 0.274 | 1.175 | 1.012 | 1.364 | 0.032 |
| **CARDIAC DISEASE/OUTCOME** | | | | | | | | |
| Cardiac problem | 0.967 | 0.297 | 2.460 | 1.000 | 1.320 | 0.342 | 3.673 | 0.550 |
| Heart failure* | NA | NA | NA | NA | NA | NA | NA | NA |
| Cardiomyopathy* | NA | NA | NA | NA | NA | NA | NA | NA |
| Phenotype positive† | NA | NA | NA | NA | NA | NA | NA | NA |
| Dilated cardiomyopathy* | NA | NA | NA | NA | NA | NA | NA | NA |
| Hypertrophic cardiomyopathy* | NA | NA | NA | NA | NA | NA | NA | NA |
| Ventricular arrhythmias | 1.005 | 0.257 | 2.859 | 1.000 | 1.718 | 0.438 | 4.892 | 0.306 |
| Atrial arrhythmias | 1.143 | 0.723 | 1.741 | 0.504 | 1.431 | 0.836 | 2.319 | 0.156 |
| Heart arrhythmia | 0.579 | 0.152 | 1.577 | 0.403 | 0.741 | 0.148 | 2.296 | 0.797 |
| Chronic ischemic heart disease* | NA | NA | NA | NA | NA | NA | NA | NA |
| Acute myocardial infarction | 0.860 | 0.581 | 1.235 | 0.487 | 0.862 | 0.516 | 1.366 | 0.583 |
| Cardiac arrest | 1.143 | 0.348 | 2.954 | 0.799 | 1.561 | 0.401 | 4.405 | 0.339 |
| Angina pectoris | 1.378 | 1.011 | 1.849 | 0.038 | 1.341 | 0.894 | 1.950 | 0.136 |
| Conduction disorders | 1.071 | 0.632 | 1.724 | 0.713 | 0.910 | 0.425 | 1.734 | 1.000 |
| Valvular disease | 1.145 | 0.765 | 1.665 | 0.486 | 1.245 | 0.751 | 1.966 | 0.313 |
| Congenital heart disease | 1.207 | 0.305 | 3.503 | 0.770 | 1.545 | 0.298 | 5.079 | 0.451 |
| Pulmonary obstructive disease | 0.844 | 0.619 | 1.130 | 0.266 | 0.898 | 0.606 | 1.291 | 0.659 |
| Cardiovascular death | 0.669 | 0.365 | 1.141 | 0.163 | 0.531 | 0.210 | 1.125 | 0.105 |
| All-cause mortality | 0.863 | 0.638 | 1.148 | 0.338 | 1.061 | 0.742 | 1.481 | 0.729 |

* Used to define P+, therefore not included in some tests.
† Defined as diagnosis of cardiomyopathy, DCM, HCM or heart failure, in absence of chronic ischemic heart disease.

Abbreviations:
ARVC: arrhythmogenic right ventricular cardiomyopathy; DCM: dilated cardiomyopathy;
G+: carriers of likely pathogenic and pathogenic variants associated with one of the cardiomyopathies; HCM: hypertrophic cardiomyopathy;
LCI: lower limit confidence interval; UCI: upper limit confidence interval.

| Supplementary Table VIII: P-values of Mann-Whitney U tests | ARVC G+ vs G- | DCM G+ vs G- | HCM G+ vs G- | strict HCM* G+ vs G- |
|---|---|---|---|---|
| **CARDIOVASCULAR RISK FACTORS** | | | | |
| **BMI** | 0.990 | 0.812 | 0.271 | 0.621 |
| **Mean systolic blood pressure** | 0.774 | 0.901 | 0.258 | 0.508 |
| **Mean diastolic blood pressure** | 0.341 | 0.734 | 0.997 | 0.776 |
| **Total cholesterol** | 0.369 | 0.844 | 0.242 | 0.666 |
| **HDL** | 0.941 | 0.357 | 0.076 | 0.161 |
| **LDL** | 0.214 | 0.898 | 0.484 | 0.778 |
| **MET minutes per week for walking** | 0.545 | 0.055 | 0.233 | 0.017 |
| **MET minutes per week for moderate activity** | 0.950 | 0.913 | 0.578 | 0.155 |
| **MET minutes per week for vigorous activity** | 0.352 | 0.963 | 0.350 | 0.589 |
| **Total MET minutes per week** | 0.278 | 0.619 | 0.980 | 0.052 |
| **ECG MEASUREMENTS** | | | | |
| **P duration** | NA | NA | NA | NA |
| **P axis** | NA | NA | NA | NA |
| **PQ interval** | NA | NA | NA | NA |
| **QRS duration** | NA | NA | NA | NA |
| **R axis** | NA | NA | NA | NA |
| **QTC interval** | NA | NA | NA | NA |
| **T axis** | NA | NA | NA | NA |
| **CMR MEASUREMENTS** | | | | |
| **RVEDVi** | NA | NA | NA | NA |
| **RVESVi** | NA | NA | NA | NA |
| **RVSV** | NA | NA | NA | NA |
| **RVSVi** | NA | NA | NA | NA |
| **RVEF** | NA | NA | NA | NA |
| **RVPER** | NA | NA | NA | NA |
| **RVPFR** | NA | NA | NA | NA |
| **RVPAFR** | NA | NA | NA | NA |
| **LVEDVi** | NA | NA | NA | NA |
| **LVESVi** | NA | NA | NA | NA |
| **LVSV** | NA | NA | NA | NA |
| **LVSVi** | NA | NA | NA | NA |
| **LVEF** | NA | NA | NA | NA |
| **LVPER** | NA | NA | NA | NA |
| **LVPFR** | NA | NA | NA | NA |
| **LVPAFR** | NA | NA | NA | NA |
| **LVEDMi** | NA | NA | NA | NA |
| **LVMVR** | NA | NA | NA | NA |
| **LVEDV/RVEDV** | NA | NA | NA | NA |
| **LVESV/RVESV** | NA | NA | NA | NA |
| **peakEcc** | NA | NA | NA | NA |
| **TPKEcc** | NA | NA | NA | NA |
| **peakEll2Ch** | NA | NA | NA | NA |
| **TPKEll2Ch** | NA | NA | NA | NA |
| **peakEll4Ch** | NA | NA | NA | NA |
| **TPKEll4Ch** | NA | NA | NA | NA |
| **Wall thickness segment 1** | NA | NA | NA | NA |
| **Wall thickness segment 2** | NA | NA | NA | NA |
| **Wall thickness segment 3** | NA | NA | NA | NA |
| **Wall thickness segment 4** | NA | NA | NA | NA |
| **Wall thickness segment 5** | NA | NA | NA | NA |
| **Wall thickness segment 6** | NA | NA | NA | NA |
| **Wall thickness segment 7** | NA | NA | NA | NA |
| **Wall thickness segment 8** | NA | NA | NA | NA |
| **Wall thickness segment 9** | NA | NA | NA | NA |
| **Wall thickness segment 10** | NA | NA | NA | NA |
| **Wall thickness segment 11** | NA | NA | NA | NA |
| **Wall thickness segment 12** | NA | NA | NA | NA |
| **Wall thickness segment 13** | NA | NA | NA | NA |
| **Wall thickness segment 14** | NA | NA | NA | NA |
| **Wall thickness segment 15** | NA | NA | NA | NA |
| **Wall thickness segment 16** | NA | NA | NA | NA |
| **Global wall thickness** | NA | NA | NA | NA |
| **Septal wall thickness** | NA | NA | NA | NA |
| **Maximum wall thickness** | NA | NA | NA | NA |

* strict HCM group: HCM group after excluding carriers of the 3628-41_3628-17del *MYBPC3* and the Arg278Cys 862C>T *TNNT2* variant.

Abbreviations:
ARVC: arrythmogenic right ventricular cardiomyopathy; BMI: body mass index; CMR: cardiac magnetic resonance imaging; DCM: dilated cardiomyopathy; ECG: Electrocardiography; EDVi: indexed end-diastolic volume; EDMi: indexed end-diastolic mass; EF: ejection fraction; ESVi: indexed end-systolic volume; G+: carriers of likely pathogenic and pathogenic variants associated with one of the cardiomyopathies; HCM: hypertrophic cardiomyopathy; HDL: high-density lipoprotein; LDL: low-density lipoprotein; LV: left ventricular; MET: metabolic equivalent of task; MVR: mass to volume ratio; PAFR: peak atrial filling rate; peakEcc: peak circumferential strain; peakEll2Ch: longitudinal strain analyzed in 2-chamber view; peakEll4Ch: longitudinal strain analyzed in 4-chamber view; PER: peak ejection rate; PFR: peak filling rate; RV: right ventricular; SVi: indexed stroke volume; TPKEcc: global time to peak circumferential strain; TPKEll2Ch: global time to longitudinal strain analyzed in 2-chamber view; TPKEll4Ch: global time to longitudinal strain analyzed in 4-chamber view.

**Supplementary Table VIII: P-values of Mann-Whitney U tests**

| | ARVC G+P- vs G-P- | DCM G+P- vs G-P- | HCM G+P- vs G-P- | strict HCM* G+P- vs G-P- |
|---|---|---|---|---|
| **CARDIOVASCULAR RISK FACTORS** | | | | |
| **BMI** | 0.945 | 0.800 | 0.317 | 0.712 |
| **Mean systolic blood pressure** | 0.917 | 0.913 | 0.257 | 0.608 |
| **Mean diastolic blood pressure** | 0.367 | 0.760 | 0.934 | 0.737 |
| **Total cholesterol** | 0.381 | 0.780 | 0.211 | 0.808 |
| **HDL** | 0.999 | 0.471 | 0.070 | 0.161 |
| **LDL** | 0.223 | 0.991 | 0.404 | 0.991 |
| **MET minutes per week for walking** | 0.500 | 0.084 | 0.189 | 0.010 |
| **MET minutes per week for moderate activity** | 0.989 | 0.965 | 0.605 | 0.137 |
| **MET minutes per week for vigorous activity** | 0.434 | 0.885 | 0.379 | 0.557 |
| **Total MET minutes per week** | 0.290 | 0.722 | 0.943 | 0.038 |
| **ECG MEASUREMENTS** | | | | |
| **P duration** | 0.315 | 0.304 | 0.997 | 0.999 |
| **P axis** | 0.477 | 0.162 | 0.085 | 0.179 |
| **PQ interval** | 0.617 | 0.989 | 0.527 | 0.904 |
| **QRS duration** | 0.385 | 0.043 | 0.445 | 0.436 |
| **R axis** | 0.208 | 0.156 | 0.868 | 0.699 |
| **QTC interval** | 0.255 | 0.422 | 0.300 | 0.270 |
| **T axis** | 0.818 | 0.572 | 0.074 | 0.128 |
| **CMR MEASUREMENTS** | | | | |
| **RVEDVi** | 0.780 | 0.058 | 0.177 | 0.722 |
| **RVESVi** | 0.707 | 0.287 | 0.051 | 0.118 |
| **RVSV** | 0.713 | 0.071 | 0.910 | 0.140 |
| **RVSVi** | 0.546 | 0.155 | 0.872 | 0.106 |
| **RVEF** | 0.950 | 0.765 | 0.025 | 0.015 |
| **RVPER** | 0.869 | 0.038 | 0.711 | 0.615 |
| **RVPFR** | 0.908 | 0.120 | 0.064 | 0.249 |
| **RVPAFR** | 0.661 | 0.385 | 0.192 | 0.025 |
| **LVEDVi** | 0.060 | 0.125 | 0.378 | 0.444 |
| **LVESVi** | 0.414 | 0.032 | 0.276 | 0.460 |
| **LVSV** | 0.100 | 0.509 | 0.889 | 0.214 |
| **LVSVi** | 0.052 | 0.430 | 0.921 | 0.318 |
| **LVEF** | 0.452 | 0.009 | 0.366 | 0.607 |
| **LVPER** | 0.465 | 0.023 | 0.190 | 0.420 |
| **LVPFR** | 0.114 | 0.436 | 0.567 | 0.485 |
| **LVPAFR** | 0.670 | 0.412 | 0.659 | 0.189 |
| **LVEDMi** | 0.800 | 0.738 | 0.928 | 0.188 |
| **LVMVR** | 0.295 | 0.061 | 0.784 | 0.559 |
| **LVEDV/RVEDV** | 0.360 | 0.001 | 0.533 | 0.747 |
| **LVESV/RVESV** | 0.904 | 0.000 | 0.585 | 0.027 |
| **peakEcc** | 0.319 | 0.107 | 0.643 | 0.812 |
| **TPKEcc** | 0.555 | 0.155 | 0.723 | 0.850 |
| **peakEll2Ch** | 0.751 | 0.019 | 0.751 | 0.179 |
| **TPKEll2Ch** | 0.616 | 0.701 | 0.978 | 0.314 |
| **peakEll4Ch** | 0.483 | 0.009 | 0.079 | 0.286 |
| **TPKEll4Ch** | 0.941 | 0.708 | 0.227 | 0.403 |
| **Wall thickness segment 1** | 0.211 | 0.439 | 0.740 | 0.029 |
| **Wall thickness segment 2** | 0.446 | 0.022 | 0.160 | 0.155 |
| **Wall thickness segment 3** | 0.268 | 0.144 | 0.450 | 0.254 |
| **Wall thickness segment 4** | 0.020 | 0.626 | 0.460 | 0.919 |
| **Wall thickness segment 5** | 0.110 | 0.132 | 0.189 | 0.745 |
| **Wall thickness segment 6** | 0.736 | 0.363 | 0.978 | 0.503 |
| **Wall thickness segment 7** | 0.234 | 0.435 | 0.539 | 0.826 |
| **Wall thickness segment 8** | 0.622 | 0.251 | 0.348 | 0.753 |
| **Wall thickness segment 9** | 0.087 | 0.438 | 0.850 | 0.303 |
| **Wall thickness segment 10** | 0.035 | 0.508 | 0.789 | 0.220 |
| **Wall thickness segment 11** | 0.083 | 0.484 | 0.482 | 0.502 |
| **Wall thickness segment 12** | 0.237 | 0.108 | 0.361 | 0.943 |
| **Wall thickness segment 13** | 0.974 | 0.828 | 0.987 | 0.575 |
| **Wall thickness segment 14** | 0.832 | 0.243 | 0.479 | 0.869 |
| **Wall thickness segment 15** | 0.988 | 0.277 | 0.571 | 0.748 |
| **Wall thickness segment 16** | 0.938 | 0.423 | 0.836 | 0.333 |
| **Global wall thickness** | 0.159 | 0.232 | 0.961 | 0.270 |
| **Septal wall thickness** | 0.229 | 0.071 | 0.523 | 0.174 |
| **Maximum wall thickness** | 0.210 | 0.621 | 0.166 | 0.008 |

* strict HCM group: HCM group after excluding carriers of the 3628-41_3628-17del *MYBPC3* and the Arg278Cys 862C>T *TNNT2* variant.

Abbreviations:
ARVC: arrythmogenic right ventricular cardiomyopathy; BMI: body mass index; CMR: cardiac magnetic resonance imaging; DCM: dilated cardiomyopathy; ECG: Electrocardiography; EDVi: indexed end-diastolic volume; EDMi: indexed end-diastolic mass; EF: ejection fraction; ESVi: indexed end-systolic volume; G+: carriers of likely pathogenic and pathogenic variants associated with one of the cardiomyopathies; HCM: hypertrophic cardiomyopathy; HDL: high-density lipoprotein; LDL: low-density lipoprotein; LV: left ventricular; MET: metabolic equivalent of task; MVR: mass to volume ratio; PAFR: peak atrial filling rate; peakEcc: peak circumferential strain; peakEll2Ch: longitudinal strain analyzed in 2-chamber view; peakEll4Ch: longitudinal strain analyzed in 4-chamber view; PER: peak ejection rate; PFR: peak filling rate; RV: right ventricular; SVi: indexed stroke volume; TPKEcc: global time to peak circumferential strain; TPKEll2Ch: global time to longitudinal strain analyzed in 2-chamber view; TPKEll4Ch: global time to longitudinal strain analyzed in 4-chamber view.

| Supplementary Table IX: Extensive baseline table of P- CMR participants | | | | | |
|---|---|---|---|---|---|
| | ARVC G+ | DCM G+ | HCM G+ | Controls G- | Missing |
| n | 33 | 87 | 130 | 986 | |
| **Sex = Female (%)** | 19 (57.6) | 46 (52.9) | 62 (47.7) | 486 (49.3) | 0 |
| **Age (median [IQR])** | 54.00 [50.00, 61.00] | 55.00 [50.00, 59.50] | 54.00 [48.00, 60.00] | 55.00 [49.00, 60.00] | 0 |
| **Ethnicity (%)** | | | | | 0 |
| Asian | 0 (0.0) | 1 (1.1) | 18 (13.8) | 76 (7.7) | |
| Black | 0 (0.0) | 1 (1.1) | 2 (1.5) | 2 (0.2) | |
| Chinese | 1 (3.0) | 1 (1.1) | 1 (0.8) | 9 (0.9) | |
| Mixed | 0 (0.0) | 1 (1.1) | 5 (3.8) | 15 (1.5) | |
| Other | 0 (0.0) | 0 (0.0) | 1 (0.8) | 5 (0.5) | |
| White | 32 (97.0) | 83 (95.4) | 103 (79.2) | 879 (89.1) | |
| CARDIOVASCULAR RISK FACTORS | | | | | |
| **BMI (median [IQR])** | 26.10 [24.23, 28.72] | 26.27 [23.68, 29.11] | 26.07 [23.82, 28.82] | 25.84 [23.58, 28.62] | 0 |
| **Diabetes (%)** | 2 (6.1) | 7 (8.0) | 12 (9.2) | 78 (7.9) | 0 |
| **Hypertension (%)** | 6 (18.2) | 27 (31.0) | 42 (32.3) | 309 (31.3) | 0 |
| **Mean systolic blood pressure (median [IQR])** | 131.00 [117.50, 141.00] | 134.50 [120.75, 146.75] | 133.00 [122.00, 146.50] | 133.50 [122.00, 146.00] | 0.2 |
| **Mean diastolic blood pressure (median [IQR])** | 79.50 [73.00, 86.50] | 80.25 [74.50, 87.38] | 81.50 [76.00, 87.50] | 81.00 [74.50, 87.00] | 0.2 |
| **Hypercholesterolaemia (%)** | 9 (27.3) | 26 (29.9) | 45 (34.6) | 269 (27.3) | 0 |
| **Total cholesterol (median [IQR])** | 5.39 [4.92, 6.09] | 5.51 [4.76, 6.34] | 5.66 [4.85, 6.48] | 5.63 [4.94, 6.44] | 3.5 |
| **HDL (median [IQR])** | 1.41 [1.12, 1.63] | 1.42 [1.19, 1.61] | 1.38 [1.14, 1.62] | 1.40 [1.18, 1.71] | 8.8 |
| **LDL (median [IQR])** | 3.31 [3.00, 4.02] | 3.45 [2.87, 4.13] | 3.58 [2.89, 4.24] | 3.50 [3.00, 4.11] | 3.7 |
| **Ever Smoked (%)** | 13 (39.4) | 45 (51.7) | 53 (40.8) | 397 (40.3) | 0 |
| **MET minutes per week for walking (median [IQR])** | 693.00 [309.38, 1,386.00] | 462.00 [198.00, 1,188.00] | 495.00 [247.50, 1,039.50] | 594.00 [247.50, 1,188.00] | 14.3 |
| **MET minutes per week for moderate activity (median [IQR])** | 360.00 [90.00, 880.00] | 570.00 [240.00, 960.00] | 360.00 [160.00, 840.00] | 360.00 [120.00, 1,080.00] | 14.3 |
| **MET minutes per week for vigorous activity (median [IQR])** | 240.00 [0.00, 1,110.00] | 280.00 [0.00, 960.00] | 320.00 [0.00, 960.00] | 288.00 [0.00, 960.00] | 14.3 |
| **Total MET minutes per week (median [IQR])** | 1,840.50 [1,113.00, 2,447.25 | 1,483.00 [937.50, 3,097.50] | 1,436.00 [793.00, 2,559.00] | 1,737.00 [736.88, 3,352.50] | 14.3 |
| **Family heart disease (%)** | 17 (51.5) | 47 (54.0) | 82 (61.2) | 542 (54.7) | 0 |
| CARDIAC DISEASE/OUTCOME | | | | | |
| **Cardiac problem (%)** | 0 (0.0) | 1 (1.1) | 1 (0.8) | 6 (0.6) | 0 |
| **Heart failure (%)** | 0 (0.0) | 0 (0.0) | 1 (0.8) | 5 (0.5) | 0 |
| **Cardiomyopathy (%)** | 0 (0.0) | 0 (0.0) | 0 (0.0) | 0 (0.0) | 0 |
| **Dilated cardiomyopathy (%)** | 0 (0.0) | 0 (0.0) | 0 (0.0) | 0 (0.0) | 0 |
| **Hypertrophic cardiomyopathy (%)** | 0 (0.0) | 0 (0.0) | 0 (0.0) | 0 (0.0) | 0 |
| **Ventricular arrhythmias (%)** | 0 (0.0) | 0 (0.0) | 0 (0.0) | 2 (0.2) | 0 |
| **Atrial arrhythmias (%)** | 0 (0.0) | 5 (5.7) | 3 (2.3) | 12 (1.2) | 0 |
| **Heart arrhythmia (%)** | 1 (3.0) | 3 (3.4) | 0 (0.0) | 5 (0.5) | 0 |
| **Chronic ischemic heart disease (%)** | 2 (6.1) | 3 (3.4) | 3 (2.3) | 43 (4.4) | 0 |
| **Acute myocardial infarction (%)** | 0 (0.0) | 0 (0.0) | 1 (0.8) | 29 (2.9) | 0 |
| **Cardiac arrest (%)** | 0 (0.0) | 0 (0.0) | 0 (0.0) | 1 (0.1) | 0 |
| **Angina pectoris (%)** | 1 (3.0) | 2 (2.3) | 6 (4.6) | 18 (1.8) | 0 |
| **Conduction disorders (%)** | 0 (0.0) | 0 (0.0) | 2 (1.5) | 10 (1.0) | 0 |
| **Valvular disease (%)** | 0 (0.0) | 2 (2.3) | 0 (0.0) | 23 (2.3) | 0 |
| **Congenital heart disease (%)** | 0 (0.0) | 0 (0.0) | 0 (0.0) | 0 (0.0) | 0 |
| **Pulmonary obstructive disease (%)** | 2 (6.1) | 3 (3.4) | 4 (3.1) | 35 (3.5) | 0 |
| **Cardiovascular death (%)** | 0 (0.0) | 0 (0.0) | 1 (0.8) | 3 (0.3) | 0 |
| **All-cause mortality (%)** | 0 (0.0) | 1 (1.1) | 1 (0.8) | 12 (1.2) | 0 |
| ECG MEASUREMENTS | | | | | |
| **n (%)** | 29 (87.9) | 80 (92.0) | 110 (84.6) | 856 (86.8) | 0 |
| **P duration (median [IQR])** | 100.00 [92.00, 110.00] | 98.00 [86.00, 106.00] | 100.00 [90.00, 107.00] | 100.00 [90.00, 108.00] | 16.3 |
| **P axis (median [IQR])** | 54.00 [42.25, 61.50] | 48.00 [36.00, 61.00] | 49.00 [35.00, 62.50] | 55.00 [41.00, 67.00] | 37.4 |
| **PQ interval (median [IQR])** | 171.00 [147.00, 183.00] | 165.00 [144.50, 176.00] | 160.00 [146.00, 170.00] | 160.00 [144.00, 178.00] | 37.5 |
| **QRS duration (median [IQR])** | 88.00 [80.00, 94.00] | 83.00 [78.00, 92.00] | 84.00 [80.00, 92.00] | 86.00 [80.00, 94.00] | 13 |
| **R axis (median [IQR])** | 23.50 [-1.75, 50.00] | 27.50 [-2.50, 50.00] | 40.00 [13.00, 54.00] | 35.50 [8.00, 58.00] | 35.4 |
| **QTC interval (median [IQR])** | 429.50 [403.25, 440.00] | 422.00 [405.50, 435.75] | 414.00 [401.00, 429.00] | 416.00 [402.00, 432.00] | 35.4 |
| **T axis (median [IQR])** | 35.50 [20.25, 54.25] | 42.00 [26.25, 57.00] | 45.00 [31.00, 61.00] | 40.00 [23.00, 55.75] | 35.4 |
| CMR MEASUREMENTS | | | | | |
| **RVEDVi (median [IQR])** | 79.14 [73.73, 92.49] | 76.54 [69.50, 84.81] | 77.12 [67.27, 90.71] | 80.19 [70.61, 90.27] | 8.4 |
| **RVESVi (median [IQR])** | 35.16 [29.98, 38.71] | 32.21 [27.10, 37.43] | 31.40 [26.20, 37.28] | 32.90 [27.42, 39.70] | 8.4 |
| **RVSV (median [IQR])** | 87.13 [71.60, 106.86] | 82.22 [69.02, 93.57] | 86.09 [69.79, 105.70] | 85.85 [72.77, 101.86] | 8.4 |
| **RVSVi (median [IQR])** | 48.22 [41.97, 52.24] | 44.50 [40.75, 51.29] | 45.77 [40.64, 54.04] | 46.53 [40.91, 52.81] | 8.4 |
| **RVEF (median [IQR])** | 58.30 [53.30, 62.06] | 59.31 [52.99, 62.59] | 59.56 [54.80, 63.99] | 58.37 [54.19, 62.74] | 8.7 |
| **RVPER (median [IQR])** | 405.50 [291.70, 489.35] | 361.23 [290.08, 443.96] | 389.50 [310.20, 475.80] | 388.65 [316.52, 465.95] | 8.4 |
| **RVPFR (median [IQR])** | 302.80 [225.65, 375.82] | 295.76 [220.44, 343.18] | 278.70 [230.80, 334.70] | 300.30 [245.10, 363.40] | 8.5 |
| **RVPAFR (median [IQR])** | 274.70 [213.70, 343.90] | 275.00 [224.92, 344.91] | 300.10 [236.43, 366.40] | 282.86 [222.52, 360.08] | 8.4 |
| **LVEDVi (median [IQR])** | 80.77 [73.11, 88.68] | 77.32 [68.06, 86.15] | 72.34 [64.33, 84.59] | 74.37 [66.38, 83.15] | 17.8 |
| **LVESVi (median [IQR])** | 31.74 [25.91, 39.55] | 31.69 [26.19, 39.84] | 29.37 [24.09, 34.83] | 30.02 [25.13, 35.72] | 17.8 |
| **LVSV (median [IQR])** | 91.46 [74.68, 102.10] | 80.50 [68.12, 95.54] | 82.92 [64.93, 100.22] | 81.72 [70.11, 95.72] | 17.6 |
| **LVSVi (median [IQR])** | 46.82 [43.25, 50.82] | 43.18 [37.50, 49.11] | 44.07 [38.21, 50.19] | 44.06 [39.37, 50.29] | 17.6 |
| **LVEF (median [IQR])** | 59.69 [56.59, 66.23] | 57.34 [52.60, 62.80] | 59.74 [56.25, 63.62] | 59.48 [55.29, 63.52] | 17.8 |
| **LVPER (median [IQR])** | 407.30 [307.20, 455.45] | 339.21 [259.00, 430.80] | 340.46 [264.61, 460.80] | 373.80 [302.47, 453.27] | 17.6 |
| **LVPFR (median [IQR])** | 346.20 [290.80, 422.04] | 314.20 [258.81, 366.80] | 320.25 [248.49, 371.20] | 321.50 [259.92, 385.64] | 17.6 |
| **LVPAFR (median [IQR])** | 208.70 [158.60, 298.40] | 253.35 [178.90, 330.36] | 241.28 [167.28, 321.50] | 233.50 [167.46, 305.10] | 17.7 |
| **LVEDMi (median [IQR])** | 42.81 [36.04, 48.38] | 42.96 [36.56, 46.70] | 42.41 [34.90, 49.27] | 41.85 [36.55, 48.61] | 17.7 |
| **LVMVR (median [IQR])** | 0.55 [0.49, 0.60] | 0.54 [0.49, 0.59] | 0.56 [0.50, 0.64] | 0.56 [0.50, 0.62] | 17.6 |
| **LVEDV/RVEDV (median [IQR])** | 0.94 [0.90, 1.05] | 1.00 [0.91, 1.08] | 0.94 [0.86, 0.99] | 0.93 [0.86, 1.03] | 20.3 |
| **LVESV/RVESV (median [IQR])** | 0.91 [0.82, 1.00] | 1.02 [0.89, 1.19] | 0.90 [0.82, 1.02] | 0.91 [0.80, 1.04] | 20.3 |
| **peakEcc (median [IQR])** | -22.87 [-26.91, -21.63] | -22.67 [-24.40, -19.13] | -22.91 [-25.18, -20.88] | -22.72 [-24.98, -20.42] | 36.2 |
| **TPKEcc (median [IQR])** | 326.90 [318.44, 363.82] | 334.75 [320.35, 360.47] | 331.96 [308.18, 353.62] | 331.30 [309.75, 354.70] | 36.4 |
| **peakEll2Ch (median [IQR])** | -21.37 [-23.84, -19.31] | -20.29 [-22.24, -17.97] | -21.54 [-23.50, -18.88] | -21.17 [-23.32, -18.93] | 37.3 |
| **TPKEll2Ch (median [IQR])** | 346.80 [321.40, 370.50] | 353.30 [330.98, 381.65] | 353.10 [320.40, 379.60] | 349.88 [321.78, 379.08] | 37.4 |
| **peakEll4Ch (median [IQR])** | -24.25 [-26.79, -21.38] | -22.30 [-24.57, -19.76] | -24.05 [-26.94, -22.28] | -23.30 [-25.98, -21.37] | 38.3 |
| **TPKEll4Ch (median [IQR])** | 354.30 [328.00, 406.60] | 354.80 [325.40, 392.55] | 349.21 [318.52, 390.17] | 357.30 [326.94, 397.80] | 38.6 |
| **Wall thickness segment 1 (median [IQR])** | 7.05 [6.25, 8.57] | 7.44 [6.78, 8.21] | 7.59 [6.72, 8.55] | 7.65 [6.81, 8.49] | 31.6 |
| **Wall thickness segment 2 (median [IQR])** | 6.81 [5.24, 7.75] | 6.03 [5.31, 7.39] | 7.06 [5.87, 8.21] | 6.75 [5.74, 7.90] | 31.6 |
| **Wall thickness segment 3 (median [IQR])** | 5.58 [4.74, 7.16] | 6.10 [4.85, 6.66] | 6.22 [5.04, 7.39] | 6.05 [5.17, 6.95] | 31.6 |
| **Wall thickness segment 4 (median [IQR])** | 6.06 [5.49, 6.76] | 6.57 [5.89, 6.99] | 6.47 [5.85, 6.92] | 6.54 [5.81, 7.21] | 31.6 |
| **Wall thickness segment 5 (median [IQR])** | 5.96 [5.48, 6.40] | 6.08 [5.57, 6.50] | 6.03 [5.57, 6.64] | 6.20 [5.62, 6.96] | 31.6 |

| | ARVC G+ | DCM G+ | HCM G+ | Controls G- | Missing |
|---|---|---|---|---|---|
| n | 33 | 87 | 130 | 986 | |
| **Wall thickness segment 6 (median [IQR])** | 6.45 [6.15, 6.79] | 6.58 [6.05, 7.01] | 6.56 [5.93, 7.09] | 6.55 [5.97, 7.31] | 31.6 |
| **Wall thickness segment 7 (median [IQR])** | 5.52 [5.21, 6.06] | 5.56 [5.28, 6.14] | 5.75 [5.21, 6.19] | 5.73 [5.29, 6.31] | 31.7 |
| **Wall thickness segment 8 (median [IQR])** | 6.97 [6.44, 7.28] | 6.90 [6.27, 7.47] | 6.89 [6.16, 7.49] | 7.01 [6.28, 7.75] | 31.7 |
| **Wall thickness segment 9 (median [IQR])** | 6.96 [6.18, 7.46] | 7.18 [6.64, 7.85] | 7.21 [6.38, 8.20] | 7.38 [6.48, 8.25] | 31.7 |
| **Wall thickness segment 10 (median [IQR])** | 5.88 [5.33, 6.23] | 6.28 [5.90, 6.89] | 6.18 [5.44, 7.00] | 6.24 [5.60, 6.96] | 31.7 |
| **Wall thickness segment 11 (median [IQR])** | 5.42 [4.96, 5.81] | 5.56 [5.08, 6.08] | 5.53 [5.02, 6.37] | 5.62 [5.10, 6.32] | 31.7 |
| **Wall thickness segment 12 (median [IQR])** | 5.38 [5.17, 6.12] | 5.50 [5.14, 6.02] | 5.59 [5.08, 6.13] | 5.60 [5.22, 6.26] | 31.7 |
| **Wall thickness segment 13 (median [IQR])** | 5.38 [5.24, 5.80] | 5.44 [5.13, 5.91] | 5.49 [5.12, 5.93] | 5.48 [5.10, 5.91] | 31.7 |
| **Wall thickness segment 14 (median [IQR])** | 5.92 [5.41, 6.60] | 5.90 [5.44, 6.28] | 5.95 [5.38, 6.58] | 6.00 [5.36, 6.66] | 31.7 |
| **Wall thickness segment 15 (median [IQR])** | 5.12 [4.78, 5.37] | 4.99 [4.56, 5.39] | 4.99 [4.44, 5.68] | 5.09 [4.48, 5.69] | 31.7 |
| **Wall thickness segment 16 (median [IQR])** | 5.32 [4.86, 5.67] | 5.13 [4.82, 5.67] | 5.23 [4.76, 5.65] | 5.26 [4.80, 5.72] | 31.7 |
| **Global wall thickness (median [IQR])** | 6.07 [5.57, 6.29] | 6.17 [5.90, 6.59] | 6.28 [5.70, 6.87] | 6.30 [5.72, 6.84] | 31.6 |
| **Septal wall thickness (median [IQR])** | 6.43 [5.69, 7.01] | 6.37 [6.00, 6.90] | 6.62 [6.12, 7.52] | 6.67 [5.88, 7.34] | 31.7 |
| **Maximum wall thickness (median [IQR])** | 7.81 [6.97, 8.59] | 8.01 [7.43, 8.66] | 8.16 [7.45, 9.58] | 8.09 [7.24, 9.01] | 31.7 |

Abbreviations:
ARVC: arrythmogenic right ventricular cardiomyopathy; BMI: body mass index; CMR: cardiac magnetic resonance imaging; DCM: dilated cardiomyopathy; ECG: Electrocardiography;
EDVi: indexed end-diastolic volume; EDMi: indexed end-diastolic mass; EF: ejection fraction; ESVi: indexed end-systolic volume;
G+: carriers of likely pathogenic and pathogenic variants associated with one of the cardiomyopathies; HCM: hypertrophic cardiomyopathy; HDL: high-density lipoprotein;
IQR: interquartile range; LDL: low-density lipoprotein; LV: left ventricular; MET: metabolic equivalent of task; MVR: mass to volume ratio; PAFR: peak atrial filling rate;
peakEcc: peak circumferential strain; peakEll2Ch: longitudinal strain analyzed in 2-chamber view; peakEll4Ch: longitudinal strain analyzed in 4-chamber view; PER: peak ejection rate;
PFR: peak filling rate; RV: right ventricular; SVi: indexed stroke volume; TPKEcc: global time to peak circumferential strain;
TPKEll2Ch: global time to longitudinal strain analyzed in 2-chamber view; TPKEll4Ch: global time to longitudinal strain analyzed in 4-chamber view.

| Supplementary Table X: Outcome risk stratified by cardiomyopathy and gene | | | | | | | | | | | | | | | |
|---|---|---|---|---|---|---|---|---|---|---|---|---|---|---|---|
| | ARVC G+ vs G- | | | | | DCM G+ vs G- | | | | | HCM G+ vs G- | | | | |
| Phenotype | OR | 95% LCI | 95% UCI | p-value | Gene | OR | 95% LCI | 95% UCI | p-value | Gene | OR | 95% LCI | 95% UCI | p-value | Gene |
| Heart failure | 8.270 | 0.900 | 36.898 | 0.030 | | 0.000 | 0.000 | 2031.506 | 1.000 | | 0.000 | 0.000 | 2031.506 | 1.000 | |
| Cardiomyopathy | 19.151 | 0.442 | 132.223 | 0.056 | | 0.000 | 0.000 | 9000.382 | 1.000 | | 0.000 | 0.000 | 9000.382 | 1.000 | |
| Heart failure + Cardiomyopathy | 7.476 | 0.814 | 33.321 | 0.036 | | 0.000 | 0.000 | 1841.531 | 1.000 | | 0.000 | 0.000 | 1841.531 | 1.000 | |
| Phenotype positive | 8.110 | 0.190 | 54.456 | 0.124 | | 0.000 | 0.000 | 4144.720 | 1.000 | | 0.000 | 0.000 | 4144.720 | 1.000 | |
| Ventricular arrhythmias | 0.000 | 0.000 | 88.063 | 1.000 | | 0.000 | 0.000 | 9925.577 | 1.000 | | 0.000 | 0.000 | 9925.577 | 1.000 | |
| Atrial arrhythmias | 3.657 | 0.086 | 24.275 | 0.253 | *DES* | 0.000 | 0.000 | 1936.952 | 1.000 | *ACTC1* | 0.000 | 0.000 | 1936.952 | 1.000 | *ACTC1* |
| Heart arrhythmia | 28.201 | 3.018 | 128.955 | 0.003 | | 0.000 | 0.000 | 6442.674 | 1.000 | | 0.000 | 0.000 | 6442.674 | 1.000 | |
| Chronic ischemic heart disease | 3.188 | 0.576 | 11.850 | 0.091 | | 0.000 | 0.000 | 493.649 | 1.000 | | 0.000 | 0.000 | 493.649 | 1.000 | |
| Angina pectoris | 2.211 | 0.052 | 14.617 | 0.380 | | 0.000 | 0.000 | 1185.278 | 1.000 | | 0.000 | 0.000 | 1185.278 | 1.000 | |
| Cardiovascular death | 3.863 | 0.091 | 25.642 | 0.241 | | 0.000 | 0.000 | 2042.573 | 1.000 | | 0.000 | 0.000 | 2042.573 | 1.000 | |
| All-cause mortality | 2.836 | 0.310 | 12.580 | 0.179 | | 0.000 | 0.000 | 711.200 | 1.000 | | 0.000 | 0.000 | 711.200 | 1.000 | |
| Heart failure | 1.312 | 0.032 | 7.822 | 0.540 | | 10.751 | 0.226 | 96.809 | 0.105 | | 0.000 | 0.000 | 7.948 | 1.000 | |
| Cardiomyopathy | 0.000 | 0.000 | 25.853 | 1.000 | | 0.000 | 0.000 | 235.208 | 1.000 | | 0.000 | 0.000 | 41.147 | 1.000 | |
| Heart failure + Cardiomyopathy | 1.186 | 0.029 | 7.061 | 0.576 | | 9.715 | 0.205 | 87.209 | 0.116 | | 0.000 | 0.000 | 7.178 | 1.000 | |
| Phenotype positive | 0.000 | 0.000 | 10.646 | 1.000 | | 0.000 | 0.000 | 97.932 | 1.000 | | 0.000 | 0.000 | 16.960 | 1.000 | |
| Ventricular arrhythmias | 0.000 | 0.000 | 29.170 | 1.000 | | 0.000 | 0.000 | 264.754 | 1.000 | | 0.000 | 0.000 | 46.435 | 1.000 | |
| Atrial arrhythmias | 0.000 | 0.000 | 4.745 | 1.000 | *DSC2* | 0.000 | 0.000 | 43.769 | 1.000 | *ACTN2* | 0.000 | 0.000 | 7.565 | 1.000 | *CSRP3* |
| Heart arrhythmia | 4.479 | 0.109 | 27.435 | 0.207 | | 0.000 | 0.000 | 159.535 | 1.000 | | 0.000 | 0.000 | 27.753 | 1.000 | |
| Chronic ischemic heart disease | 0.638 | 0.075 | 2.468 | 0.767 | | 2.551 | 0.054 | 22.836 | 0.365 | | 0.491 | 0.012 | 2.995 | 0.719 | |
| Angina pectoris | 0.755 | 0.019 | 4.480 | 1.000 | | 0.000 | 0.000 | 26.400 | 1.000 | | 0.000 | 0.000 | 4.558 | 1.000 | |
| Cardiovascular death | 1.319 | 0.032 | 7.866 | 0.538 | | 10.812 | 0.227 | 97.361 | 0.105 | | 0.000 | 0.000 | 7.993 | 1.000 | |
| All-cause mortality | 0.922 | 0.108 | 3.572 | 1.000 | | 3.687 | 0.078 | 33.037 | 0.272 | | 0.000 | 0.000 | 2.709 | 0.399 | |
| Heart failure | 0.000 | 0.000 | 6.863 | 1.000 | | 8.270 | 0.900 | 36.898 | 0.030 | | 0.000 | 0.000 | 45.996 | 1.000 | |
| Cardiomyopathy | 0.000 | 0.000 | 35.539 | 1.000 | | 41.175 | 4.362 | 192.166 | 0.002 | | 0.000 | 0.000 | 235.208 | 1.000 | |
| Heart failure + Cardiomyopathy | 0.000 | 0.000 | 6.196 | 1.000 | | 7.476 | 0.814 | 33.321 | 0.036 | | 0.000 | 0.000 | 41.524 | 1.000 | |
| Phenotype positive | 0.000 | 0.000 | 14.641 | 1.000 | | 17.455 | 1.885 | 78.723 | 0.008 | | 0.000 | 0.000 | 97.932 | 1.000 | |
| Ventricular arrhythmias | 0.000 | 0.000 | 40.048 | 1.000 | | 21.465 | 0.494 | 148.888 | 0.050 | | 0.000 | 0.000 | 264.754 | 1.000 | |
| Atrial arrhythmias | 0.000 | 0.000 | 6.530 | 1.000 | *DSG2* | 3.657 | 0.086 | 24.275 | 0.253 | *BAG3* | 10.231 | 0.215 | 92.112 | 0.110 | *JPH2* |
| Heart arrhythmia | 0.000 | 0.000 | 23.957 | 1.000 | | 13.101 | 0.305 | 89.207 | 0.080 | | 0.000 | 0.000 | 159.535 | 1.000 | |
| Chronic ischemic heart disease | 2.453 | 0.733 | 6.515 | 0.072 | | 0.911 | 0.022 | 6.004 | 1.000 | | 2.551 | 0.054 | 22.836 | 0.365 | |
| Angina pectoris | 2.135 | 0.246 | 8.503 | 0.254 | | 0.000 | 0.000 | 8.675 | 1.000 | | 15.464 | 1.394 | 108.566 | 0.014 | |
| Cardiovascular death | 0.000 | 0.000 | 6.902 | 1.000 | | 3.863 | 0.091 | 25.642 | 0.241 | | 0.000 | 0.000 | 46.258 | 1.000 | |
| All-cause mortality | 1.272 | 0.147 | 5.051 | 0.673 | | 1.317 | 0.031 | 8.689 | 0.548 | | 0.000 | 0.000 | 15.706 | 1.000 | |
| Heart failure | 1.121 | 0.028 | 6.628 | 0.596 | | 3.507 | 0.691 | 11.071 | 0.061 | | 1.609 | 0.966 | 2.554 | 0.047 | |
| Cardiomyopathy | 5.591 | 0.135 | 34.667 | 0.170 | | 5.591 | 0.135 | 34.667 | 0.170 | | 6.075 | 3.139 | 11.256 | 2.30E-07 | |
| Heart failure + Cardiomyopathy | 2.068 | 0.242 | 7.995 | 0.261 | | 3.170 | 0.625 | 9.992 | 0.077 | | 2.547 | 1.720 | 3.681 | 4.50E-06 | |
| Phenotype positive | 4.833 | 0.560 | 18.977 | 0.070 | | 2.367 | 0.058 | 14.183 | 0.352 | | 3.732 | 2.234 | 6.010 | 9.20E-07 | |
| Ventricular arrhythmias | 0.000 | 0.000 | 24.850 | 1.000 | | 12.800 | 1.448 | 52.549 | 0.013 | | 2.097 | 0.637 | 5.428 | 0.180 | |
| Atrial arrhythmias | 0.000 | 0.000 | 4.042 | 1.000 | *DSP* | 4.551 | 1.177 | 12.667 | 0.015 | *DES* | 1.011 | 0.540 | 1.751 | 0.889 | *MYBPC3* |
| Heart arrhythmia | 0.000 | 0.000 | 14.836 | 1.000 | | 7.811 | 0.897 | 31.194 | 0.031 | | 0.509 | 0.060 | 1.939 | 0.588 | |
| Chronic ischemic heart disease | 2.126 | 0.803 | 4.798 | 0.088 | | 1.780 | 0.617 | 4.218 | 0.169 | | 1.030 | 0.757 | 1.375 | 0.824 | |
| Angina pectoris | 2.752 | 0.714 | 7.618 | 0.068 | | 0.645 | 0.016 | 3.797 | 1.000 | | 1.294 | 0.846 | 1.913 | 0.188 | |
| Cardiovascular death | 2.302 | 0.269 | 8.907 | 0.225 | | 1.127 | 0.028 | 6.666 | 0.594 | | 0.529 | 0.209 | 1.118 | 0.106 | |
| All-cause mortality | 0.785 | 0.092 | 3.011 | 1.000 | | 1.202 | 0.238 | 3.761 | 0.740 | | 0.660 | 0.420 | 0.995 | 0.043 | |
| Heart failure | 0.000 | 0.000 | 9.023 | 1.000 | | 1.121 | 0.028 | 6.628 | 0.596 | | 1.673 | 0.656 | 3.576 | 0.208 | |
| Cardiomyopathy | 0.000 | 0.000 | 46.694 | 1.000 | | 5.591 | 0.135 | 34.667 | 0.170 | | 9.583 | 3.810 | 21.218 | 6.60E-06 | |
| Heart failure + Cardiomyopathy | 0.000 | 0.000 | 8.143 | 1.000 | | 2.068 | 0.242 | 7.995 | 0.261 | | 3.360 | 1.814 | 5.798 | 1.20E-04 | |
| Phenotype positive | 0.000 | 0.000 | 19.247 | 1.000 | | 4.833 | 0.560 | 18.977 | 0.070 | | 5.116 | 2.338 | 10.038 | 6.80E-05 | |
| Ventricular arrhythmias | 0.000 | 0.000 | 52.650 | 1.000 | | 0.000 | 0.000 | 24.850 | 1.000 | | 2.618 | 0.303 | 10.344 | 0.189 | |
| Atrial arrhythmias | 0.000 | 0.000 | 8.586 | 1.000 | *JUP* | 0.000 | 0.000 | 4.042 | 1.000 | *DSP* | 1.829 | 0.769 | 3.740 | 0.094 | *MYH7* |
| Heart arrhythmia | 0.000 | 0.000 | 31.449 | 1.000 | | 0.000 | 0.000 | 14.836 | 1.000 | | 0.795 | 0.020 | 4.666 | 1.000 | |
| Chronic ischemic heart disease | 0.555 | 0.013 | 3.425 | 1.000 | | 2.126 | 0.803 | 4.798 | 0.088 | | 1.008 | 0.573 | 1.665 | 0.899 | |
| Angina pectoris | 1.346 | 0.033 | 8.345 | 0.534 | | 2.752 | 0.714 | 7.618 | 0.068 | | 1.395 | 0.653 | 2.648 | 0.337 | |
| Cardiovascular death | 0.000 | 0.000 | 9.074 | 1.000 | | 3.527 | 0.695 | 11.134 | 0.061 | | 0.949 | 0.254 | 2.506 | 1.000 | |
| All-cause mortality | 0.000 | 0.000 | 3.074 | 0.633 | | 1.202 | 0.238 | 3.761 | 0.740 | | 1.458 | 0.827 | 2.413 | 0.135 | |
| Heart failure | 1.502 | 0.476 | 3.637 | 0.395 | | 3.044 | 0.603 | 9.532 | 0.084 | | 0.000 | 0.000 | 10.428 | 1.000 | |
| Cardiomyopathy | 1.467 | 0.036 | 8.797 | 0.501 | | 0.000 | 0.000 | 19.186 | 1.000 | | 0.000 | 0.000 | 53.909 | 1.000 | |
| Heart failure + Cardiomyopathy | 1.638 | 0.586 | 3.700 | 0.281 | | 2.751 | 0.545 | 8.603 | 0.105 | | 0.000 | 0.000 | 9.415 | 1.000 | |
| Phenotype positive | 0.621 | 0.015 | 3.596 | 1.000 | | 4.207 | 0.489 | 16.404 | 0.088 | | 0.000 | 0.000 | 22.239 | 1.000 | |
| Ventricular arrhythmias | 11.900 | 4.383 | 27.862 | 6.40E-06 | | 0.000 | 0.000 | 21.656 | 1.000 | | 0.000 | 0.000 | 60.833 | 1.000 | |
| Atrial arrhythmias | 1.726 | 0.617 | 3.902 | 0.174 | *PKP2* | 1.897 | 0.222 | 7.284 | 0.293 | *FLNC* | 2.560 | 0.061 | 16.167 | 0.335 | *MYL2* |
| Heart arrhythmia | 3.044 | 0.603 | 9.522 | 0.084 | | 0.000 | 0.000 | 12.921 | 1.000 | | 0.000 | 0.000 | 36.379 | 1.000 | |
| Chronic ischemic heart disease | 1.298 | 0.734 | 2.157 | 0.315 | | 0.981 | 0.257 | 2.678 | 1.000 | | 1.343 | 0.151 | 5.582 | 0.663 | |
| Angina pectoris | 1.224 | 0.481 | 2.606 | 0.522 | | 0.000 | 0.000 | 2.121 | 0.422 | | 1.548 | 0.037 | 9.738 | 0.488 | |
| Cardiovascular death | 2.139 | 0.836 | 4.591 | 0.086 | | 2.003 | 0.235 | 7.700 | 0.272 | | 0.000 | 0.000 | 10.487 | 1.000 | |
| All-cause mortality | 1.172 | 0.571 | 2.169 | 0.612 | | 2.634 | 1.001 | 5.884 | 0.025 | | 0.922 | 0.022 | 5.787 | 1.000 | |
| Heart failure | 0.000 | 0.000 | 2031.506 | 1.000 | | 1.312 | 0.032 | 7.822 | 0.540 | | 0.000 | 0.000 | 2031.506 | 1.000 | |
| Cardiomyopathy | 0.000 | 0.000 | 9000.382 | 1.000 | | 0.000 | 0.000 | 25.853 | 1.000 | | 0.000 | 0.000 | 9000.382 | 1.000 | |
| Heart failure + Cardiomyopathy | 0.000 | 0.000 | 1841.531 | 1.000 | | 1.186 | 0.029 | 7.061 | 0.576 | | 0.000 | 0.000 | 1841.531 | 1.000 | |
| Phenotype positive | 0.000 | 0.000 | 4144.720 | 1.000 | | 0.000 | 0.000 | 10.646 | 1.000 | | 0.000 | 0.000 | 4144.720 | 1.000 | |
| Ventricular arrhythmias | 0.000 | 0.000 | 9925.577 | 1.000 | | 15.041 | 1.692 | 62.322 | 0.009 | | 0.000 | 0.000 | 9925.577 | 1.000 | |
| Atrial arrhythmias | 0.000 | 0.000 | 1936.952 | 1.000 | *PLN* | 2.560 | 0.298 | 9.995 | 0.194 | *LMNA* | 0.000 | 0.000 | 1936.952 | 1.000 | *MYL3* |
| Heart arrhythmia | 0.000 | 0.000 | 6442.674 | 1.000 | | 14.115 | 2.708 | 46.576 | 0.002 | | 0.000 | 0.000 | 6442.674 | 1.000 | |
| Chronic ischemic heart disease | 0.000 | 0.000 | 493.649 | 1.000 | | 1.343 | 0.347 | 3.744 | 0.545 | | 0.000 | 0.000 | 493.649 | 1.000 | |
| Angina pectoris | 0.000 | 0.000 | 1185.278 | 1.000 | | 0.755 | 0.019 | 4.480 | 1.000 | | 0.000 | 0.000 | 1185.278 | 1.000 | |
| Cardiovascular death | 0.000 | 0.000 | 2042.573 | 1.000 | | 0.000 | 0.000 | 5.015 | 1.000 | | 0.000 | 0.000 | 2042.573 | 1.000 | |
| All-cause mortality | 0.000 | 0.000 | 711.200 | 1.000 | | 0.450 | 0.011 | 2.662 | 0.724 | | 0.000 | 0.000 | 711.200 | 1.000 | |
| Heart failure | | | | | | 1.758 | 0.556 | 4.268 | 0.219 | | 8.961 | 0.194 | 74.405 | 0.122 | |
| Cardiomyopathy | | | | | | 12.440 | 4.605 | 28.856 | 4.60E-06 | | 0.000 | 0.000 | 192.656 | 1.000 | |
| Heart failure + Cardiomyopathy | | | | | | 3.994 | 1.985 | 7.346 | 1.20E-04 | | 8.097 | 0.175 | 67.326 | 0.133 | |
| Phenotype positive | | | | | | 6.057 | 2.490 | 12.775 | 1.10E-04 | | 0.000 | 0.000 | 79.918 | 1.000 | |
| Ventricular arrhythmias | | | | | | 3.860 | 0.445 | 15.325 | 0.103 | | 0.000 | 0.000 | 216.918 | 1.000 | |
| Atrial arrhythmias | | | | | | 2.021 | 0.721 | 4.584 | 0.132 | *MYH7* | 0.000 | 0.000 | 35.791 | 1.000 | *TNNC1* |
| Heart arrhythmia | | | | | | 1.170 | 0.029 | 6.896 | 0.580 | | 0.000 | 0.000 | 130.632 | 1.000 | |
| Chronic ischemic heart disease | | | | | | 1.240 | 0.658 | 2.163 | 0.439 | | 2.126 | 0.046 | 17.555 | 0.411 | |
| Angina pectoris | | | | | | 1.870 | 0.831 | 3.693 | 0.101 | | 5.158 | 0.112 | 42.747 | 0.200 | |
| Cardiovascular death | | | | | | 1.047 | 0.212 | 3.166 | 0.765 | | 0.000 | 0.000 | 37.806 | 1.000 | |
| All-cause mortality | | | | | | 1.653 | 0.853 | 2.943 | 0.100 | | 7.373 | 0.700 | 45.159 | 0.047 | |
| Heart failure | | | | | | 0.000 | 0.000 | 82.035 | 1.000 | | 0.000 | 0.000 | 4.160 | 1.000 | |
| Cardiomyopathy | | | | | | 0.000 | 0.000 | 415.844 | 1.000 | | 5.478 | 0.132 | 33.866 | 0.173 | |
| Heart failure + Cardiomyopathy | | | | | | 0.000 | 0.000 | 73.980 | 1.000 | | 0.992 | 0.025 | 5.856 | 1.000 | |
| Phenotype positive | | | | | | 0.000 | 0.000 | 174.556 | 1.000 | | 2.318 | 0.057 | 13.880 | 0.357 | |
| Ventricular arrhythmias | | | | | | 0.000 | 0.000 | 465.410 | 1.000 | | 6.143 | 0.148 | 38.292 | 0.157 | |
| Atrial arrhythmias | | | | | | 0.000 | 0.000 | 77.944 | 1.000 | *NEXN* | 1.045 | 0.026 | 6.172 | 0.621 | *TNNI3* |
| Heart arrhythmia | | | | | | 0.000 | 0.000 | 282.879 | 1.000 | | 0.000 | 0.000 | 14.529 | 1.000 | |
| Chronic ischemic heart disease | | | | | | 0.000 | 0.000 | 19.338 | 1.000 | | 0.814 | 0.162 | 2.539 | 1.000 | |
| Angina pectoris | | | | | | 0.000 | 0.000 | 47.053 | 1.000 | | 1.976 | 0.391 | 6.192 | 0.207 | |
| Cardiovascular death | | | | | | 0.000 | 0.000 | 82.505 | 1.000 | | 1.104 | 0.027 | 6.523 | 0.601 | |
| All-cause mortality | | | | | | 0.000 | 0.000 | 27.978 | 1.000 | | 1.177 | 0.234 | 3.676 | 0.743 | |
| Heart failure | | | | | | 0.000 | 0.000 | 31.741 | 1.000 | | 1.000 | 0.318 | 2.406 | 1.000 | |
| Cardiomyopathy | | | | | | 0.000 | 0.000 | 163.294 | 1.000 | | 1.974 | 0.229 | 7.721 | 0.280 | |
| Heart failure + Cardiomyopathy | | | | | | 0.000 | 0.000 | 28.673 | 1.000 | | 1.088 | 0.391 | 2.442 | 0.826 | |
| Phenotype positive | | | | | | 0.000 | 0.000 | 67.578 | 1.000 | | 0.416 | 0.010 | 2.401 | 0.733 | |
| Ventricular arrhythmias | | | | | | 0.000 | 0.000 | 183.598 | 1.000 | | 0.000 | 0.000 | 4.318 | 1.000 | |
| Atrial arrhythmias | | | | | | 0.000 | 0.000 | 30.192 | 1.000 | *PLN* | 1.343 | 0.527 | 2.859 | 0.376 | *TNNT2* |
| Heart arrhythmia | | | | | | 0.000 | 0.000 | 110.596 | 1.000 | | 0.673 | 0.017 | 3.943 | 1.000 | |
| Chronic ischemic heart disease | | | | | | 0.000 | 0.000 | 7.488 | 1.000 | | 0.791 | 0.443 | 1.319 | 0.410 | |
| Angina pectoris | | | | | | 0.000 | 0.000 | 18.219 | 1.000 | | 1.173 | 0.550 | 2.220 | 0.596 | |
| Cardiovascular death | | | | | | 0.000 | 0.000 | 31.922 | 1.000 | | 1.418 | 0.557 | 3.023 | 0.355 | |
| All-cause mortality | | | | | | 0.000 | 0.000 | 10.830 | 1.000 | | 1.068 | 0.585 | 1.812 | 0.781 | |

| | ARVC G+ vs G- | | | | | DCM G+ vs G- | | | | | HCM G+ vs G- | | | | |
|---|---|---|---|---|---|---|---|---|---|---|---|---|---|---|---|
| Phenotype | OR | 95% LCI | 95% UCI | p-value | Gene | OR | 95% LCI | 95% UCI | p-value | Gene | OR | 95% LCI | 95% UCI | p-value | Gene |
| **Heart failure** | | | | | | 0.000 | 0.000 | 59.126 | 1.000 | | 0.000 | 0.000 | 45.996 | 1.000 | |
| **Cardiomyopathy** | | | | | | 0.000 | 0.000 | 301.276 | 1.000 | | 0.000 | 0.000 | 235.208 | 1.000 | |
| **Heart failure + Cardiomyopathy** | | | | | | 0.000 | 0.000 | 53.348 | 1.000 | | 0.000 | 0.000 | 41.524 | 1.000 | |
| **Phenotype positive** | | | | | | 0.000 | 0.000 | 125.163 | 1.000 | | 0.000 | 0.000 | 97.932 | 1.000 | |
| **Ventricular arrhythmias** | | | | | | 0.000 | 0.000 | 339.088 | 1.000 | | 0.000 | 0.000 | 264.754 | 1.000 | |
| **Atrial arrhythmias** | | | | | | 0.000 | 0.000 | 56.235 | 1.000 | *RBM20* | 0.000 | 0.000 | 43.769 | 1.000 | *TPM1* |
| **Heart arrhythmia** | | | | | | 0.000 | 0.000 | 204.364 | 1.000 | | 0.000 | 0.000 | 159.535 | 1.000 | |
| **Chronic ischemic heart disease** | | | | | | 0.000 | 0.000 | 13.944 | 1.000 | | 0.000 | 0.000 | 10.853 | 1.000 | |
| **Angina pectoris** | | | | | | 0.000 | 0.000 | 33.910 | 1.000 | | 0.000 | 0.000 | 26.400 | 1.000 | |
| **Cardiovascular death** | | | | | | 0.000 | 0.000 | 59.466 | 1.000 | | 0.000 | 0.000 | 46.258 | 1.000 | |
| **All-cause mortality** | | | | | | 0.000 | 0.000 | 20.179 | 1.000 | | 0.000 | 0.000 | 15.706 | 1.000 | |
| **Heart failure** | | | | | | 0.927 | 0.023 | 5.440 | 1.000 | | | | | | |
| **Cardiomyopathy** | | | | | | 0.000 | 0.000 | 18.183 | 1.000 | | | | | | |
| **Heart failure + Cardiomyopathy** | | | | | | 0.838 | 0.021 | 4.912 | 1.000 | | | | | | |
| **Phenotype positive** | | | | | | 1.959 | 0.048 | 11.638 | 0.406 | | | | | | |
| **Ventricular arrhythmias** | | | | | | 5.191 | 0.126 | 32.118 | 0.182 | | | | | | |
| **Atrial arrhythmias** | | | | | | 0.883 | 0.022 | 5.177 | 1.000 | *SCN5A* | | | | | |
| **Heart arrhythmia** | | | | | | 3.166 | 0.077 | 19.105 | 0.278 | | | | | | |
| **Chronic ischemic heart disease** | | | | | | 1.444 | 0.505 | 3.372 | 0.444 | | | | | | |
| **Angina pectoris** | | | | | | 0.000 | 0.000 | 2.010 | 0.265 | | | | | | |
| **Cardiovascular death** | | | | | | 0.000 | 0.000 | 3.525 | 0.629 | | | | | | |
| **All-cause mortality** | | | | | | 0.988 | 0.197 | 3.055 | 1.000 | | | | | | |
| **Heart failure** | | | | | | 8.961 | 0.194 | 74.405 | 0.122 | | | | | | |
| **Cardiomyopathy** | | | | | | 0.000 | 0.000 | 192.656 | 1.000 | | | | | | |
| **Heart failure + Cardiomyopathy** | | | | | | 8.097 | 0.175 | 67.326 | 0.133 | | | | | | |
| **Phenotype positive** | | | | | | 0.000 | 0.000 | 79.918 | 1.000 | | | | | | |
| **Ventricular arrhythmias** | | | | | | 0.000 | 0.000 | 216.918 | 1.000 | | | | | | |
| **Atrial arrhythmias** | | | | | | 0.000 | 0.000 | 35.791 | 1.000 | *TNNC1* | | | | | |
| **Heart arrhythmia** | | | | | | 0.000 | 0.000 | 130.632 | 1.000 | | | | | | |
| **Chronic ischemic heart disease** | | | | | | 2.126 | 0.046 | 17.555 | 0.411 | | | | | | |
| **Angina pectoris** | | | | | | 5.158 | 0.112 | 42.747 | 0.200 | | | | | | |
| **Cardiovascular death** | | | | | | 0.000 | 0.000 | 37.806 | 1.000 | | | | | | |
| **All-cause mortality** | | | | | | 7.373 | 0.700 | 45.159 | 0.047 | | | | | | |
| **Heart failure** | | | | | | 0.000 | 0.000 | 6.036 | 1.000 | | | | | | |
| **Cardiomyopathy** | | | | | | 0.000 | 0.000 | 31.281 | 1.000 | | | | | | |
| **Heart failure + Cardiomyopathy** | | | | | | 0.000 | 0.000 | 5.451 | 1.000 | | | | | | |
| **Phenotype positive** | | | | | | 0.000 | 0.000 | 12.879 | 1.000 | | | | | | |
| **Ventricular arrhythmias** | | | | | | 8.852 | 0.212 | 56.217 | 0.112 | | | | | | |
| **Atrial arrhythmias** | | | | | | 1.506 | 0.037 | 9.076 | 0.493 | *TNNI3* | | | | | |
| **Heart arrhythmia** | | | | | | 0.000 | 0.000 | 21.072 | 1.000 | | | | | | |
| **Chronic ischemic heart disease** | | | | | | 1.196 | 0.234 | 3.836 | 0.739 | | | | | | |
| **Angina pectoris** | | | | | | 2.902 | 0.566 | 9.356 | 0.097 | | | | | | |
| **Cardiovascular death** | | | | | | 1.591 | 0.039 | 9.594 | 0.475 | | | | | | |
| **All-cause mortality** | | | | | | 1.117 | 0.130 | 4.390 | 0.701 | | | | | | |
| **Heart failure** | | | | | | 1.735 | 0.042 | 10.530 | 0.447 | | | | | | |
| **Cardiomyopathy** | | | | | | 8.655 | 0.207 | 54.922 | 0.115 | | | | | | |
| **Heart failure + Cardiomyopathy** | | | | | | 3.240 | 0.373 | 12.920 | 0.137 | | | | | | |
| **Phenotype positive** | | | | | | 3.664 | 0.089 | 22.504 | 0.247 | | | | | | |
| **Ventricular arrhythmias** | | | | | | 0.000 | 0.000 | 38.788 | 1.000 | | | | | | |
| **Atrial arrhythmias** | | | | | | 1.652 | 0.040 | 10.021 | 0.463 | *TNNT2* | | | | | |
| **Heart arrhythmia** | | | | | | 0.000 | 0.000 | 23.166 | 1.000 | | | | | | |
| **Chronic ischemic heart disease** | | | | | | 1.319 | 0.257 | 4.272 | 0.504 | | | | | | |
| **Angina pectoris** | | | | | | 0.999 | 0.024 | 6.034 | 1.000 | | | | | | |
| **Cardiovascular death** | | | | | | 3.605 | 0.414 | 14.395 | 0.116 | | | | | | |
| **All-cause mortality** | | | | | | 1.907 | 0.371 | 6.184 | 0.227 | | | | | | |
| **Heart failure** | | | | | | 0.000 | 0.000 | 287.327 | 1.000 | | | | | | |
| **Cardiomyopathy** | | | | | | 0.000 | 0.000 | 1454.551 | 1.000 | | | | | | |
| **Heart failure + Cardiomyopathy** | | | | | | 0.000 | 0.000 | 259.607 | 1.000 | | | | | | |
| **Phenotype positive** | | | | | | 0.000 | 0.000 | 604.765 | 1.000 | | | | | | |
| **Ventricular arrhythmias** | | | | | | 0.000 | 0.000 | 1644.031 | 1.000 | | | | | | |
| **Atrial arrhythmias** | | | | | | 0.000 | 0.000 | 273.509 | 1.000 | *TPM1* | | | | | |
| **Heart arrhythmia** | | | | | | 0.000 | 0.000 | 987.049 | 1.000 | | | | | | |
| **Chronic ischemic heart disease** | | | | | | 0.000 | 0.000 | 67.908 | 1.000 | | | | | | |
| **Angina pectoris** | | | | | | 0.000 | 0.000 | 165.176 | 1.000 | | | | | | |
| **Cardiovascular death** | | | | | | 0.000 | 0.000 | 288.947 | 1.000 | | | | | | |
| **All-cause mortality** | | | | | | 0.000 | 0.000 | 98.466 | 1.000 | | | | | | |
| **Heart failure** | | | | | | 3.585 | 2.012 | 6.014 | 2.20E-05 | | | | | | |
| **Cardiomyopathy** | | | | | | 10.241 | 4.492 | 21.287 | 3.20E-07 | | | | | | |
| **Heart failure + Cardiomyopathy** | | | | | | 3.444 | 1.969 | 5.695 | 2.10E-05 | | | | | | |
| **Phenotype positive** | | | | | | 3.442 | 1.426 | 7.187 | 0.004 | | | | | | |
| **Ventricular arrhythmias** | | | | | | 4.494 | 1.148 | 12.764 | 0.016 | | | | | | |
| **Atrial arrhythmias** | | | | | | 3.200 | 1.766 | 5.433 | 1.20E-04 | *TTN* | | | | | |
| **Heart arrhythmia** | | | | | | 2.741 | 0.716 | 7.515 | 0.068 | | | | | | |
| **Chronic ischemic heart disease** | | | | | | 1.178 | 0.728 | 1.823 | 0.477 | | | | | | |
| **Angina pectoris** | | | | | | 1.182 | 0.554 | 2.238 | 0.595 | | | | | | |
| **Cardiovascular death** | | | | | | 2.064 | 0.962 | 3.946 | 0.037 | | | | | | |
| **All-cause mortality** | | | | | | 1.229 | 0.699 | 2.027 | 0.404 | | | | | | |

Abbreviations:
*ACTC1* : Actin Alpha Cardiac Muscle 1; *ACTN2* : Alpha-actinin 2; ARVC: Arrhythmogenic right ventricular cardiomyopathy; *BAG3* : BAG Cochaperone 3; *CSRP3* : Cysteine And Glycine Rich Protein 3; DCM: Dilated cardiomyopathy; *DES* : Desmin; *DSC2* : Desmocollin 2; *DSG2* : Desmoglein 2; *DSG2* : Desmoglein 2; *DSP* : desmoplakin; *FLNC* : Filamin-C; G+: carriers of likely pathogenic and pathogenic variants associated with one of the cardiomyopathies;
HCM: Hypertrophic cardiomyopathy; *JPH2* : Junctophilin 2; *JUP* : Junction Plakoglobin; LCI: lower limit confidence interval; *LMNA* : Lamin A/C; *MYBPC3* : Myosin Binding Protein C3; *MYH7* : Myosin Heavy Chain 7;
*NEXN: Nexilin F-Actin Binding Protein; PKP2* : Plakophilin 2; *PLN* : phospholamban; *RBM20* : RNA Binding Motif Protein 20; *SCN5A* : Sodium Voltage-Gated Channel Alpha Subunit 5; *TNNC1* : Troponin C1, Slow Skeletal And Cardiac Type; *TNNI3: Troponin I3, Cardiac Type; TNNT2* : Troponin T2, Cardiac Type; *TPM1* : Tropomyosin 1; *TTN* : Titin; UCI: upper limit confidence interval.

**Supplementary Table XI: Results of Fisher's Exact tests when excluding overlapping genes**

| | ARVC G+ vs G- | | | | DCM G+ vs G- (without ARVC genes) | | | | DCM G+ vs G- (without HCM genes) | | | | HCM G+ vs G- | | | |
|---|---|---|---|---|---|---|---|---|---|---|---|---|---|---|---|---|
| | OR | 95% LCI | 95% UCI | p-value | OR | 95% LCI | 95% UCI | p-value | OR | 95% LCI | 95% UCI | p-value | OR | 95% LCI | 95% UCI | p-value |
| CARDIOVASCULAR RISK FACTORS | | | | | | | | | | | | | | | | |
| Diabetes | 1.136 | 0.741 | 1.684 | 0.530 | 0.819 | 0.602 | 1.095 | 0.194 | 0.835 | 0.595 | 1.147 | 0.293 | 1.682 | 1.348 | 2.083 | 4.20E-06 |
| Hypertension | 0.989 | 0.762 | 1.276 | 0.949 | 1.092 | 0.927 | 1.285 | 0.283 | 0.999 | 0.831 | 1.198 | 1.000 | 1.025 | 0.875 | 1.197 | 0.753 |
| Hypercholesterolaemia | 1.033 | 0.774 | 1.365 | 0.833 | 1.137 | 0.949 | 1.356 | 0.156 | 1.057 | 0.864 | 1.288 | 0.579 | 1.192 | 1.006 | 1.407 | 0.041 |
| Ever Smoked | 1.158 | 0.905 | 1.479 | 0.244 | 1.223 | 1.045 | 1.432 | 0.011 | 1.192 | 1.002 | 1.418 | 0.044 | 0.869 | 0.745 | 1.013 | 0.069 |
| Family heart disease | 1.386 | 1.086 | 1.770 | 0.007 | 1.134 | 0.969 | 1.327 | 0.114 | 1.074 | 0.902 | 1.277 | 0.434 | 1.010 | 0.869 | 1.173 | 0.910 |
| CARDIAC DISEASE/OUTCOME | | | | | | | | | | | | | | | | |
| Cardiac problem | 2.604 | 0.513 | 8.240 | 0.120 | 1.052 | 0.208 | 3.309 | 0.762 | 1.291 | 0.255 | 4.065 | 0.511 | 1.262 | 0.327 | 3.497 | 0.563 |
| Heart failure | 1.169 | 0.420 | 2.628 | 0.650 | 2.600 | 1.713 | 3.837 | 8.40E-06 | 3.010 | 1.955 | 4.497 | 1.00E-06 | 1.504 | 0.903 | 2.386 | 0.097 |
| Cardiomyopathy | 0.956 | 0.023 | 5.708 | 1.000 | 7.963 | 4.354 | 14.167 | 1.70E-10 | 6.807 | 3.379 | 12.983 | 2.90E-07 | 5.681 | 2.936 | 10.526 | 5.20E-07 |
| Dilated cardiomyopathy | 2.531 | 0.060 | 16.741 | 0.342 | 8.290 | 3.003 | 21.263 | 4.40E-05 | 11.486 | 4.365 | 28.657 | 1.40E-06 | 0.923 | 0.022 | 6.079 | 1.000 |
| Hypertrophic cardiomyopathy | 0.000 | 0.000 | 20.787 | 1.000 | 10.856 | 3.095 | 35.820 | 1.40E-04 | 2.204 | 0.050 | 16.486 | 0.39 | 17.990 | 6.569 | 51.615 | 1.10E-08 |
| Ventricular arrhythmias | 7.664 | 2.835 | 17.818 | 9.50E-05 | 4.849 | 2.201 | 9.888 | 8.70E-05 | 5.415 | 2.368 | 11.317 | 7.10E-05 | 1.963 | 0.597 | 5.081 | 0.192 |
| Atrial arrhythmias | 1.113 | 0.400 | 2.499 | 0.663 | 2.313 | 1.507 | 3.444 | 1.40E-04 | 2.465 | 1.556 | 3.766 | 1.10E-04 | 1.015 | 0.554 | 1.727 | 0.892 |
| Heart arrhythmia | 2.642 | 0.690 | 7.242 | 0.075 | 2.685 | 1.213 | 5.356 | 0.008 | 3.639 | 1.706 | 7.089 | 0.001 | 0.477 | 0.056 | 1.815 | 0.435 |
| Chronic ischemic heart disease | 1.241 | 0.783 | 1.889 | 0.297 | 1.207 | 0.901 | 1.594 | 0.176 | 1.318 | 0.964 | 1.770 | 0.069 | 1.017 | 0.754 | 1.349 | 0.886 |
| Acute myocardial infarction | 1.443 | 0.728 | 2.600 | 0.215 | 1.163 | 0.728 | 1.780 | 0.490 | 1.005 | 0.573 | 1.653 | 0.900 | 0.863 | 0.517 | 1.368 | 0.659 |
| Cardiac arrest | 0.000 | 0.000 | 4.064 | 1.000 | 2.549 | 0.872 | 6.170 | 0.043 | 2.080 | 0.534 | 5.863 | 0.145 | 1.140 | 0.224 | 3.638 | 0.747 |
| Angina pectoris | 1.257 | 0.613 | 2.316 | 0.486 | 1.157 | 0.732 | 1.756 | 0.500 | 0.901 | 0.505 | 1.500 | 0.803 | 1.252 | 0.824 | 1.839 | 0.242 |
| Conduction disorders | 1.899 | 0.797 | 3.891 | 0.084 | 1.633 | 0.921 | 2.723 | 0.080 | 1.409 | 0.708 | 2.554 | 0.289 | 1.289 | 0.700 | 2.208 | 0.361 |
| Valvular disease | 1.179 | 0.498 | 2.394 | 0.558 | 2.016 | 1.345 | 2.937 | 0.001 | 1.866 | 1.173 | 2.852 | 0.006 | 1.074 | 0.640 | 1.708 | 0.716 |
| Congenital heart disease | 2.536 | 0.291 | 10.141 | 0.199 | 1.542 | 0.299 | 5.010 | 0.452 | 1.259 | 0.145 | 5.016 | 0.675 | 1.385 | 0.269 | 4.500 | 0.486 |
| Pulmonary obstructive disease | 1.704 | 1.051 | 2.643 | 0.026 | 1.205 | 0.845 | 1.677 | 0.280 | 1.112 | 0.739 | 1.618 | 0.551 | 0.695 | 0.450 | 1.033 | 0.081 |
| Cardiovascular death | 1.579 | 0.665 | 3.223 | 0.254 | 1.605 | 0.951 | 2.574 | 0.058 | 1.879 | 1.097 | 3.049 | 0.016 | 0.495 | 0.196 | 1.046 | 0.064 |
| All-cause mortality | 1.036 | 0.567 | 1.756 | 0.891 | 1.432 | 1.037 | 1.940 | 0.022 | 1.290 | 0.888 | 1.826 | 0.145 | 0.643 | 0.413 | 0.961 | 0.032 |

Abbreviations:
ARVC: arrhythmogenic right ventricular cardiomyopathy; DCM: dilated cardiomyopathy; G+: carriers of likely pathogenic and pathogenic variants associated with one of the cardiomyopathies; HCM: hypertrophic cardiomyopathy;
LCI: lower limit confidence interval; UCI: upper limit confidence interval.

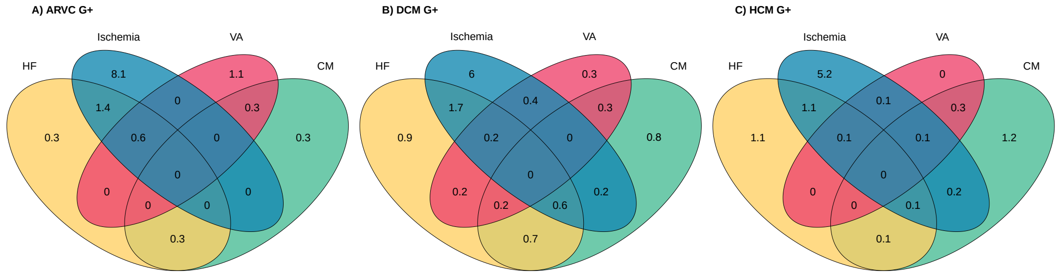
A) ARVC G+
Ischemia
VA
HF
CM
8.1
1.1
0
1.4
0.3
0.3
0.6
0
0.3
0
0
0
0
0
0.3
B) DCM G+
Ischemia
VA
HF
CM
6
0.3
0.4
1.7
0.3
0.9
0.2
0
0.8
0
0.2
0.2
0.6
0.2
0.7
C) HCM G+
Ischemia
VA
HF
CM
5.2
0
0.1
1.1
0.3
1.1
0.1
0.1
1.2
0
0
0
0.1
0.2
0.1

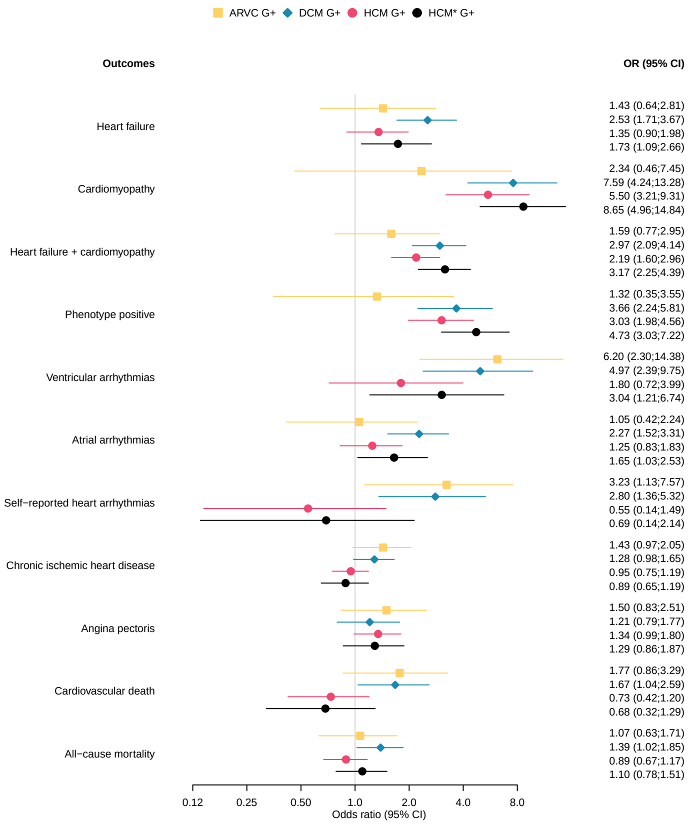
ARVC G+ DCM G+ HCM G+ HCM* G+
Outcomes
OR (95% CI)
Heart failure
1.43 (0.64;2.81)
2.53 (1.71;3.67)
1.35 (0.90;1.98)
1.73 (1.09;2.66)
Cardiomyopathy
2.34 (0.46;7.45)
7.59 (4.24;13.28)
5.50 (3.21;9.31)
8.65 (4.96;14.84)
Heart failure + cardiomyopathy
1.59 (0.77;2.95)
2.97 (2.09;4.14)
2.19 (1.60;2.96)
3.17 (2.25;4.39)
Phenotype positive
1.32 (0.35;3.55)
3.66 (2.24;5.81)
3.03 (1.98;4.56)
4.73 (3.03;7.22)
Ventricular arrhythmias
6.20 (2.30;14.38)
4.97 (2.39;9.75)
1.80 (0.72;3.99)
3.04 (1.21;6.74)
Atrial arrhythmias
1.05 (0.42;2.24)
2.27 (1.52;3.31)
1.25 (0.83;1.83)
1.65 (1.03;2.53)
Self−reported heart arrhythmias
3.23 (1.13;7.57)
2.80 (1.36;5.32)
0.55 (0.14;1.49)
0.69 (0.14;2.14)
Chronic ischemic heart disease
1.43 (0.97;2.05)
1.28 (0.98;1.65)
0.95 (0.75;1.19)
0.89 (0.65;1.19)
Angina pectoris
1.50 (0.83;2.51)
1.21 (0.79;1.77)
1.34 (0.99;1.80)
1.29 (0.86;1.87)
Cardiovascular death
1.77 (0.86;3.29)
1.67 (1.04;2.59)
0.73 (0.42;1.20)
0.68 (0.32;1.29)
All−cause mortality
1.07 (0.63;1.71)
1.39 (1.02;1.85)
0.89 (0.67;1.17)
1.10 (0.78;1.51)
0.12 0.25 0.50 1.0 2.0 4.0 8.0
Odds ratio (95% CI)

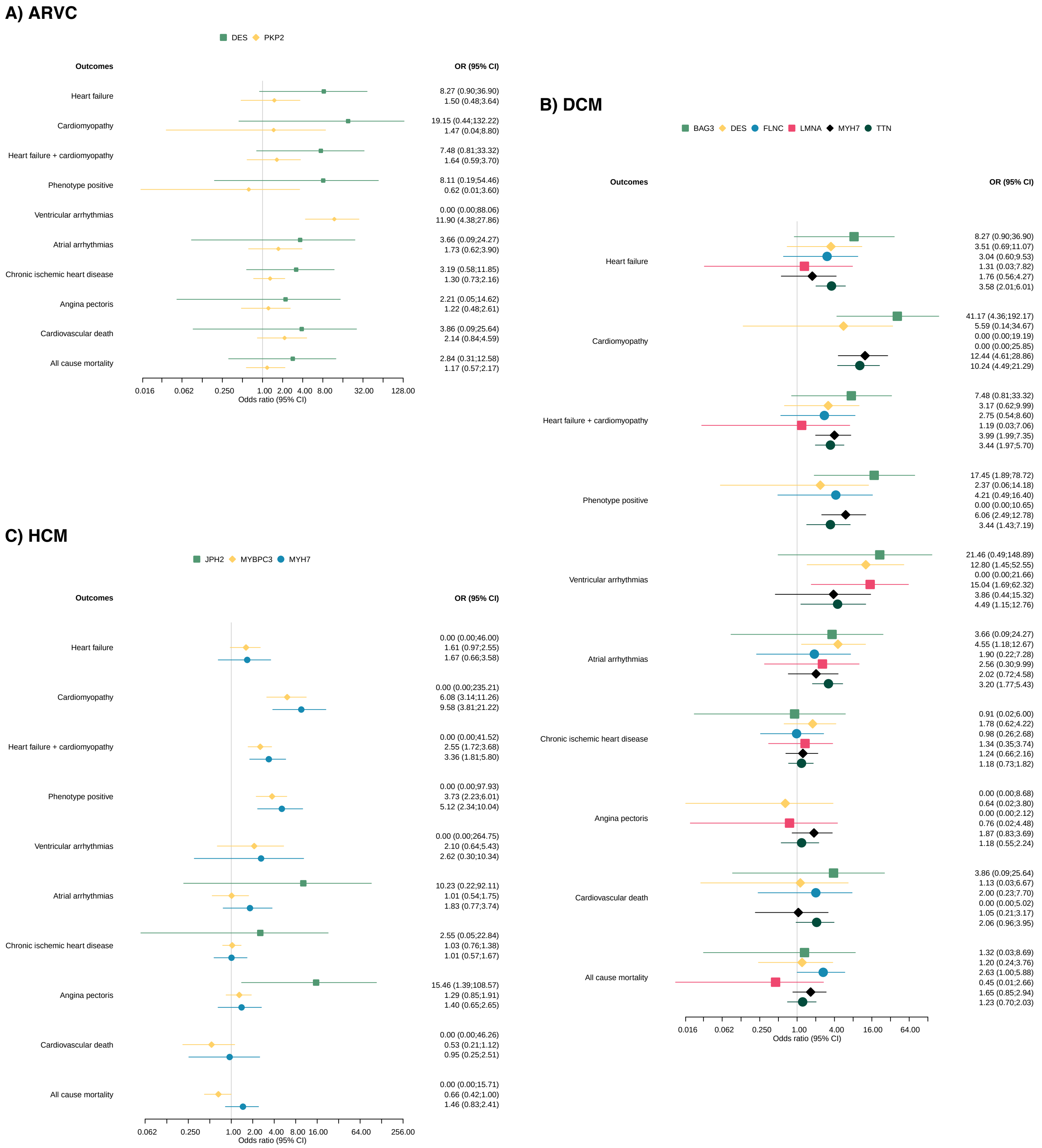
A) ARVC
DES PKP2
Outcomes
OR (95% CI)
Heart failure
8.27 (0.90;36.90)
1.50 (0.48;3.64)
Cardiomyopathy
19.15 (0.44;132.22)
1.47 (0.04;8.80)
Heart failure + cardiomyopathy
7.48 (0.81;33.32)
1.64 (0.59;3.70)
Phenotype positive
8.11 (0.19;54.46)
0.62 (0.01;3.60)
Ventricular arrhythmias
0.00 (0.00;88.06)
11.90 (4.38;27.86)
Atrial arrhythmias
3.66 (0.09;24.27)
1.73 (0.62;3.90)
Chronic ischemic heart disease
3.19 (0.58;11.85)
1.30 (0.73;2.16)
Angina pectoris
2.21 (0.05;14.62)
1.22 (0.48;2.61)
Cardiovascular death
3.86 (0.09;25.64)
2.14 (0.84;4.59)
All cause mortality
2.84 (0.31;12.58)
1.17 (0.57;2.17)
0.016 0.062 0.250 1.00 2.00 4.00 8.00 32.00 128.00
Odds ratio (95% CI)
B) DCM
BAG3 DES FLNC LMNA MYH7 TTN
Outcomes
OR (95% CI)
Heart failure
8.27 (0.90;36.90)
3.51 (0.69;11.07)
3.04 (0.60;9.53)
1.31 (0.03;7.82)
1.76 (0.56;4.27)
3.58 (2.01;6.01)
Cardiomyopathy
41.17 (4.36;192.17)
5.59 (0.14;34.67)
0.00 (0.00;19.19)
0.00 (0.00;25.85)
12.44 (4.61;28.86)
10.24 (4.49;21.29)
Heart failure + cardiomyopathy
7.48 (0.81;33.32)
3.17 (0.62;9.99)
2.75 (0.54;8.60)
1.19 (0.03;7.06)
3.99 (1.99;7.35)
3.44 (1.97;5.70)
Phenotype positive
17.45 (1.89;78.72)
2.37 (0.06;14.18)
4.21 (0.49;16.40)
0.00 (0.00;10.65)
6.06 (2.49;12.78)
3.44 (1.43;7.19)
Ventricular arrhythmias
21.46 (0.49;148.89)
12.80 (1.45;52.55)
0.00 (0.00;21.66)
15.04 (1.69;62.32)
3.86 (0.44;15.32)
4.49 (1.15;12.76)
Atrial arrhythmias
3.66 (0.09;24.27)
4.55 (1.18;12.67)
1.90 (0.22;7.28)
2.56 (0.30;9.99)
2.02 (0.72;4.58)
3.20 (1.77;5.43)
Chronic ischemic heart disease
0.91 (0.02;6.00)
1.78 (0.62;4.22)
0.98 (0.26;2.68)
1.34 (0.35;3.74)
1.24 (0.66;2.16)
1.18 (0.73;1.82)
Angina pectoris
0.00 (0.00;8.68)
0.64 (0.02;3.80)
0.00 (0.00;2.12)
0.76 (0.02;4.48)
1.87 (0.83;3.69)
1.18 (0.55;2.24)
Cardiovascular death
3.86 (0.09;25.64)
1.13 (0.03;6.67)
2.00 (0.23;7.70)
0.00 (0.00;5.02)
1.05 (0.21;3.17)
2.06 (0.96;3.95)
All cause mortality
1.32 (0.03;8.69)
1.20 (0.24;3.76)
2.63 (1.00;5.88)
0.45 (0.01;2.66)
1.65 (0.85;2.94)
1.23 (0.70;2.03)
0.016 0.062 0.250 1.00 4.00 16.00 64.00
Odds ratio (95% CI)
C) HCM
JPH2 MYBPC3 MYH7
Outcomes
OR (95% CI)
Heart failure
0.00 (0.00;46.00)
1.61 (0.97;2.55)
1.67 (0.66;3.58)
Cardiomyopathy
0.00 (0.00;235.21)
6.08 (3.14;11.26)
9.58 (3.81;21.22)
Heart failure + cardiomyopathy
0.00 (0.00;41.52)
2.55 (1.72;3.68)
3.36 (1.81;5.80)
Phenotype positive
0.00 (0.00;97.93)
3.73 (2.23;6.01)
5.12 (2.34;10.04)
Ventricular arrhythmias
0.00 (0.00;264.75)
2.10 (0.64;5.43)
2.62 (0.30;10.34)
Atrial arrhythmias
10.23 (0.22;92.11)
1.01 (0.54;1.75)
1.83 (0.77;3.74)
Chronic ischemic heart disease
2.55 (0.05;22.84)
1.03 (0.76;1.38)
1.01 (0.57;1.67)
Angina pectoris
15.46 (1.39;108.57)
1.29 (0.85;1.91)
1.40 (0.65;2.65)
Cardiovascular death
0.00 (0.00;46.26)
0.53 (0.21;1.12)
0.95 (0.25;2.51)
All cause mortality
0.00 (0.00;15.71)
0.66 (0.42;1.00)
1.46 (0.83;2.41)
0.062 0.250 1.00 2.00 4.00 8.00 16.00 64.00 256.00
Odds ratio (95% CI)

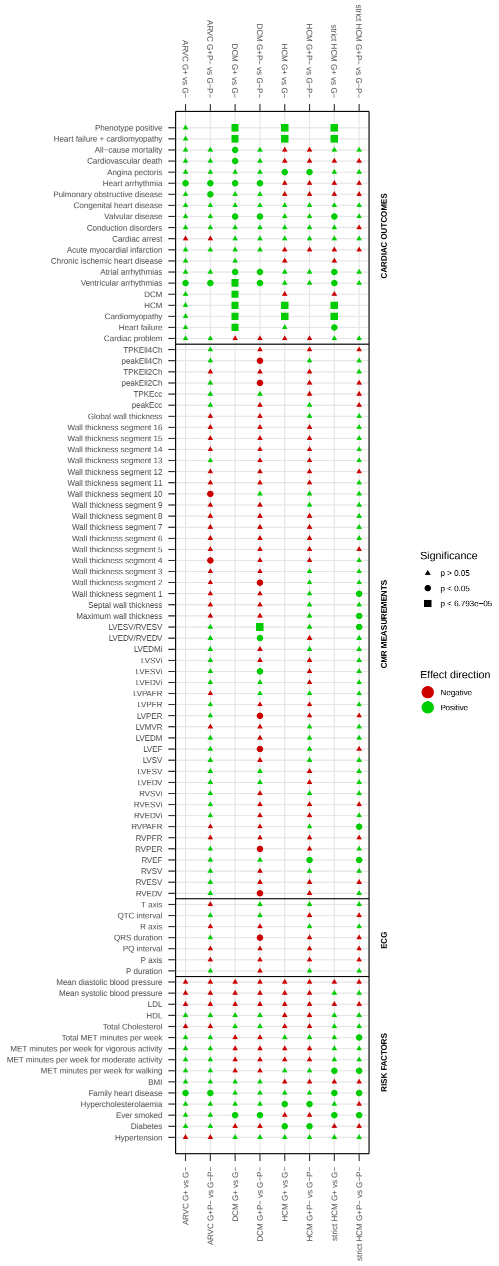
ARVC G+ vs G−
ARVC G+P− vs G−P−
DCM G+ vs G−
DCM G+P− vs G−P−
HCM G+ vs G−
HCM G+P− vs G−P−
strict HCM G+ vs G−
strict HCM G+P− vs G−P−
CARDIAC OUTCOMES
Phenotype positive
Heart failure + cardiomyopathy
All−cause mortality
Cardiovascular death
Angina pectoris
Heart arrhythmia
Pulmonary obstructive disease
Congenital heart disease
Valvular disease
Conduction disorders
Cardiac arrest
Acute myocardial infarction
Chronic ischemic heart disease
Atrial arrhythmias
Ventricular arrhythmias
DCM
HCM
Cardiomyopathy
Heart failure
Cardiac problem
CMR MEASUREMENTS
TPKEll4Ch
peakEll4Ch
TPKEll2Ch
peakEll2Ch
TPKEcc
peakEcc
Global wall thickness
Wall thickness segment 16
Wall thickness segment 15
Wall thickness segment 14
Wall thickness segment 13
Wall thickness segment 12
Wall thickness segment 11
Wall thickness segment 10
Wall thickness segment 9
Wall thickness segment 8
Wall thickness segment 7
Wall thickness segment 6
Wall thickness segment 5
Wall thickness segment 4
Wall thickness segment 3
Wall thickness segment 2
Wall thickness segment 1
Septal wall thickness
Maximum wall thickness
LVESV/RVESV
LVEDV/RVEDV
LVEDMi
LVSVi
LVESVi
LVEDVi
LVPAFR
LVPFR
LVPER
LVMVR
LVEDM
LVEF
LVSV
LVESV
LVEDV
RVSVi
RVESVi
RVEDVi
RVPAFR
RVPFR
RVPER
RVEF
RVSV
RVESV
RVEDV
ECG
T axis
QTC interval
R axis
QRS duration
PQ interval
P axis
P duration
RISK FACTORS
Mean diastolic blood pressure
Mean systolic blood pressure
LDL
HDL
Total Cholesterol
Total MET minutes per week
MET minutes per week for vigorous activity
MET minutes per week for moderate activity
MET minutes per week for walking
BMI
Family heart disease
Hypercholesterolaemia
Ever smoked
Diabetes
Hypertension
Significance
p > 0.05
p < 0.05
p < 6.793e−05
Effect direction
Negative
Positive

**Figure I: Overlap cardiac diagnoses per inherited cardiomyopathy**

The Venn diagram of the overlap between cardiomyopathy, heart failure, ventricular arrhythmia and chronic ischemic heart diagnoses in G+ individuals. The numbers in the diagram are the percentages of individuals diagnosed in the G+ of the specified cardiomyopathy.
*Abbreviations: ARVC= arrhythmogenic right ventricular cardiomyopathy; CM= cardiomyopathy; DCM= dilated cardiomyopathy; G+= pathogenic variant carrier; HCM= hypertrophic cardiomyopathy; HF= heart failure; VA= ventricular arrhythmias.*

**Figure II: Forest plot cardiac outcomes stratified per inherited cardiomyopathy**

Odds ratios and 95% confidence intervals are given for the associations between cardiac outcomes and ARVC, DCM, or HCM pathogenic variant carriers.
*Abbreviations: ARVC= arrhythmogenic right ventricular cardiomyopathy; DCM= dilated cardiomyopathy; G+= pathogenic variant carrier; HCM= hypertrophic cardiomyopathy.*
* strict HCM group: HCM group after excluding carriers of the 3628-41_3628-17del *MYBPC3* and the Arg278Cys 862C>T *TNNT2* variant.

**Figure III: Forest plot cardiac outcomes stratified per inherited cardiomyopathy and gene**

Odds ratios and 95% confidence intervals are given for the associations between cardiac outcomes and A) ARVC, B) DCM, or C) HCM pathogenic variant carriers stratified by gene. Results are only visualized for genes with at least one significant result.
*Abbreviations: ARVC= arrhythmogenic right ventricular cardiomyopathy; DCM= dilated cardiomyopathy; HCM= hypertrophic cardiomyopathy.*

**Figure IV: Matrix of all differences tested**

Summary of p-values and effect direction for all performed tests in this study. The p-value is indicated by the shape of the symbol, whereas the effect direction is represented by the color. Red color indicates an odds ratio smaller than one for categorical variables and a lower mean value for the G+ compared to the G- group for continuous variables. No symbol indicates the test was not performed.
*Abbreviations: ARVC= arrhythmogenic right ventricular cardiomyopathy; CMR= cardiac magnetic resonance imaging; DCM= dilated cardiomyopathy; ECG= Electrocardiography; G+= pathogenic variant carrier; HCM= hypertrophic cardiomyopathy; P-= phenotype negative individual; P+= phenotype positive individual.*
